# A comparison of the effectiveness of biologic therapies for asthma: a systematic review and network meta-analysis

**DOI:** 10.1101/2022.08.07.22278522

**Authors:** Tyler Pitre, Tanvir Jassal, Albi Angjeli, Vineeth Jarabana, Sricherry Nannapaneni, Ayesha Umair, Muizz Hussain, Gareth Leung, Sarah Kirsh, Johnny Su, Kairavi Desai, Jade Coyne, Sindu Mohan, Dena Zeraatkar

## Abstract

**Background:** Trials have not directly compared biologics for the treatment of asthma.

**Objective:** To comparative the relative efficacy of biologics in asthma.

**Methods:** We searched MEDLINE, EMBASE, CENTRAL, and clinicaltrials.gov from inception to May 31, 2022, for randomized trials addressing biologic therapies for asthma. Reviewers worked independently and in duplicate to screen references, extract data, and assess risk of bias. We performed a frequentist network meta-analysis and assessed the certainty of evidence using the GRADE approach. We present dichotomous outcomes as absolute risk differences per 1000 patients and relative risk (RR) with 95% confidence intervals (95% CI) and continuous outcomes as mean difference (MD) and 95% CI.

**Results:** We identified 64 trials, including 26,630 patients. For patients with eosinophilic asthma, tezepelumab (329 fewer exacerbations per 1000 [95% CI 272.6 to 366.6 fewer]) and dupilumab (319.6 fewer exacerbations per 1000 [95% CI 272.6 to 357.2 fewer]) reduce exacerbations compared to placebo (high certainty). Tezepelumab (MD 0.24 L [95% CI 0.16 to 0.32]) and dupilumab (0.25 L (95% CI 0.21 to 0.29) improve lung function (FEV1) compared to placebo (high certainty). Both tezepelumab (110.97 fewer hospital admissions per 1000 (95% CI 94.53 to 120.56 fewer) and dupilumab (97.27 fewer hospitalizations [4.11 to 124.67 fewer]) probably reduce hospital admissions compared to placebo (moderate certainty). For patients with low eosinophils, biologics probably do not improve asthma outcomes. For these patients, tezepelumab (MD 0.1 L [95% CI 0 to 0.19]) and dupilumab (MD 0.1 L [95% CI 0 to 0.20)] may improve lung function (low certainty).

**Conclusion:** Tezepelumab and dupilumab are effective at reducing exacerbations. For patients with low eosinophils, however, clinicians should probably be more judicious in use of biologics, including tezepelumab since they probably do not confer substantial benefit.

## Introduction

Asthma is a respiratory condition that affects an estimated 300 million people and is associated with significant morbidity^1^. Patients with asthma that is inadequately controlled by high dose inhaled corticosteroids and long-acting β2-agonists and patients who rely on oral corticosteroids may benefit from biologics^2^. Clinicians need to consider both disease severity and inflammatory endotype (type 2 high and type 2 low) to select optimal patients for biologic therapies.

To date, no trial has directly compared biologics for the treatment of uncontrolled asthma and the choice of biologic has been largely guided by biological and practical considerations, such as assessing inflammatory biomarkers (e.g., blood and sputum eosinophils, fractional exhaled nitric oxide, IgE levels) and access and payer coverage. Additionally, the optimal biologic for patients who qualify for more than one option and the order in which clinicians should trial biologics for patients are unclear^3^. Access to most biologics has been limited to patients with specific clinical phenotypes such as allergic and nonallergic eosinophilic asthma. The emergence of anti-alarmins—treatments that should theoretically be effective regardless of eosinophil levels—further necessitates additional evidence to help select optimal biologics for patients^4^.

To address these challenges, we present a systematic review and network meta-analysis comparing biologics for severe uncontrolled asthma. A network meta-analysis pools results across two or more studies and, unlike a traditional pairwise meta-analysis, facilitates comparisons of three or more interventions simultaneously^5^. A network meta-analysis can also yield more precise estimates than a traditional pairwise meta-analysis and can facilitate comparison of interventions that have not been directly compared against each other. Our network meta-analysis improves on previous reviews on the topic by including the last trial data not included in previous reviews, by considering eosinophil levels, and by using the GRADE guidance to interpret and contextualize findings^6-12^.

## Methods

We registered our protocol on Open Science Framework (https://osf.io/m29du/) on June 14, 2022. We report our results using the PRISMA-NMA extension^13^.

### Search strategy

In consultation with an experienced research librarian, we searched MEDLINE, EMBASE, CENTRAL, and clinicaltrials.gov from inception to May 31, 2022, for randomized trials addressing biologic therapies for asthma. We did not restrict the search by language or date of publication. We supplemented our search by reviewing references of similar systematic reviews^7 8 14-16^. eTable 1 presents our full search strategy.

### Screening

Following training and calibration exercises to ensure sufficient agreement, pairs of reviewers, working independently and in duplicate, reviewed titles and abstracts of search results and subsequently the full-texts of records deemed potentially eligible at the title and abstract screening stage.

We included trials that randomized adults with asthma to one or more biologic therapies with standard care, placebo, or other biologic therapies, regardless of language or date of publication. We included trials that recruited both children and adults if 80% or more patients were adults (>18 years). We excluded trials that reported on the effects of aerosolized or nebulized biologics or trials that included fewer than 10 participants in each arm.

Reviewers resolved discrepancies by discussion, or, when necessary, by adjudication with a third party.

### Data extraction

Following training and calibration exercises to ensure sufficient agreement, pairs of reviewers were assigned the same trials, and then they worked independently and in duplicate, to extract data from eligible studies. This was done to avoid collection error. We extracted data on trial characteristics (e.g., country of recruitment), patient characteristics (e.g., age, eosinophil levels), intervention characteristics (e.g., dose and duration), and outcomes. The coreASTHMA core outcome set—a multistakeholder core outcome set for asthma—informed our outcomes of interest^17^, which include exacerbations, asthma control as measured by the Asthma Control Questionnaire, lung function measured by prebronchodilator forced expiratory volume in 1 second (FEV1), hospitalizations, reduction in use of systemic corticosteroids by ≥50%, and adverse events leading to discontinuation.

We anticipated that the effects of biologics on exacerbations, asthma control, and lung function may differ according to baseline eosinophil count. To facilitate subgroup analyses, we extracted subgroup data based on baseline blood and sputum eosinophil count, when reported. We classified trials as reporting on eosinophilic asthma when 80% or more of patients had blood eosinophils of ≥300 eosinophils per μL in the past year, ≥150 eosinophils per μL in the trial screening period (to allow for random fluctuations in blood eosinophils), or sputum eosinophils ≥3%. These cut-offs were based on common classifications found in randomized trials and broadly fits the research and clinical definitions.

### Risk of bias assessments

Following training and calibration exercises to ensure sufficient agreement, pairs of reviewers, working independently and in duplicate, assessed the risk of bias of trials using a modification of the Cochrane-endorsed RoB 2.0 tool^18 19^. We rated each outcome as either at (1) low risk of bias, (2) probably low risk of bias, (3) probably high risk of bias or (4) high risk of bias, across the following domains: bias arising from the randomisation process; bias owing to departures from the intended intervention; bias from missing outcome data; bias in measurement of the outcome; bias in selection of the reported results. Trials were rated at high risk of bias overall if one or more domains were rated at probably high or high risk of bias. Reviewers resolved discrepancies by discussion and, when not possible, with adjudication by a third party. eTable 2 presents the modified risk of bias tool.

### Data analysis

We present the results of dichotomous outcomes as relative risks (RRs) and continuous outcomes as mean differences (MDs), with associated 95% confidence intervals (95% CIs). To facilitate interpretation, for dichotomous outcomes, we calculate absolute effects expressed as events per 1,000 patients, using the median risk in the placebo arms as the assumed baseline risk without biologics.

For each outcome, we performed frequentist random-effects network meta-analysis using the restricted maximum likelihood (REML) estimator. Although Bayesian methods are also available, a recent study that investigated the differences between the two approaches found there are seldom important differences in the results of Bayesian and frequentist approaches for network meta-analysis^20^. We categorized each biologic drug as a separate node in the network. We anticipated that the effects of biologic therapies on exacerbations, asthma control, and lung function may differ based on baseline eosinophil count. For these outcomes, we performed three analyses: one restricted to patients with high eosinophils, one restricted to patients with low eosinophils, and one including all patients.

To test our assumption of transitivity and to determine whether there were important differences between trial results and trial and patient characteristics, we performed meta-regressions with FEV1, age, sex, blood eosinophils (mean), baseline oral corticosteroid use, duration of asthma, duration of follow up, and year of publication as independent variables. Meta-regressions test whether one or more independent variables influence the outcome^21^.

We summarize heterogeneity using the I^2^ statistic. We considered heterogeneity ranging from 0%–40% as potentially unimportant, 30%–60% as moderate, 50%–90% as substantial and 75%–100% as critical ^21^. We used the node-splitting method to test for local incoherence (difference between direct and indirect evidence in closed loops) ^22^. For analyses with 10 or more trials, we assessed for publication bias using comparison-adjusted funnel plots and the Egger’s test^23^.

We performed meta-analyses using meta and netmeta packages in R v. 4.1.2 (Vienna, Austria) and we used the networkplot command in Stata v.17 (StataCorp) to generate network plots^24-26^.

### Evaluation of the certainty (quality) of evidence

Reviewers, working independently and in duplicate, assessed the certainty (quality) of evidence using the GRADE approach for network meta-analysis^27-29^. Reviewers resolved discrepancies by discussion or by adjudication by a third reviewer.

For each comparison and for each outcome, we rated the certainty of evidence as either high, moderate, low, or very low based on considerations of risk of bias (i.e., study limitations), inconsistency (i.e., heterogeneity in trial results), indirectness (i.e., differences between the questions addressed in trials and the question of interest), publication bias (i.e., the tendency for trials with statistically significant results or positive results to be published, published faster, or published in journals with higher visibility), intransitivity (i.e., differences in trial characteristics across the network), incoherence (i.e., difference between direct and indirect effects), and imprecision (i.e., random error). High certainty evidence indicates situations in which we are confident that the estimated effect represents the true effect, and low or very low certainty evidence indicates situations in which the estimated effect may be substantially different from the true effect.

We made judgements about imprecision using the minimally contextualized GRADE approach^29^. This approach does not consider statistical significance as the only indicator of whether an intervention is effective. An estimate may not be statistically significant but may still have evidence of moderate certainty for benefit or harm, depending on the width of the confidence intervals and whether they cross the thresholds of clinical significance. Conversely, an intervention may produce results that are statistically significant but that indicate no important benefit or harm (e.g., a 10 mL change in FEV1). A minimally contextualized approach considers whether confidence intervals include the minimally important difference (the smallest change in the outcome considered to be important to patients) and does not consider whether it includes both minimally important and large effects.

We sourced minimally important differences from the literature or by consensus of the authors. We considered a 20% reduction in risk of exacerbations ^30 31^, 0.5 reduction in the Asthma Control Questionnaire^31^, 100 mL reduction in FEV1^30 32^, 5% reduction in risk of hospital admissions, 20% increase in risk of reducing oral corticosteroids by ≥50%, and a 10% increase in risk of adverse events leading to discontinuation to be minimally important.

We report our results using guidance from the GRADE Working Group, which involves describing the effect of an intervention based on the certainty of evidence (i.e., high certainty evidence the drug is effective, moderate certainty evidence the drug is probably effective, low certainty evidence the drug may be effective and very low certainty evidence the effect of the drug is unclear) ^33^.

We assessed the credibility of meta-regressions using the ICEMAN tool^34^.

## Results

### Search results

Our search yielded 3,225 unique references. We identified a total of 58 eligible publications reporting on 64 unique trials (i.e., some publications reported on more than one trial) with 26,630 patients^10-12 35-88^. Four trials were included in the systematic review but did not report on outcomes to be included in the meta-analysis^38 55 64 69^. All trials were published in peer-reviewed journals in English. eFigure 1 presents additional details related to study selection.

### Trial and participant characteristics

Table 1 presents trial and participant characteristics. Nineteen trials (4,974) reported on omalizumab, eight (4,511 patients) on fevipiprant, seven (2,511 patients) on reslizumab, six (1,966 patients) on mepolizumab, four (2,312 patients) on tralokinumab, four (1,799 patients) on tezepelumab, three (1,090 patients) on dupilumab, eight (4,043 patients) on benralizumab, one (502 patients) on astegolimab, and one (296 patients) on itepekimab^10-12 35-88^. We included two additional studies that reported on subgroups for omalizumab and eosinophils^89 90^.

Trials typically recruited adult patients with moderate to severe asthma on medium to high dose inhaled corticosteroids and other controller medications and with a history of one or more exacerbations in the previous year that required a short course of prednisone or hospitalization. Only five trials reported on mild patients, three of which also included moderate patients. Patients were also generally required to be symptomatic, as assessed by the Asthma Control Questionnaire.

Fifteen trials (5,791 patients) recruited patients with high eosinophils (blood eosinophils of ≥300 eosinophils per μL in the past year, ≥150 eosinophils per μL in the trial screening period, or sputum eosinophils ≥3%) and 51 trials (22,432 patients) recruited patients with both low and high eosinophils. Among trials that recruited patients with both low and high eosinophils, 19 (13,592 patients) reported subgroup analyses based on eosinophil levels for one or more of our outcomes of interest.

### Out of the trials included in the pooled-analyses (60 trials), 55 (91.7%) included patients ranging from moderate-to-severe and five trials (8.3%) included patients with mild-to-moderate asthma^45 47 57 72 76^. Of these five trials, two reported on omalizumab, and one each for mepolizumab, benralizumab, and fevipiprant. Risk of bias

eTable 3 presents risk of bias assessments. We judged two trials to be at high risk of bias due to their open-label design and inadequate description of allocation concealment^40 83^. We judged all other outcomes to be at low risk of bias^10 11 35-39 41-82 84-87 10 11 35-87^. Pharmaceutical companies funded all trials.

### Exacerbations

Forty-two trials, including 20,964 patients, reported exacerbations^10-12 35-37 39 40 42 44 45 48-51 56 59-62 65 67 68 71 73-75 77 79 80 82-87^. Of these, thirty trials reported on 11,329 patients with high eosinophils^10-12 35 36 39 42 44 49-51 56 59 61 62 67 74 75 77 79 85-90 10-12 35 44 49 61 74 75 85 87^ and fifteen trials reported 5,137 patients with low eosinophils. Figure 1 presents the geometry of the networks and figure 2 presents the network forest plot.

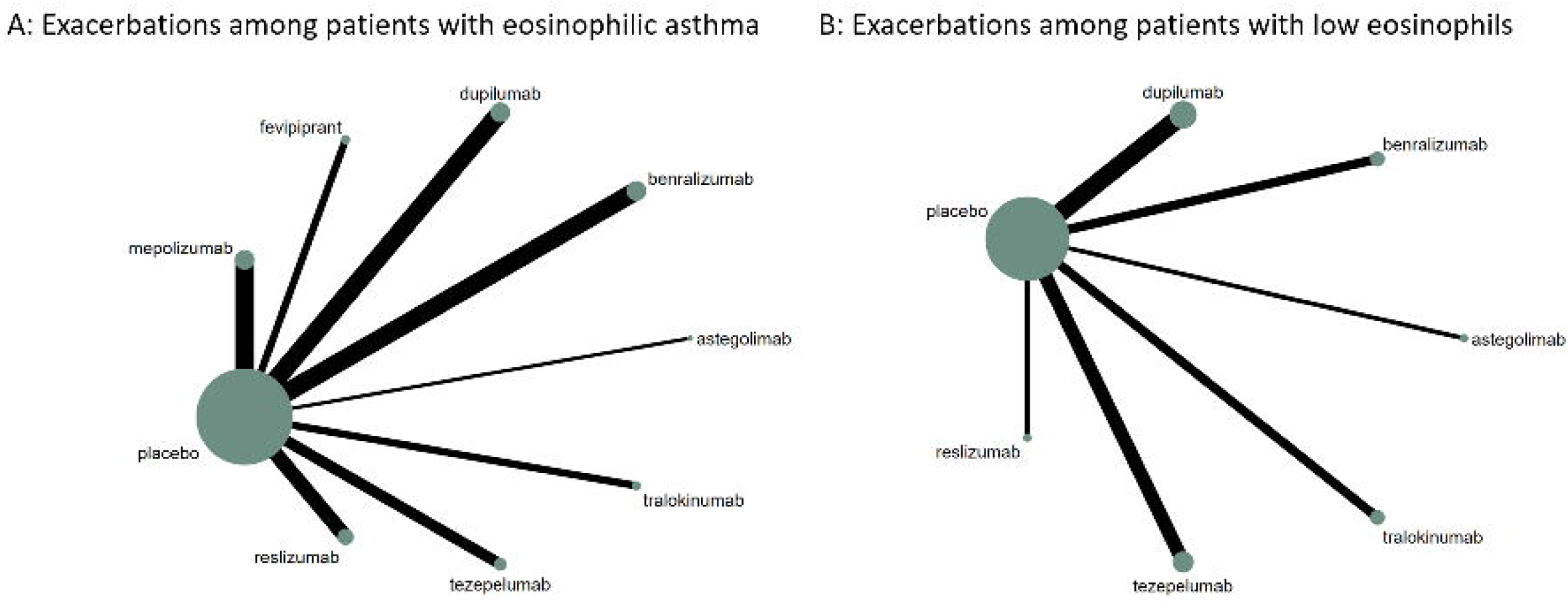

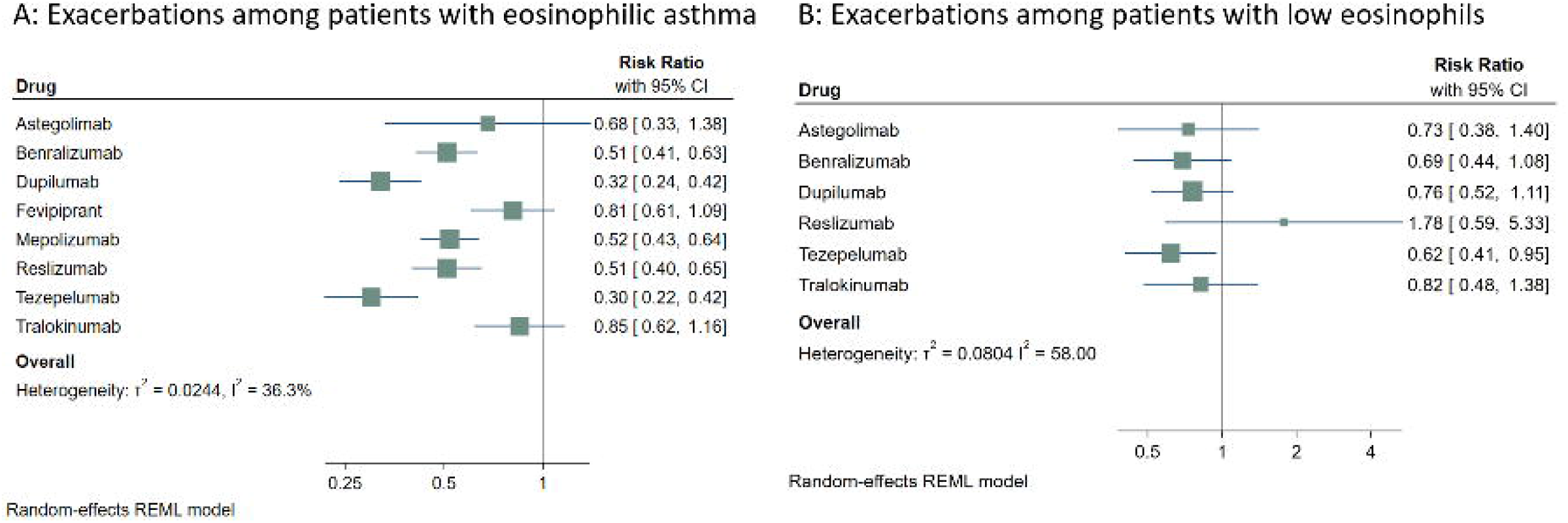

For patients with eosinophilic asthma (>=300 cells/uL), we found moderate to high certainty evidence that benralizumab (RR 0.51 [95% CI 0.41 to 0.63]), dupilumab (RR 0.32 [95% 0.24 to 0.42,]) mepolizumab (RR 0.52 [95% CI 0.43 to 0.64]), reslizumab (RR 0.51 [95% CI 0.4 to 0.65]), omalizumab (RR 0.52 [0.37 to 0.72]) and tezepelumab (RR 0.3 [0.22 to 0.43]) probably reduce exacerbations compared to placebo. While dupilumab and tezepelumab appear superior to the other biologics, the difference does not reach the minimally important difference.

For patients with low eosinophils (< 300 cells/uL), we did not find moderate or high certainty evidence that any of the drugs reduce exacerbations (i.e., none met the pre-specified minimally important difference of 20%).

For all patients, omalizumab, astegolimab, and tralokinumab probably do not reduce exacerbations as compared to placebo.

Table 2 presents the summary of findings and eTables 4-6 presents all comparisons. eFigure 2-13 present the forest plots, funnel plot and network diagrams.

eFigures 2-13 present the forest plots, funnel plot and network diagrams.

### Asthma control (Asthma Control Questionnaire)

Thirty-six trials, including 12,161 patients, reported on asthma control using the asthma control questionnaire^10-12 36 39 42 45 46 49-51 54 56-59 61 62 66 72 73 76-79 81 82 84-86 91 92^. Of these, ten trials reported on 8,212 patients with high eosinophils^10 11 76 77 79 81 85 86^ and eight trials reported on 1,905 patients with low eosinophils^10-12 49 54 61 76 85^

For patients with eosinophilic asthma, we found low certainty evidence that dupilumab may improve asthma control compared to placebo (MD -0.73 [95% CI -0.98 to -0.48]). We did not find moderate or high certainty evidence that any of the biologics improve asthma control for this population.

For patients with low eosinophils, we found moderate to high certainty evidence that benralizumab (MD -0.23 [95% CI -0.41 to -0.06]), dupilumab (MD -0.2 [95% -0.42 to 0.02]), reslizumab (MD 0.12 [95% CI - 0.09 to 0.33]), and tezepelumab (MD -0.23 [95% CI -0.36 to -0.09]) probably do not improve asthma control compared to placebo.

Table 2 presents the summary of findings and eTables 7-9 presents all comparisons. eFigure 14-25 present the forest plots, funnel plot and network diagrams.

### Lung function (FEV1)

Forty-two trials, including 17,965 patients, reported lung function^10 11 35 36 39 41 42 44-47 49-51 54 56 58 59 61-63 65 66 74 75 77-80 82-85 87^. Of these, twenty-nine trials reported on 9,242 patients with high eosinophils^59 62 66 77 79^ and twelve trials reported on 2745 patients with low eosinophils^10 11 58 61 75 85^.

For patients with high eosinophils, we found moderate to high certainty evidence that dupilumab (MD 0.25 L [95% CI 0.21 to 0.29]), reslizumab (MD 0.19 L [95% CI 0.12 to 0.25]), and tezepelumab (MD 0.24 L [0.16 to 0.32]) probably improve lung function compared to placebo. We found moderate certainty evidence that dupilumab is probably superior to benralizumab (MD 0.11 L [95% CI 0.05 to 0.16]) and mepolizumab (MD 0.15 L [95% CI 0.08 to 0.22]) for improving lung function and that tezepelumab is probably superior to mepolizumab (MD 0.15 L [95% CI 0.05 to 0.24]). We found moderate certainty evidence that dupilumab and tezepelumab probably improve lung function equally well (MD 0.01 L [95% CI -0.08 to 0.1]).

For patients with low eosinophils, we did not find moderate or high certainty evidence that any biologics improve lung function.

Table 2 presents the summary of findings and eTables 10-12 presents all comparisons. eFigures 26-37 present the forest plots, funnel plot and network diagrams.

### Hospital admissions

Thirteen trials, including 3,113 patients, reported on hospital admission^10 11 35 40 45 48 49 62 67 71 82 85^

We found moderate certainty evidence that mepolizumab (RR 0.29 [95% CI 0.09 to 0.97]), omalizumab (RR 0.38 [95% CI 0.23 to 0.65]), and tezepelumab (RR 0.19 [95% CI [0.12 to 0.31]) probably reduce hospital admissions compared to placebo.

We found moderate to high certainty evidence that omalizumab is probably superior to benralizumab (RR 0.44 [95% CI 0.25 to 0.78]) and tralokinumab (RR 0.5 [95% CI 0.27 to 0.95]) for reducing hospital admissions. We found moderate certainty evidence that tezepelumab is probably superior to benralizumab for reducing hospitalizations (RR 0.22 [95% CI 0.13 to 0.37]).

Table 2 presents the summary of findings and eTable 13 presents all comparisons. eFigures 38-41 present the forest plots, funnel plot and network diagram.

### Reduction in use of oral corticosteroids (≥50% reduction in oral corticosteroids)

Six trials, including 4849 patients, reported ≥50% reduction in oral corticosteroids^12 39 44 48 75 86^.

We found moderate certainty evidence that benralizumab (RR 1.77 [95% CI 1.29 to 2.43]), dupilumab (RR 1.49 [95% CI 1.22 to 1.83], and mepolizumab [RR 1.61 [95% CI 1.07 to 2.41]) probably reduce use of oral corticosteroids compared to placebo. We did not find moderate or high certainty evidence that either of these drugs are superior to each other.

Table 2 presents the summary of findings and eTable 14 presents all comparisons. eFigures 42-45 present the forest plots, funnel plot and network diagrams.

### Adverse events leading to discontinuation

Forty-seven trials, including 8,781 patients, reported adverse events leading to discontinuation^10-12 35-37 39-41 43-46 48-52 54 56 60-63 65 67 68 70-73 75-80 82-87^.

We have moderate to high certainty evidence that benralizumab (RR 1.65 [95% CI 0.79 to 3.45]), dupilumab (RR 1.03 [95% CI 0.46 to 2.3]), fevipiprant (RR 1.01 [95% CI 0.65 to 1.57]), mepolizumab [RR 0.65 [95% CI 0.36 to 1.16]), omalizumab (RR 1.2 [95% CI 0.80 to 1.81]), reslizumab (RR 0.65 [0.41 to 1.02]), tezepelumab (RR 0.68 [95% CI 0.0.34 to 1.35]), and tralokinumab (RR 2.7 [95% CI 1.45 to 5]) probably do not increase the risk adverse events compared to placebo.

Table 2 presents the summary of findings and eTable 15 presents all comparisons. eFigures 46-49 present the forest plots, funnel plot and network diagrams.

### Meta-regressions and sensitivity analyses

We did not find credible effect modification based on year of the trial publication, age, sex, duration of asthma, duration of follow-up, year of publication, and disease severity (assessed based on baseline FEV1 and oral corticosteroid use). eTable 16 presents the results of this analysis.

When excluding mild-to-moderate trials from the pooled analyses, there was no difference in the overall effect on any outcomes. eFigure 50-58 presents the forest plots for the sensitivity analyses.

## Discussion

### Main findings

Our systematic review and network meta-analysis presents data from 63 trials addressing the effectiveness and safety of biologics for uncontrolled severe asthma. We show that for patients with eosinophilic asthma, tezepelumab and dupilumab are the most effective at reducing asthma exacerbations and improving lung function. Furthermore, we found that tezepelumab and dupiliumab are probably better at improving lung function than other available biologic treatments. We did not find moderate or high certainty evidence that either of these drugs are superior to each other.

Contrary to expectations, for patients with low eosinophils, biologics, including tezepelumab that has been marketed as effective even in patients with low eosinophils^93^, probably do not prevent exacerbations or improve asthma control or lung function.

While our findings may appear to deviate from previously published trials, this is not surprising^10-12^. In interpreting the results, we use latest GRADE guidance and consider both the magnitude of effect and the certainty (quality) of evidence. We also consider effects in relation to the minimally important difference for each outcome, while trials generally interpret results based on statistical significance. Results from network meta-analyses may also be less precise than those reported in trials since the network estimate incorporates network wide heterogeneity^21^.

Considering these factors, while our findings may seem incongruent with the interpretation of previous trials, they are not incongruent with previously published trial results. The SOURCE trial, for example, did not find evidence that tezepelumab reduces oral corticosteroids or exacerbations in patients with low eosinophils and the effect of tezepelumab on lung function in patients with high eosinophils was twice that of patients with low eosinophils^12^. Similarly, NAVIGATOR found the effect of tezepelumab on lung function to be much greater in patients with high eosinophils than low eosinophils^11^.

### Strengths and limitation

The strengths of this review include our comprehensive search for trials addressing biologics for asthma, duplicate screening and data extraction, and rigorous evaluation of the certainty (quality) of evidence. We considered the potential differential effects of biologics based on eosinophil counts, which play a key role in the pathogenesis of severe asthma. We focused on patient-important outcomes and our choice of outcomes was guided by a core outcome set developed by stakeholders including patients^17^. We apply rigorous methods for assessing the certainty of evidence from network meta-analyses using a minimally contextualized approach and minimally important differences^29^. Furthermore, we considered the comparability of trials by performing meta-regression, demonstrating that there was no appreciable difference that would obviate the assumption of transitivity.

Although we performed a rigorous search for eligible studies and supplemented our search with previous systematic reviews of biologics for asthma, it is possible that we missed eligible trials.

While the GRADE framework presents a transparent system for assessing the certainty of evidence and accounts for all factors that may bear on the certainty of evidence, its application is subjective and others rating the certainty of evidence may come to different conclusions. Further, our assessment of the certainty of evidence using the minimally contextualized approach required us to identify the minimally important difference. We encourage evidence users to consider effects that their patients would consider to be important when interpreting our results. For example, some patients may place a high value on reductions in exacerbations and may consider a reduction in exacerbations less than 20% to still be important.

In our network meta-analysis, we categorized different doses of the same biologic and different routes of administration (i.e., subcutaneous vs. intravenous) in the same node. While this approach maximized the number of patients included in each node, the effects of biologics may vary by dose and route of administration. Most trials, however, that tested different doses and routes of administration did not find evidence of clinically important differences in effects ^10 61^.

Clinicians may be concerned that the included trials were not sufficiently similar to justify pooling. Although we detected moderate-to-substantial level network heterogeneity in some of the outcomes, we took this into account when providing GRADE ratings. For most part, the degree of heterogeneity did not warrant rating down the certainty and differences in the studies did not have appreciable effect on the results. The overwhelming majority of studies reported on moderate to severe asthma and only a few trials reported on patients with mild asthma. We performed meta-regressions testing for potential effect modification based on trial characteristics (e.g., year of publication) and patient characteristics (e.g., severity, duration of asthma) and did not find evidence that the effects of biologics varied based on these factors. Although there was a statistically significant effect with age and sex in the dupiliumab trials, this was not seen in other comparisons and likely resulting from chance. Using the ICEMAN tool, we found the credibility of this subgroup to be very low.

Trials reported data on the effects of biologics between 12 and 52 weeks. The effects of biologics, including potential adverse events, beyond these timeframes are uncertain.

We detected possible evidence of publication bias in overall networks for exacerbations, ACQ and FEV1, but not in the subgroups. It is possible we were underpowered to detect publication bias in the subgroup networks.

We report data on adverse events that led to discontinuation. While we found biologics to be well tolerated, serious and life-threatening adverse events are possible and were not captured by our review. A very small proportion of patients, for example, may experience anaphylaxis—typically after the first three doses.

We present convincing evidence that the effects of biologics vary based on eosinophils. We show that biologics, including tezepelumab, are probably not effective for patients with low eosinophils—a finding that is consistent with a previous review and previously published trials, although not explicitly emphasized in the review or trials^11 12 16^. Patients’ eosinophils also inherently vary over time. It is possible that trials may capture patients at a random high or low in eosinophil count as many trials recruiting patients with raised blood eosinophils usually obtain a single time point result of greater than 150-300 cells/uL. Furthermore, trials varied in their cut offs for “high eosinophils”, while most reported >=300 cells/uL, some trials included thresholds higher than 500 cells/uL. Therefore, our review may underestimate differences in effects of biologics based on eosinophils.

Our review focuses on adults and includes limited data on the effects of biologics in children. Given the effectiveness of these drugs in adults with eosinophilic asthma, additional clinical trials in children are warranted. Currently, omalizumab—which we found to not be effective—is the only biologic approved by the FDA for children under 6 years^94^.

### Implications

Our results have implications for the management of patients with severe uncontrolled asthma. Our results support the use of biologics for the treatment of patients with high eosinophils. For patients with eosinophilic asthma, tezepelumab and dupilumab are effective at reducing exacerbations, without important differences between the two biologics.

Our results, however, suggest that clinicians should be more cautious using biologics for patients with low eosinophils. In December 2021, the FDA approved tezepelumab for patients with severe uncontrolled asthma, regardless of eosinophil levels^93^. Tezepelumab is regarded as the first biologic to reduce asthma exacerbations consistently and significantly in a broad population of asthma patients. For patients with low eosinophils, tezepelumab, however, did now show evidence of effectiveness. Other investigators have also been skeptical of the effects of tezepelumab in patients with low eosinophils^95^.

Suboptimal response in patients with low eosinophils may be related to several pathways leading to asthma, including non-eosinophilic inflammation, airflow obstruction related to airway remodeling, significant airways hyperresponsiveness and/or mucus plugging being driven by non-eosinophil derived cytokines.

Our results suggest that clinicians should be mindful when using tezepelumab in patients with low eosinophils given the costs and practical issues associated with the drug (i.e., subcutaneous injection) and since these patients are unlikely to derive as substantial of a benefit as patients with eosinophilic asthma. However, for patients who have moderate-to-severe non-eosinophilic asthma, there are currently few options. Clinicians may consider a short trial of anti-alarmin therapy in patients who may benefit and evaluate for response in select patients.

Our results also do not support the use of omalizumab, which was markedly less effective than other biologics.

## Conclusion

For patients with eosinophilic asthma, all available biologics are effective at reducing exacerbations, without important differences in the effects of these biologics. Tezepelumab and dupiliumab probably improve lung function as compared to other available treatments. For patients with low eosinophils, however, clinicians should probably be more judicious in use of biologics, including tezepelumab, since they probably do not confer substantial benefit.

## Supporting information

esupplement

## Data Availability

On final publication of the article.

## Acknowledgements

None

## References

1. Global Asthma Network. Global Asthma Network Global Asthma Report 2018 https://www.globalasthmareport.org/ Date last updated: January 2018. Date last accessed: January 17, 2020.

2. Global Initiative for Asthma. (2002). Global strategy for asthma management and prevention. Bethesda, Md.: National Institutes of Health, National Heart, Lung, and Blood Institute.

3. Mauer Y, Taliercio RM. Managing adult asthma: The 2019 GINA guidelines. Cleve Clin J Med 2020;87(9):569–75. doi: 10.3949/ccjm.87a.19136 [published Online First: 20200831]

4. Porsbjerg CM, Sverrild A, Lloyd CM, Menzies-Gow AN, Bel EH. Anti-alarmins in asthma: targeting the airway epithelium with next-generation biologics. European Respiratory Journal 2020;56(5)

5. Sadeghirad B, Foroutan F, Zoratti MJ, Busse JW, Brignardello-Petersen R, Guyatt G, et al. Theory and practice of Bayesian and frequentist frameworks for network meta-analysis. BMJ Evid Based Med 2022 doi: 10.1136/bmjebm-2022-111928 [published Online First: 20220627]

6. Ando K, Fukuda Y, Tanaka A, Sagara H. Comparative Efficacy and Safety of Tezepelumab and Other Biologics in Patients with Inadequately Controlled Asthma According to Thresholds of Type 2 Inflammatory Biomarkers: A Systematic Review and Network Meta-Analysis. Cells 2022;11(5) doi: 10.3390/cells11050819 [published Online First: 20220226]

7. Edris A, Lahousse L. Monoclonal antibodies in type 2 asthma: an updated network meta-analysis. Minerva Med 2021;112(5):573–81. doi: 10.23736/s0026-4806.21.07623-0 [published Online First: 20210514]

8. Edris A, De Feyter S, Maes T, Joos G, Lahousse L. Monoclonal antibodies in type 2 asthma: a systematic review and network meta-analysis. Respir Res 2019;20(1):179. doi: 10.1186/s12931-019-1138-3 [published Online First: 20190808]

9. Ramonell RP, Iftikhar IH. Effect of Anti-IL5, Anti-IL5R, Anti-IL13 Therapy on Asthma Exacerbations: A Network Meta-analysis. Lung 2020;198(1):95–103. doi: 10.1007/s00408-019-00310-8 [published Online First: 20200101]

10. Corren J, Parnes JR, Wang L, Mo M, Roseti SL, Griffiths JM, et al. Tezepelumab in Adults with Uncontrolled Asthma. N Engl J Med 2017;377(10):936–46. doi: 10.1056/NEJMoa1704064

11. Menzies-Gow A, Corren J, Bourdin A, Chupp G, Israel E, Wechsler ME, et al. Tezepelumab in Adults and Adolescents with Severe, Uncontrolled Asthma. N Engl J Med 2021;384(19):1800. doi: https://doi.org/10.1056/NEJMoa2034975

12. Wechsler ME, Menzies-Gow A, Brightling CE, Kuna P, Korn S, Welte T, et al. Evaluation of the oral corticosteroid-sparing effect of tezepelumab in adults with oral corticosteroid-dependent asthma (SOURCE): a randomised, placebo-controlled, phase 3 study. Lancet respiratory medicine 2022 doi: https://doi.org/10.1016/S2213-2600(21)00537-3

13. Hutton B, Catalá-López F, Moher D. [The PRISMA statement extension for systematic reviews incorporating network meta-analysis: PRISMA-NMA]. Med Clin (Barc) 2016;147(6):262–6. doi: 10.1016/j.medcli.2016.02.025 [published Online First: 20160331]

14. Normansell R, Walker S, Milan SJ, Walters EH, Nair P. Omalizumab for asthma in adults and children. Cochrane Database Syst Rev 2014(1):Cd003559. doi: 10.1002/14651858.CD003559.pub4 [published Online First: 20140113]

15. Farne HA, Wilson A, Powell C, Bax L, Milan SJ. Anti-IL5 therapies for asthma. Cochrane Database Syst Rev 2017;9(9):Cd010834. doi: 10.1002/14651858.CD010834.pub3 [published Online First: 20170921]

16. Menzies-Gow A, Steenkamp J, Singh S, Erhardt W, Rowell J, Rane P, et al. Tezepelumab compared with other biologics for the treatment of severe asthma: a systematic review and indirect treatment comparison. J Med Econ 2022;25(1):679–90. doi: 10.1080/13696998.2022.2074195

17. tejwani V, Chang HY, Tran AP, Naber JA, Gutzwiller FS, Winders TA, et al. A multistakeholder Delphi consensus core outcome set for clinical trials in moderate-to-severe asthma (coreASTHMA). Ann Allergy Asthma Immunol 2021;127(1):116-22.e7. doi: 10.1016/j.anai.2021.03.022 [published Online First: 20210327]

18. Sterne JAC, Savović J, Page MJ, Elbers RG, Blencowe NS, Boutron I, et al. RoB 2: a revised tool for assessing risk of bias in randomised trials. Bmj 2019;366:n4898. doi: 10.1136/bmj.l4898 [published Online First: 20190828]

19. Pitre T, Van Alstine R, Chick G, Leung G, Mikhail D, Cusano E, et al. Antiviral drug treatment for nonsevere COVID-19: a systematic review and network meta-analysis. Cmaj 2022;194(28):E969–e80. doi: 10.1503/cmaj.220471

20. Sadeghirad B, Foroutan F, Zoratti MJ, Busse JW, Brignardello-Petersen R, Guyatt G, et al. Theory and practice of Bayesian and frequentist frameworks for network meta-analysis. BMJ Evidence-Based Medicine 2022:bmjebm-2022-111928. doi: 10.1136/bmjebm-2022-111928

21. Cumpston M, Li T, Page MJ, Chandler J, Welch VA, Higgins JP, et al. Updated guidance for trusted systematic reviews: a new edition of the Cochrane Handbook for Systematic Reviews of Interventions. Cochrane Database Syst Rev 2019;10:Ed000142. doi: 10.1002/14651858.Ed000142

22. van Valkenhoef G, Dias S, Ades AE, Welton NJ. Automated generation of node-splitting models for assessment of inconsistency in network meta-analysis. Res Synth Methods 2016;7(1):80–93. doi: 10.1002/jrsm.1167 [published Online First: 20151013]

23. Chaimani A, Salanti G. Using network meta-analysis to evaluate the existence of small-study effects in a network of interventions. Res Synth Methods 2012;3(2):161–76. doi: 10.1002/jrsm.57 [published Online First: 20120601]

24. Rücker G, Schwarzer G, Krahn U, König J, Schwarzer MG. Package ‘netmeta’. Network Meta-Analysis using Frequentist Methods (Version 07-0) 2015

25. Schwarzer G. meta: An R package for meta-analysis. R news 2007;7(3):40–45.

26. Chaimani A, Higgins JP, Mavridis D, Spyridonos P, Salanti G. Graphical tools for network meta-analysis in STATA. PloS one 2013;8(10):e76654.

27. Brignardello-Petersen R, Bonner A, Alexander PE, Siemieniuk RA, Furukawa TA, Rochwerg B, et al. Advances in the GRADE approach to rate the certainty in estimates from a network meta-analysis. J Clin Epidemiol 2018;93:36–44. doi: 10.1016/j.jclinepi.2017.10.005 [published Online First: 20171017]

28. Guyatt GH, Oxman AD, Vist GE, Kunz R, Falck-Ytter Y, Alonso-Coello P, et al. GRADE: an emerging consensus on rating quality of evidence and strength of recommendations. Bmj 2008;336(7650):924–6. doi: 10.1136/bmj.39489.470347.AD

29. Brignardello-Petersen R, Florez ID, Izcovich A, Santesso N, Hazlewood G, Alhazanni W, et al. GRADE approach to drawing conclusions from a network meta-analysis using a minimally contextualised framework. Bmj 2020;371:m3900. doi: 10.1136/bmj.m3900 [published Online First: 20201111]

30. Rogliani P, Calzetta L. Clinical Interpretation of Efficacy Outcomes in Pharmacological Studies on Triple Fixed-Dose Combination Therapy for Uncontrolled Asthma: Assessment of IRIDIUM and ARGON Studies. J Exp Pharmacol 2022;14:1–5. doi: 10.2147/jep.S336304 [published Online First: 20220111]

31. Bonini M, Di Paolo M, Bagnasco D, Baiardini I, Braido F, Caminati M, et al. Minimal clinically important difference for asthma endpoints: an expert consensus report. Eur Respir Rev 2020;29(156) doi: 10.1183/16000617.0137-2019 [published Online First: 20200603]

32. tepper RS, Wise RS, Covar R, Irvin CG, Kercsmar CM, Kraft M, et al. Asthma outcomes: pulmonary physiology. J Allergy Clin Immunol 2012;129(3 Suppl):S65–87. doi: 10.1016/j.jaci.2011.12.986

33. Santesso N, Glenton C, Dahm P, Garner P, Akl EA, Alper B, et al. GRADE guidelines 26: informative statements to communicate the findings of systematic reviews of interventions. J Clin Epidemiol 2020;119:126–35. doi: 10.1016/j.jclinepi.2019.10.014 [published Online First: 20191109]

34. Schandelmaier S, Briel M, Varadhan R, Schmid CH, Devasenapathy N, Hayward RA, et al. Development of the Instrument to assess the Credibility of Effect Modification Analyses (ICEMAN) in randomized controlled trials and meta-analyses. Cmaj 2020;192(32):E901–e06. doi: 10.1503/cmaj.200077

35. Panettieri RA, Sjobring U, Peterffy A, Wessman P, Bowen K, Piper E, et al. Tralokinumab for severe, uncontrolled asthma (STRATOS 1 and STRATOS 2): two randomised, double-blind, placebo-controlled, phase 3 clinical trials. Lancet respiratory medicine 2018;6(7):511. doi: https://doi.org/10.1016/S2213-2600(18)30184-X

36. Brightling CE, Gaga M, Inoue H, Li J, Maspero J, Wenzel S, et al. Effectiveness of fevipiprant in reducing exacerbations in patients with severe asthma (LUSTER-1 and LUSTER-2): two phase 3 randomised controlled trials. Lancet respiratory medicine 2021;9(1):43. doi: https://doi.org/10.1016/S2213-2600(20)30412-4

37. Vignola AM, Humbert M, Bousquet J, Boulet LP, Hedgecock S, Blogg M, et al. Efficacy and tolerability of anti-immunoglobulin E therapy with omalizumab in patients with concomitant allergic asthma and persistent allergic rhinitis: SOLAR. Allergy 2004;59(7):709. doi: https://doi.org/10.1111/j.1398-9995.2004.00550.x

38. Gevaert P, Bachert C, Maspero JF, Cuevas M, Steele D, Acharya S, et al. Phase 3b randomized controlled trial of fevipiprant in patients with nasal polyposis with asthma (THUNDER). Journal of Allergy and Clinical Immunology 2022 doi: https://doi.org/10.1016/j.jaci.2021.12.759

39. Nair P, Wenzel S, Rabe KF, Bourdin A, Lugogo NL, Kuna P, et al. Oral Glucocorticoid-Sparing Effect of Benralizumab in Severe Asthma. New England journal of medicine 2017;376(25):2448. doi: https://doi.org/10.1056/NEJMoa1703501

40. Ayres JG, Higgins B, Chilvers ER, Ayre G, Blogg M, Fox H. Efficacy and tolerability of anti-immunoglobulin E therapy with omalizumab in patients with poorly controlled (moderate-to-severe) allergic asthma. Allergy 2004;59(7):701. doi: https://doi.org/10.1111/j.1398-9995.2004.00533.x

41. Bardelas J, Figliomeni M, Kianifard F, Meng X. A 26-week, randomized, double-blind, placebo-controlled, multicenter study to evaluate the effect of omalizumab on asthma control in patients with persistent allergic asthma. Journal of asthma 2012;49(2):144. doi: https://doi.org/10.3109/02770903.2011.648296

42. Harrison TW, Chanez P, Menzella F, Canonica GW, Louis R, Cosio BG, et al. Onset of effect and impact on health-related quality of life, exacerbation rate, lung function, and nasal polyposis symptoms for patients with severe eosinophilic asthma treated with benralizumab (ANDHI): a randomised, controlled, phase 3b trial. Lancet respiratory medicine 2021;9(3):260. doi: https://doi.org/10.1016/S2213-2600(20)30414-8

43. Bateman ED, Guerreros AG, Brockhaus F, Holzhauer B, Pethe A, Kay RA, et al. Fevipiprant, an oral prostaglandin DP2 receptor (CRTh2) antagonist, in allergic asthma uncontrolled on low-dose inhaled corticosteroids. The european respiratory journal 2017;50(2) doi: https://doi.org/10.1183/13993003.00670-2017

44. Bernstein JA, Virchow JC, Murphy K, Maspero JF, Jacobs J, Adir Y, et al. Effect of fixed-dose subcutaneous reslizumab on asthma exacerbations in patients with severe uncontrolled asthma and corticosteroid sparing in patients with oral corticosteroid-dependent asthma: results from two phase 3, randomised, double-blind, placebo-controlled trials. Lancet respiratory medicine 2020;8(5):461. doi: https://doi.org/10.1016/S2213-2600(19)30372-8

45. Ferguson GT, FitzGerald JM, Bleecker ER, Laviolette M, Bernstein D, LaForce C, et al. Benralizumab for patients with mild to moderate, persistent asthma (BISE): a randomised, double-blind, placebo-controlled, phase 3 trial. Lancet respiratory medicine 2017;5(7):568. doi: https://doi.org/10.1016/S2213-2600(17)30190-X

46. Bjermer L, Lemiere C, Maspero J, Weiss S, Zangrilli J, Germinaro M. Reslizumab for Inadequately Controlled Asthma With Elevated Blood Eosinophil Levels: a Randomized Phase 3 Study. Chest 2016;150(4):789. doi: https://doi.org/10.1016/j.chest.2016.03.032

47. Boulet LP, Chapman KR, ocirc, eacute, Kalra S, Bhagat R, et al. Inhibitory effects of an anti-IgE antibody E25 on allergen-induced early asthmatic response. American journal of respiratory and critical care medicine 1997;155(6):1835. doi: https://doi.org/10.1164/ajrccm.155.6.9196083

48. Busse WW. Anti-immunoglobulin E (omalizumab) therapy in allergic asthma. American Journal of Respiratory and Critical Care Medicine 2001;164(8 II):S12–S17. doi: http://dx.doi.org/10.1164/ajrccm.164.supplement_1.2103026

49. FitzGerald JM, Bleecker ER, Nair P, Korn S, Ohta K, Lommatzsch M, et al. Benralizumab, an anti-interleukin-5 receptor &agr; monoclonal antibody, as add-on treatment for patients with severe, uncontrolled, eosinophilic asthma (CALIMA): a randomised, double-blind, placebo-controlled phase 3 trial. Lancet 2016;388(10056):2128. doi: https://doi.org/10.1016/S0140-6736(16)31322-8

50. Castro M, Zangrilli J, Wechsler ME, Bateman ED, Brusselle GG, Bardin P, et al. Reslizumab for inadequately controlled asthma with elevated blood eosinophil counts: results from two multicentre, parallel, double-blind, randomised, placebo-controlled, phase 3 trials. Lancet respiratory medicine 2015;3(5):355. doi: https://doi.org/10.1016/S2213-2600(15)00042-9

51. Castro M, Mathur S, Hargreave F, Boulet LP, Xie F, Young J, et al. Reslizumab for poorly controlled, eosinophilic asthma: a randomized, placebo-controlled study. American journal of respiratory and critical care medicine 2011;184(10):1125. doi: https://doi.org/10.1164/rccm.201103-0396OC

52. Chanez P, Contin-Bordes C, Garcia G, Verkindre C, Didier A, De Blay F, et al. Omalizumab-induced decrease of FcepsilonRI expression in patients with severe allergic asthma. Respiratory Medicine 2010;104(11):1608–17. doi: http://dx.doi.org/10.1016/j.rmed.2010.07.011

53. Corren J, Wood RA, Patel D, Zhu J, Yegin A, Dhillon G, et al. Effects of omalizumab on changes in pulmonary function induced by controlled cat room challenge. Journal of allergy and clinical immunology 2011;127(2):398. doi: https://doi.org/10.1016/j.jaci.2010.09.043

54. Corren J, Weinstein S, Janka L, Zangrilli J, Garin M. Phase 3 Study of Reslizumab in Patients With Poorly Controlled Asthma: effects Across a Broad Range of Eosinophil Counts. Chest 2016;150(4):799. doi: https://doi.org/10.1016/j.chest.2016.03.018

55. Djukanovic R, Wilson SJ, Kraft M, Jarjour N, Steel M, Chung KF, et al. Effect of treatment with anti-IgE antibody (Omalizumab) on airway inflammation in mild atopic asthma. American thoracic society 99th international conference 2003:C082.

56. Pavord ID, Korn S, Howarth P, Bleecker ER, Buhl R, Keene ON, et al. Mepolizumab for severe eosinophilic asthma (DREAM): a multicentre, double-blind, placebo-controlled trial. Lancet (london, england) 2012;380(9842):651. doi: https://doi.org/10.1016/S0140-6736(12)60988-X

57. Erpenbeck VJ, Popov TA, Miller D, Weinstein SF, Spector S, Magnusson B, et al. The oral CRTh2 antagonist QAW039 (fevipiprant): a phase II study in uncontrolled allergic asthma. Pulmonary pharmacology & therapeutics 2016;39:54. doi: https://doi.org/10.1016/j.pupt.2016.06.005

58. Wechsler ME, Ruddy MK, Pavord ID, Israel E, Rabe KF, Ford LB, et al. Efficacy and Safety of Itepekimab in Patients with Moderate-to-Severe Asthma. New England journal of medicine 2021;385(18):1656. doi: https://doi.org/10.1056/NEJMoa2024257

59. Panettieri RA, Welte T, Shenoy KV, Korn S, Jandl M, Kerwin EM, et al. Onset of effect, changes in airflow obstruction and lung volume, and health-related quality of life improvements with benralizumab for patients with severe eosinophilic asthma: phase iiib randomized, controlled trial (SOLANA). Journal of asthma and allergy 2020;13:115. doi: https://doi.org/10.2147/JAA.S240044

60. Sol, egrave r M, Matz J, Townley R, Buhl R, et al. The anti-IgE antibody omalizumab reduces exacerbations and steroid requirement in allergic asthmatics. The european respiratory journal 2001;18(2):254. doi: https://doi.org/10.1183/09031936.01.00092101

61. Wenzel S, Castro M, Corren J, Maspero J, Wang L, Zhang B, et al. Dupilumab efficacy and safety in adults with uncontrolled persistent asthma despite use of medium-to-high-dose inhaled corticosteroids plus a long-acting β2 agonist: a randomised double-blind placebo-controlled pivotal phase 2b dose-ranging trial. Lancet 2016;388(10039):31–44. doi: 10.1016/s0140-6736(16)30307-5 [published Online First: 20160427]

62. Wenzel S, Ford L, Pearlman D, Spector S, Sher L, Skobieranda F, et al. Dupilumab in persistent asthma with elevated eosinophil levels. New England journal of medicine 2013;368(26):2455. doi: https://doi.org/10.1056/NEJMoa1304048

63. Castro M, Kerwin E, Miller D, Pedinoff A, Sher L, Cardenas P, et al. Efficacy and safety of fevipiprant in patients with uncontrolled asthma: Two replicate, phase 3, randomised, double-blind, placebo-controlled trials (ZEAL-1 and ZEAL-2). EClinicalMedicine 2021;35 doi: https://doi.org/10.1016/j.eclinm.2021.100847

64. Zielen S, Lieb A, De La Motte S, Wagner F, De Monchy J, Fuhr R, et al. Omalizumab protects against allergen-induced bronchoconstriction in allergic (Immunoglobulin E-mediated) Asthma. International archives of allergy and immunology 2013;160(1):102. doi: https://doi.org/10.1159/000339243

65. Flood-Page P, Swenson C, Faiferman I, Matthews J, Williams M, Brannick L, et al. A study to evaluate safety and efficacy of mepolizumab in patients with moderate persistent asthma. American journal of respiratory and critical care medicine 2007;176(11):1062. doi: https://doi.org/10.1164/rccm.200701-085OC

66. Gonem S, Berair R, Singapuri A, Hartley R, Laurencin MFM, Bacher G, et al. Fevipiprant, a prostaglandin D2 receptor 2 antagonist, in patients with persistent eosinophilic asthma: a single-centre, randomised, double-blind, parallel-group, placebo-controlled trial. Lancet respiratory medicine 2016;4(9):699. doi: https://doi.org/10.1016/S2213-2600(16)30179-5

67. Haldar P, Brightling CE, Hargadon B, Gupta S, Monteiro W, Sousa A, et al. Mepolizumab and exacerbations of refractory eosinophilic asthma. New England journal of medicine 2009;360(10):973. doi: https://doi.org/10.1056/NEJMoa0808991

68. Hanania NA, Alpan O, Hamilos DL, Condemi JJ, Reyes-Rivera I, Zhu J, et al. Omalizumab in severe allergic asthma inadequately controlled with standard therapy: a randomized trial. Annals of internal medicine 2011;154(9):573. doi: https://doi.org/10.7326/0003-4819-154-9-201105030-00002

69. Hayashi H, Fukutomi Y, Mitsui C, Kajiwara K, Watai K, Kamide Y, et al. Omalizumab for Aspirin Hypersensitivity and Leukotriene Overproduction in Aspirin-exacerbated Respiratory Disease. A Randomized Controlled Trial. American journal of respiratory and critical care medicine 2020;201(12):1488. doi: https://doi.org/10.1164/rccm.201906-1215OC

70. Holgate ST, Chuchalin AG, eacute, bert J ouml, tvall J, et al. Efficacy and safety of a recombinant anti-immunoglobulin E antibody (omalizumab) in severe allergic asthma. Clinical and experimental allergy 2004;34(4):632. doi: https://doi.org/10.1111/j.1365-2222.2004.1916.x

71. Humbert M, Beasley R, Ayres J, Slavin R, eacute, bert J, et al. Benefits of omalizumab as add-on therapy in patients with severe persistent asthma who are inadequately controlled despite best available therapy (GINA 2002 step 4 treatment): INNOVATE. Allergy 2005;60(3):309. doi: https://doi.org/10.1111/j.1398-9995.2004.00772.x

72. Kopp MV, Hamelmann E, Zielen S, Kamin W, Bergmann KC, Sieder C, et al. Combination of omalizumab and specific immunotherapy is superior to immunotherapy in patients with seasonal allergic rhinoconjunctivitis and co-morbid seasonal allergic asthma. Clinical and experimental allergy 2009;39(2):271. doi: https://doi.org/10.1111/j.1365-2222.2008.03121.x

73. Li J, Kang J, Wang C, Yang J, Wang L, Kottakis I, et al. Omalizumab improves quality of life and asthma control in Chinese patients with moderate to severe asthma: a randomized phase III study. Allergy, asthma & immunology research 2016;8(4):319. doi: https://doi.org/10.4168/aair.2016.8.4.319

74. Castro M, Corren J, Pavord ID, Maspero J, Wenzel S, Rabe KF, et al. Dupilumab Efficacy and Safety in Moderate-to-Severe Uncontrolled Asthma. New England journal of medicine 2018;378(26):2486. doi: https://doi.org/10.1056/NEJMoa1804092

75. Rabe KF, Nair P, Brusselle G, Maspero JF, Castro M, Sher L, et al. Efficacy and Safety of Dupilumab in Glucocorticoid-Dependent Severe Asthma. New England journal of medicine 2018;378(26):2475. doi: https://doi.org/10.1056/NEJMoa1804093

76. Sabogal P ntilde, eros YS, Bal SM, van de Pol MA, Dierdorp BS, et al. Anti-IL-5 in Mild Asthma Alters Rhinovirus-induced Macrophage, B-Cell, and Neutrophil Responses (MATERIAL). A Placebo-controlled, Double-Blind Study. American journal of respiratory and critical care medicine 2019;199(4):508. doi: https://doi.org/10.1164/rccm.201803-0461OC

77. Ortega HG, Liu MC, Pavord ID, Brusselle GG, FitzGerald JM, Chetta A, et al. Mepolizumab treatment in patients with severe eosinophilic asthma. New England journal of medicine 2014;371(13):1198. doi: https://doi.org/10.1056/NEJMoa1403290

78. Russell RJ, Chachi L, FitzGerald JM, Backer V, Olivenstein R, Titlestad IL, et al. Effect of tralokinumab, an interleukin-13 neutralising monoclonal antibody, on eosinophilic airway inflammation in uncontrolled moderate-to-severe asthma (MESOS): a multicentre, double-blind, randomised, placebo-controlled phase 2 trial. Lancet respiratory medicine 2018;6(7):499. doi: https://doi.org/10.1016/S2213-2600(18)30201-7

79. Chupp GL, Bradford ES, Albers FC, Bratton DJ, Wang-Jairaj J, Nelsen LM, et al. Efficacy of mepolizumab add-on therapy on health-related quality of life and markers of asthma control in severe eosinophilic asthma (MUSCA): a randomised, double-blind, placebo-controlled, parallel-group, multicentre, phase 3b trial. Lancet respiratory medicine 2017;5(5):390. doi: https://doi.org/10.1016/S2213-2600(17)30125-X

80. Ohta K, Miyamoto T, Amagasaki T, Yamamoto M. Efficacy and safety of omalizumab in an Asian population with moderate-to-severe persistent asthma. Respirology 2009;14(8):1156–65. doi: http://dx.doi.org/10.1111/j.1440-1843.2009.01633.x

81. Park HS, Kim MK, Imai N, Nakanishi T, Adachi M, Ohta K, et al. A Phase 2a Study of Benralizumab for Patients with Eosinophilic Asthma in South Korea and Japan. Int Arch Allergy Immunol 2016;169(3):135–45. doi: 10.1159/000444799 [published Online First: 20160421]

82. Nowak RM, Parker JM, Silverman RA, Rowe BH, Smithline H, Khan F, et al. A randomized trial of benralizumab, an antiinterleukin 5 receptor alpha monoclonal antibody, after acute asthma. American journal of emergency medicine 2015;33(1):14. doi: https://doi.org/10.1016/j.ajem.2014.09.036

83. Rubin AS, Souza-Machado A, Andradre-Lima M, Ferreira F, Honda A, Matozo TM, et al. Effect of omalizumab as add-on therapy on asthma-related quality of life in severe allergic asthma: a Brazilian study (QUALITX). Journal of asthma 2012;49(3):288. doi: https://doi.org/10.3109/02770903.2012.660297

84. Piper E, Brightling C, Niven R, Oh C, Faggioni R, Poon K, et al. A phase II placebo-controlled study of tralokinumab in moderate-to-severe asthma. The european respiratory journal 2013;41(2):330. doi: https://doi.org/10.1183/09031936.00223411

85. Bleecker ER, FitzGerald JM, Chanez P, Papi A, Weinstein SF, Barker P, et al. Efficacy and safety of benralizumab for patients with severe asthma uncontrolled with high-dosage inhaled corticosteroids and long-acting &bgr;2-agonists (SIROCCO): a randomised, multicentre, placebo-controlled phase 3 trial. Lancet (london, england) 2016;388(10056):2115. doi: https://doi.org/10.1016/S0140-6736(16)31324-1

86. Bel EH, Wenzel SE, Thompson PJ, Prazma CM, Keene ON, Yancey SW, et al. Oral glucocorticoid-sparing effect of mepolizumab in eosinophilic asthma. New England journal of medicine 2014;371(13):1189. doi: https://doi.org/10.1056/NEJMoa1403291

87. Kelsen SG, Agache IO, Soong W, Israel E, Chupp GL, Cheung DS, et al. Astegolimab (anti-ST2) efficacy and safety in adults with severe asthma: a randomized clinical trial. Journal of allergy and clinical immunology 2021;148(3):790. doi: https://doi.org/10.1016/j.jaci.2021.03.044

88. Busse W, Spector S, Rosén K, Wang Y, Alpan O. High eosinophil count: a potential biomarker for assessing successful omalizumab treatment effects. J Allergy Clin Immunol 2013;132(2):485-6.e11. doi: 10.1016/j.jaci.2013.02.032 [published Online First: 20130413]

89. Casale TB, Chipps BE, Rosén K, Trzaskoma B, Haselkorn T, Omachi TA, et al. Response to omalizumab using patient enrichment criteria from trials of novel biologics in asthma. Allergy 2018;73(2):490–97. doi: 10.1111/all.13302 [published Online First: 20170923]

90. Hanania NA, Wenzel S, Rosén K, Hsieh HJ, Mosesova S, Choy DF, et al. Exploring the effects of omalizumab in allergic asthma: an analysis of biomarkers in the EXTRA study. Am J Respir Crit Care Med 2013;187(8):804–11. doi: 10.1164/rccm.201208-1414OC

91. Juniper EF, O’Byrne PM, Guyatt GH, Ferrie PJ, King DR. Development and validation of a questionnaire to measure asthma control. Eur Respir J 1999;14(4):902–7. doi: 10.1034/j.1399-3003.1999.14d29.x

92. Sverrild A, Hansen S, Hvidtfeldt M, Clausson CM, Cozzolino O, Cerps S, et al. The effect of tezepelumab on airway hyperresponsiveness to mannitol in asthma (UPSTREAM). European Respiratory Journal 2022;59(1):2101296. doi: https://dx.doi.org/10.1183/13993003.01296-2021

93. Tan LD, Nguyen N, Alismail A, Castro M. Management of Uncontrolled Asthma: A Framework for Novel and Legacy Biologic Treatments. J Asthma Allergy 2022;15:875–83. doi: 10.2147/jaa.S369836 [published Online First: 20220629]

94. Krings JG, McGregor MC, Bacharier LB, Castro M. Biologics for Severe Asthma: Treatment-Specific Effects Are Important in Choosing a Specific Agent. J Allergy Clin Immunol Pract 2019;7(5):1379–92. doi: 10.1016/j.jaip.2019.03.008

95. Adatia A, Wahab M, Satia I. Is tezepelumab more than just an anti-eosinophil drug? Eur Respir J 2022;59(1) doi: 10.1183/13993003.01700-2021 [published Online First: 20211231]

