## Supplementary material for "A comparison of the effectiveness of biologic therapies for asthma: a systematic review and network meta-analysis": esupplement

### eTable 1. Search strategy

| Randomized trials that have investigated the effectiveness of biologics for asthma (any type)  Biologics to include: Astegolimab, Benralizumab, Dupilumab, Fevipiprant, Itepekimab, Mepolizumab, Omalizumab, Reslizumab, Tezepelumab, Tralokinumab    **Search Disclaimer:**  Results of database searches are subject to limitations of the database(s) searched. It is the responsibility of the requestor to determine the accuracy, validity and interpretation of the search results.  **PRISMA Initial Results:** total number of results before duplicates removed |  |
| --- | --- |
| **Database [Platform]** Searches run May 31, 2022. *No date, language limits used.* | **Results** |
| Medline Epub Ahead of Print, In-Process & Other Non-Indexed Citations, Ovid MEDLINE(R) Daily and Ovid MEDLINE(R) 1946 to Present [Ovid] | 694 |
| Embase 1974 to 2022 May 27 [Ovid] | 2,342 |
| EBM Reviews - Cochrane Central Register of Controlled Trials April 2022 [Ovid] | 1,603 |
| **TOTAL** | **4,639** |

**ClinicalTrials.gov Results** Searched May 31, 2022. *No date, language limits used.*

| benralizumab OR dupilumab OR mepolizumab OR omalizumab OR reslizumab OR tezepelumab OR astegolimab OR itepekimab OR fevipiprant OR tralokinumab \| Completed, Suspended, Terminated, Withdrawn Studies \| Interventional Studies \| Asthma | 163 |
| --- | --- |
| **TOTAL** | **163** |

Medline Epub Ahead of Print, In-Process & Other Non-Indexed Citations, Ovid MEDLINE(R) Daily and Ovid MEDLINE(R) 1946 to Present

Search Strategy:

| **#** | **Searches** | **Results** |
| --- | --- | --- |
| 1 | asthma/ or asthma, aspirin-induced/ or asthma, exercise-induced/ or asthma, occupational/ or asthma-chronic obstructive pulmonary disease overlap syndrome/ or status asthmaticus/ | 137719 |
| 2 | (asthma or asthmas or asthmatic or exercise induced bronchospasm* or lung allergy or status asthmaticus).tw,kf. | 171783 |
| 3 | or/1-2 | 192649 |
| 4 | (astegolimab or "amg 282" or "amg282" or "mstt 1041a" or "mstt1041a" or "rg 6149" or "rg6149" or "ro 7187807" or "ro7187807" or "ro7187807").tw,kf. | 8 |
| 5 | (benralizumab or "biw 8405" or "biw8405" or "fasenra" or "medi 563" or "medi563").tw,kf. | 542 |
| 6 | (dupilumab or dupixent or "regn 668" or "regn668" or "sar 231893" or "sar231893").tw,kf. | 1577 |
| 7 | (fevipiprant or "qaw 039" or "qaw039").tw,kf. | 44 |
| 8 | (itepekimab or "regn 3500" or "regn3500" or "sar 440340" or "sar440340").tw,kf. | 5 |
| 9 | (mepolizumab or bosatria or nucala or "sb 240563" or "sb240563").tw,kf. | 1000 |
| 10 | Omalizumab/ | 2125 |
| 11 | (omalizumab or xolair or "fb 317" or "fb317" or "gbr 310" or "gbr310" or "hu 901" or "hu901" or "monoclonal antibody E 25" or "monoclonal antibody E25" or "rg 3648" or "rg3648" or "rhumab 25" or "rhumab e25" or "rhumab25" or "rhumabe25" or "sti 004" or "sti004" or "syn 008" or "syn008").tw,kf. | 2973 |
| 12 | (reslizumab or "cep38072" or "cep-38072" or "cinqaero" or "cinqair" or "dcp 835" or "dcp835" or "dcp-835" or "sch 55700" or "sch 55700" or "sch5500").tw,kf. | 318 |
| 13 | (tezepelumab or tezspire or "amg 157" or "amg157" or "medi 9929" or "medi9929" or "medi 19929" or "medi19929").tw,kf. | 84 |
| 14 | (tralokinumab or "cat 354" or "cat-354" or "lp 0162" or "lp0162").tw,kf. | 121 |
| 15 | or/4-14 | 5761 |
| 16 | 3 and 15 | 2799 |
| 17 | randomized controlled trial.pt. | 569197 |
| 18 | controlled clinical trial.pt. | 94883 |
| 19 | randomized.ab. | 562400 |
| 20 | placebo.ab. | 228466 |
| 21 | clinical trials as topic.sh. | 199944 |
| 22 | randomly.ab. | 383076 |
| 23 | trial.ti. | 263052 |
| 24 | 17 or 18 or 19 or 20 or 21 or 22 or 23 | 1448732 |
| 25 | animals/ not humans.sh. | 4977295 |
| 26 | 24 not 25 | 1332511 |
| 27 | 16 and 26 [Box 3.d Cochrane Highly Sensitive Search Strategy for identifying randomized trials in MEDLINE: sensitivity- and precision-maximizing version] | 694 |

### eTable 2. Modified risk of bias tool 2.0

| **Bias from the randomization process** | |
| --- | --- |
| Issues to consider:  Random sequence generation  Allocation concealment | |
| **Definitely low risk of bias** | Trials that assign participants to alternative interventions using a randomly generated sequence and maintain allocation concealment.  Examples of methods for developing a randomly generated allocation sequence include a random number generator, random number table, coin tossing, shuffling cards or envelopes, and throwing dice. If a trial is described as 'randomized' without any additional details related to how the allocation sequence was developed, we will assume that the allocation sequence was appropriately developed.  Examples of methods for maintaining allocation concealment include using central allocation via a computer or phone system, pharmacy-controlled allocation, opaque sealed envelopes, and sequentially numbered drug containers.  *Note that an explicit description of random sequence generation is not necessary for a rating of low risk of bias.* |
| **Probably low risk of bias** | Trials in which healthcare providers were blind to the intervention but which provide no information on allocation concealment.  *Note that an explicit description of random sequence generation is not necessary for a rating of probably low risk of bias.* |
| **Probably high risk of bias** | Trials in which healthcare providers were not blind to the intervention and which provide no information on allocation concealment.  Trials in which there are substantial baseline differences between trial arms that suggest a problem with the randomization process but there are no other limitations related to randomization. |
| **Definitely high risk of bias** | Trials in which allocation is by judgment of the clinician, by preference of the participant, by availability of the intervention, based on the results of a laboratory test, or other non-random rules (e.g., birthdate, etc.).  Trials in which investigators enrolling participants could possibly foresee the arm to which each subsequent patient would be randomized, such as allocation using an open allocation schedule (e.g. a list of random numbers), assignment envelopes used without appropriate safeguards (e.g. use of unsealed, non-opaque or not sequentially numbered envelopes), alternation between arms, case record number, or any other explicitly unconcealed procedure, rate as high risk. |
| **Bias due to deviations from the intended intervention** | |
| Issues to consider:  Blinding of healthcare providers/clinicians and participants  Imbalances in cointerventions or behaviors | |
| **Definitely low risk of bias** | Trials in which healthcare providers are blind to the intervention administered and in which there are no significant differences in administered co-interventions.  Trials that are described as double or triple blind. |
| **Probably low risk of bias** |  |
| **Probably high risk of bias** | Trials in which healthcare providers are not blind to the intervention administered.  Trials in which healthcare providers are blind to the intervention administered but there are significant differences in administered co-interventions that suggests that blinding may have been compromised.  Trials in which healthcare providers are described as being blind to the intervention but allocation concealment was inadequate. |
| **Definitely high risk of bias** | Trials in which healthcare providers are not blind to the intervention and in which there are significant differences in administered co-interventions. |
| **Bias due to missing data** | |
| Issues to consider:  Missing outcome measures  Loss to follow-up | |
| **Definitely low risk of bias** | Trials in which missing outcome data (including outcome data that has been imputed) < 10%. |
| **Probably low risk of bias** | Trials in which missing outcome data (including outcome data that has been imputed) is between 10% to 15% and missing outcome data is unlikely to be related to the true outcome and there is no imbalance in numbers of or reasons for missing data across intervention groups. |
| **Probably high risk of bias** | Trials in which missing outcome data (including outcome data that has been imputed) is between 10% to 15% and missing outcome data is likely to be related to the true outcome or there are imbalances in numbers of or reasons for missing data across intervention groups. |
| **Definitely high risk of bias** | Trials in which missing outcome data (including outcome data that has been imputed) > 15%. |
| **Bias due to measurement of the outcome** | |
| Issues to consider:  Blinding of outcome adjudicators  Objectivity of outcome  *Note that the judgments may differ across outcomes.* | |
| **Definitely low risk of bias** | Trials in which patients are blind to the intervention and in which outcomes are patient-reported.  Trials in which outcomes are measured by a third-party (investigator or clinician) and in which the third-party is blind to the intervention.  Trials in which the outcomes are objective (e.g., mortality, hospitalization).  Trials that are described as double or triple blind. |
| **Probably low risk of bias** |  |
| **Probably high risk of bias** |  |
| **Definitely high risk of bias** | Trials in which patients are not blind and in which outcomes are patient-reported (e.g., ACQ).  Trials in which outcome adjudicators are not blind and the outcomes are not objective (e.g., adverse events leading to discontinuation). |
| **Bias in selection of the reported results** | |
| Issues to consider:  Selective reporting of timepoints  Selective reporting of outcome measures  *Note that we are only interested in selective reporting for the outcomes for which we are extracting data.*  *Note that the judgments may differ across outcomes.* | |
| **Definitely low risk of bias** | Results for outcomes that were analyzed and reported according to a pre-specified statistical analysis plan or protocol (including the timepoint for the measurement of the outcome). |
| **Probably low risk of bias** | Results for outcomes that were analyzed and reported but that were not prespecified in a statistical analysis plan or protocol but the timepoint at which results are reported is consistent with the timepoint for other outcomes in the trial report or there is little reason to believe the outcome was selectively reported.  Please note that outcomes that were not prespecified in a protocol or statistical analysis plan and that are reported in the trial preprint or publication should be rated at probably low risk of bias unless there are other important reasons to suspect that results for those outcomes were selectively reported (e.g., results are presented at timepoints that don’t match the timepoints reported for other outcomes). |
| **Probably high risk of bias** | Results for outcomes that were analyzed and reported but that were not prespecified in a statistical analysis plan or protocol but the timepoint at which results are reported is not consistent with the timepoint for other outcomes in the trial report or there are other reasons to believe that the outcome is selectively reported. |
| **Definitely high risk of bias** | Results for outcomes that were analyzed and reported for which there are inconsistencies with the statistical analysis plan or protocol. These inconsistencies may include outcome measures of interest or the timepoints for the measurement of outcomes. |

### eTable 3. Risk of bias assessments

| **Author** | **Study** | **Outcome** | **Bias arising from the randomization process** | **Bias due to deviations from the intended intervention** | **Bias due to missing outcome data** | **Bias in measurement of the outcome** | **Bias in selection of the reported results** |
| --- | --- | --- | --- | --- | --- | --- | --- |
| Brightling | Luster 1 | ACQ | Low | Low | Low | Low | Low |
| Brightling | Luster 5 | ACQ | Low | Low | Low | Low | Low |
| Nair | Zonda | ACQ | Low | Low | Low | Low | Low |
| Berstein | trial 1 | ACQ | Low | Low | Low | Low | Low |
| Ferguson | BISE | ACQ | Low | Low | Low | Low | Low |
| Castro 2015 | trial 1 | ACQ | Low | Low | Low | Low | Low |
| Castro 2015 | trial 2 | ACQ | Low | Low | Low | Low | Low |
| Pavord | DREAM | ACQ | Low | Low | Low | Low | Low |
| Wechsler 2022 | SOURCE | ACQ | Low | Low | Low | Low | Low |
| Wechsler 2021 | NR | ACQ | Low | Low | Low | Low | Low |
| Wenzel 2013 | NR | ACQ | Low | Low | Low | Low | Low |
| Gonem | NR | ACQ | Low | Low | Low | Low | Low |
| Hayashi | NR | ACQ | Low | Low | Low | Low | Low |
| Li | NR | ACQ | Low | Low | Low | Low | Low |
| Ortega | MENSA | ACQ | Low | Low | Low | Low | Low |
| Chupp | MUSCA | ACQ | Low | Low | Low | Low | Low |
| Corren | PATHWAY | ACQ | Low | Low | Low | Low | Low |
| Bleecker | SIROCCO | ACQ | Low | Low | Low | Low | Low |
| Sverrild | UPSTREAM | ACQ | Low | Low | Low | Low | Low |
| Harrison | ANDHI | ACQ | Probably low | Low | Low | Low | Low |
| Bjermer | NR | ACQ | Probably low | Low | Low | Low | Low |
| Castro 2011 | NR | ACQ | Probably low | Low | Low | Low | Low |
| Corren | NR | ACQ | Probably low | Low | Low | Low | Low |
| Erpenbeck | NR | ACQ | Probably low | Low | Low | Low | Low |
| Panettieri | SOLANA | ACQ | Probably low | Low | Low | Low | Low |
| Kopp | EUDRACT | ACQ | Probably low | Low | Low | Low | Low |
| Menzies-Gow | NAVIGATOR | ACQ | Probably low | Low | Low | Low | Low |
| Nowak | NR | ACQ | Probably low | Low | Low | Low | Low |
| Bel | SIRIUS | ACQ | Probably low | Low | Low | Low | Low |
| FitzGerald | CALIMA | ACQ | Low | Low | Probably high | Low | Low |
| Wenzel 2016 | NR | ACQ | Low | Low | Probably low | Low | Low |
| Russell | MESOS | ACQ | Low | Low | Low | Low | Probably low |
| Ohta | NR | ACQ | Low | Low | Low | Low | Probably low |
| Park | NR | ACQ | Low | Low | Low | Low | Probably low |
| Flood-Page | NA | ACQ | Probably low | Low | Low | Low | Probably low |
| Pineros | MATERIAL | ACQ | Probably low | Low | Low | Low | Probably low |
| Piper | NR | ACQ | Probably low | Low | Low | Low | Probably low |
| Humbert | INNOVATE | Adverse events leading to discontinuation | Probably low | Low | Probably high | Low | High |
| Panettieri | STRATOS 1 | Adverse events leading to discontinuation | Low | Low | Low | Low | Low |
| Panettieri | STRATOS 2 | Adverse events leading to discontinuation | Low | Low | Low | Low | Low |
| Brightling | Luster 1 | Adverse events leading to discontinuation | Low | Low | Low | Low | Low |
| Brightling | Luster 3 | Adverse events leading to discontinuation | Low | Low | Low | Low | Low |
| Nair | Zonda | Adverse events leading to discontinuation | Low | Low | Low | Low | Low |
| Berstein | trial 1 | Adverse events leading to discontinuation | Low | Low | Low | Low | Low |
| Berstein | trial 2 | Adverse events leading to discontinuation | Low | Low | Low | Low | Low |
| Ferguson | BISE | Adverse events leading to discontinuation | Low | Low | Low | Low | Low |
| FitzGerald | CALIMA | Adverse events leading to discontinuation | Low | Low | Low | Low | Low |
| Castro 2015 | trial 1 | Adverse events leading to discontinuation | Low | Low | Low | Low | Low |
| Castro 2015 | trial 2 | Adverse events leading to discontinuation | Low | Low | Low | Low | Low |
| Pavord | DREAM | Adverse events leading to discontinuation | Low | Low | Low | Low | Low |
| Wechsler 2022 | SOURCE | Adverse events leading to discontinuation | Low | Low | Low | Low | Low |
| Wechsler 2021 | NR | Adverse events leading to discontinuation | Low | Low | Low | Low | Low |
| Castro 2021 | Zeal-1 | Adverse events leading to discontinuation | Low | Low | Low | Low | Low |
| Castro 2021 | Zeal-2 | Adverse events leading to discontinuation | Low | Low | Low | Low | Low |
| Kelsen | ZENYATTA | Adverse events leading to discontinuation | Low | Low | Low | Low | Low |
| Haldar | NR | Adverse events leading to discontinuation | Low | Low | Low | Low | Low |
| Hanania | NR | Adverse events leading to discontinuation | Low | Low | Low | Low | Low |
| Li | NR | Adverse events leading to discontinuation | Low | Low | Low | Low | Low |
| Ortega | MENSA | Adverse events leading to discontinuation | Low | Low | Low | Low | Low |
| Russell | MESOS | Adverse events leading to discontinuation | Low | Low | Low | Low | Low |
| Chupp | MUSCA | Adverse events leading to discontinuation | Low | Low | Low | Low | Low |
| Corren | PATHWAY | Adverse events leading to discontinuation | Low | Low | Low | Low | Low |
| Bleecker | SIROCCO | Adverse events leading to discontinuation | Low | Low | Low | Low | Low |
| Bjermer | NR | Adverse events leading to discontinuation | Probably low | Low | Low | Low | Low |
| Castro 2011 | NR | Adverse events leading to discontinuation | Probably low | Low | Low | Low | Low |
| Chanez | NR | Adverse events leading to discontinuation | Probably low | Low | Low | Low | Low |
| Corren | NR | Adverse events leading to discontinuation | Probably low | Low | Low | Low | Low |
| Panettieri | SOLANA | Adverse events leading to discontinuation | Probably low | Low | Low | Low | Low |
| Kopp | EUDRACT | Adverse events leading to discontinuation | Probably low | Low | Low | Low | Low |
| Rabe | LIBERTY ASTHMA VENTURE | Adverse events leading to discontinuation | Probably low | Low | Low | Low | Low |
| Menzies-Gow | NAVIGATOR | Adverse events leading to discontinuation | Probably low | Low | Low | Low | Low |
| Nowak | NR | Adverse events leading to discontinuation | Probably low | Low | Low | Low | Low |
| Bel | SIRIUS | Adverse events leading to discontinuation | Probably low | Low | Low | Low | Low |
| Castro | LIBERTY ASTHMA QUEST | Adverse events leading to discontinuation | Low | Low | Low | Low | Low |
| Wenzel 2016 | NR | Adverse events leading to discontinuation | Low | Low | Probably low | Low | Low |
| Castro | LIBERTY ASTHMA QUEST | Adverse events leading to discontinuation | Low | Low | Probably low | Low | Low |
| Vignola | Solar | Adverse events leading to discontinuation | Probably low | Low | Low | Low | Probably high |
| Ayres | NR | Adverse events leading to discontinuation | Probably high | Probably high | Low | Low | Probably high |
| Rubin | QUALITX | Adverse events leading to discontinuation | Probably high | Probably high | Low | High | Probably high |
| Ohta | NR | Adverse events leading to discontinuation | Low | Low | Low | Low | Probably low |
| Bardelas | NR | Adverse events leading to discontinuation | Probably low | Low | Low | Low | Probably low |
| Bateman | NR | Adverse events leading to discontinuation | Probably low | Low | Low | Low | Probably low |
| Busse | NR | Adverse events leading to discontinuation | Probably low | Low | Low | Low | Probably low |
| Soler | NR | Adverse events leading to discontinuation | Probably low | Low | Low | Low | Probably low |
| Flood-Page | NA | Adverse events leading to discontinuation | Probably low | Low | Low | Low | Probably low |
| Holgate | NR | Adverse events leading to discontinuation | Probably low | Low | Low | Low | Probably low |
| Pineros | MATERIAL | Adverse events leading to discontinuation | Probably low | Low | Low | Low | Probably low |
| Piper | NR | Adverse events leading to discontinuation | Probably low | Low | Low | Low | Probably low |
| Humbert | INNOVATE | Exacerbations | Probably low | Low | Probably high | Low | High |
| Panettieri | STRATOS 1 | Exacerbations | Low | Low | Low | Low | Low |
| Panettieri | STRATOS 2 | Exacerbations | Low | Low | Low | Low | Low |
| Brightling | Luster 1 | Exacerbations | Low | Low | Low | Low | Low |
| Brightling | Luster 2 | Exacerbations | Low | Low | Low | Low | Low |
| Nair | Zonda | Exacerbations | Low | Low | Low | Low | Low |
| Berstein | trial 1 | Exacerbations | Low | Low | Low | Low | Low |
| Ferguson | BISE | Exacerbations | Low | Low | Low | Low | Low |
| FitzGerald | CALIMA | Exacerbations | Low | Low | Low | Low | Low |
| Castro 2015 | trial 1 | Exacerbations | Low | Low | Low | Low | Low |
| Castro 2015 | trial 2 | Exacerbations | Low | Low | Low | Low | Low |
| Pavord | DREAM | Exacerbations | Low | Low | Low | Low | Low |
| Wechsler 2022 | SOURCE | Exacerbations | Low | Low | Low | Low | Low |
| Wechsler 2021 | NR | Exacerbations | Low | Low | Low | Low | Low |
| Wenzel 2013 | NR | Exacerbations | Low | Low | Low | Low | Low |
| Kelsen | ZENYATTA | Exacerbations | Low | Low | Low | Low | Low |
| Haldar | NR | Exacerbations | Low | Low | Low | Low | Low |
| Hanania | NR | Exacerbations | Low | Low | Low | Low | Low |
| Li | NR | Exacerbations | Low | Low | Low | Low | Low |
| Ortega | MENSA | Exacerbations | Low | Low | Low | Low | Low |
| Corren | PATHWAY | Exacerbations | Low | Low | Low | Low | Low |
| Bleecker | SIROCCO | Exacerbations | Low | Low | Low | Low | Low |
| Harrison | ANDHI | Exacerbations | Probably low | Low | Low | Low | Low |
| Castro 2011 | NR | Exacerbations | Probably low | Low | Low | Low | Low |
| Rabe | LIBERTY ASTHMA VENTURE | Exacerbations | Probably low | Low | Low | Low | Low |
| Menzies-Gow | NAVIGATOR | Exacerbations | Probably low | Low | Low | Low | Low |
| Nowak | NR | Exacerbations | Probably low | Low | Low | Low | Low |
| Bel | SIRIUS | Exacerbations | Probably low | Low | Low | Low | Low |
| Wenzel 2016 | NR | Exacerbations | Low | Low | Probably low | Low | Low |
| Vignola | Solar | Exacerbations | Probably low | Low | Low | Low | Probably high |
| Ayres | NR | Exacerbations | Probably high | Probably high | Probably low | High | Probably high |
| Chupp | MUSCA | Exacerbations | Low | Low | Low | Low | Probably low |
| Ohta | NR | Exacerbations | Low | Low | Low | Low | Probably low |
| Park | NR | Exacerbations | Low | Low | Low | Low | Probably low |
| Busse | NR | Exacerbations | Probably low | Low | Low | Low | Probably low |
| Soler | NR | Exacerbations | Probably low | Low | Low | Low | Probably low |
| Flood-Page | NA | Exacerbations | Probably low | Low | Low | Low | Probably low |
| Piper | NR | Exacerbations | Probably low | Low | Low | Low | Probably low |
| Castro 2015 | trial 1 | FEV1 | Low | Low | Low | Low | Low |
| Castro 2015 | trial 2 | FEV1 | Low | Low | Low | Low | Low |
| Pavord | DREAM | FEV1 | Low | Low | Low | Low | Low |
| Wechsler 2022 | SOURCE | FEV1 | Low | Low | Low | Low | Low |
| Wechsler 2021 | NR | FEV1 | Low | Low | Low | Low | Low |
| Wenzel 2013 | NR | FEV1 | Low | Low | Low | Low | Low |
| Castro 2021 | Zeal-1 | FEV1 | Low | Low | Low | Low | Low |
| Castro 2021 | Zeal-2 | FEV1 | Low | Low | Low | Low | Low |
| Kelsen | ZENYATTA | FEV1 | Low | Low | Low | Low | Low |
| Gonem | NR | FEV1 | Low | Low | Low | Low | Low |
| Ortega | MENSA | FEV1 | Low | Low | Low | Low | Low |
| Chupp | MUSCA | FEV1 | Low | Low | Low | Low | Low |
| Corren | PATHWAY | FEV1 | Low | Low | Low | Low | Low |
| Bleecker | SIROCCO | FEV1 | Low | Low | Low | Low | Low |
| Sverrild | UPSTREAM | FEV1 | Low | Low | Low | Low | Low |
| Corren | NR | FEV1 | Probably low | Low | Low | Low | Low |
| Erpenbeck | NR | FEV1 | Probably low | Low | Low | Low | Low |
| Panettieri | SOLANA | FEV1 | Probably low | Low | Low | Low | Low |
| Rabe | LIBERTY ASTHMA VENTURE | FEV1 | Probably low | Low | Low | Low | Low |
| Menzies-Gow | NAVIGATOR | FEV1 | Probably low | Low | Low | Low | Low |
| Nowak | NR | FEV1 | Probably low | Low | Low | Low | Low |
| Wenzel 2016 | NR | FEV1 | Low | Low | Probably low | Low | Low |
| Castro | LIBERTY ASTHMA QUEST | FEV1 | Low | Low | Probably low | Low | Low |
| Rubin | QUALITX | FEV1 | Probably high | Probably high | Low | Probably low | Probably high |
| Russell | MESOS | FEV1 | Low | Low | Low | Low | Probably low |
| Flood-Page | NA | FEV1 | Probably low | Low | Low | Low | Probably low |
| Piper | NR | FEV1 | Probably low | Low | Low | Low | Probably low |
| Hayashi | NR | FEV1 % | Low | Low | Low | Low | Low |
| Li | NR | FEV1 % | Low | Low | Low | Low | Low |
| Corren | PATHWAY | FEV1 % | Low | Low | Low | Low | Low |
| Panettieri | STRATOS 1 | FEV1 % | Low | Low | Low | Low | Low |
| Brightling | Luster 1 | FEV1 % | Low | Low | Low | Low | Low |
| Brightling | Luster 4 | FEV1 % | Low | Low | Low | Low | Low |
| Nair | Zonda | FEV1 % | Low | Low | Low | Low | Low |
| Berstein | trial 1 | FEV1 % | Low | Low | Low | Low | Low |
| Ferguson | BISE | FEV1 % | Low | Low | Low | Low | Low |
| Djukanovic | NR | FEV1 % | Probably low | Low | Low | Low | Low |
| Nowak | NR | FEV1 % | Probably low | Low | Low | Low | Low |
| Bel | SIRIUS | FEV1 % | Probably low | Low | Low | Low | Low |
| Harrison | ANDHI | FEV1 % | Probably low | Low | Low | Low | Low |
| Bjermer | NR | FEV1 % | Probably low | Low | Low | Low | Low |
| Castro 2011 | NR | FEV1 % | Probably low | Low | Low | Low | Low |
| FitzGerald | CALIMA | FEV1 % | Low | Low | Probably high | Low | Low |
| Castro | LIBERTY ASTHMA QUEST | FEV1 % | Low | Low | Probably low | Low | Low |
| Park | NR | FEV1 % | Low | Low | Low | Low | Probably low |
| Pineros | MATERIAL | FEV1 % | Probably low | Low | Low | Low | Probably low |
| Soler | NR | FEV1 % | Probably low | Low | Low | Low | Probably low |
| Bardelas | NR | FEV1 % | Probably low | Low | Low | Low | Probably low |
| Boulet | NR | FEV1 % | Probably low | Low | Low | Low | Probably low |
| Humbert | INNOVATE | Hospitalizations or hospitalization and ED visits | Probably low | Low | Probably high | Low | High |
| Panettieri | STRATOS 1 | Hospitalizations or hospitalization and ED visits | Low | Low | Low | Low | Low |
| Panettieri | STRATOS 2 | Hospitalizations or hospitalization and ED visits | Low | Low | Low | Low | Low |
| Nair | Zonda | Hospitalizations or hospitalization and ED visits | Low | Low | Low | Low | Low |
| Ferguson | BISE | Hospitalizations or hospitalization and ED visits | Low | Low | Low | Low | Low |
| FitzGerald | CALIMA | Hospitalizations or hospitalization and ED visits | Low | Low | Low | Low | Low |
| Pavord | DREAM | Hospitalizations or hospitalization and ED visits | Low | Low | Low | Low | Low |
| Wechsler 2021 | NR | Hospitalizations or hospitalization and ED visits | Low | Low | Low | Low | Low |
| Wenzel 2013 | NR | Hospitalizations or hospitalization and ED visits | Low | Low | Low | Low | Low |
| Haldar | NR | Hospitalizations or hospitalization and ED visits | Low | Low | Low | Low | Low |
| Ortega | MENSA | Hospitalizations or hospitalization and ED visits | Low | Low | Low | Low | Low |
| Corren | PATHWAY | Hospitalizations or hospitalization and ED visits | Low | Low | Low | Low | Low |
| Bleecker | SIROCCO | Hospitalizations or hospitalization and ED visits | Low | Low | Low | Low | Low |
| Menzies-Gow | NAVIGATOR | Hospitalizations or hospitalization and ED visits | Probably low | Low | Low | Low | Low |
| Nowak | NR | Hospitalizations or hospitalization and ED visits | Probably low | Low | Low | Low | Low |
| Ayres | NR | Hospitalizations or hospitalization and ED visits | Probably high | Probably high | Low | Low | Probably high |
| Chupp | MUSCA | Hospitalizations or hospitalization and ED visits | Low | Low | Low | Low | Probably low |
| Busse | NR | Hospitalizations or hospitalization and ED visits | Probably low | Low | Low | Low | Probably low |
| Humbert | INNOVATE | Patients with 50% or more reduction in use of oral corticosteroids | Probably low | Low | Probably high | Low | High |
| Nair | Zonda | Patients with 50% or more reduction in use of oral corticosteroids | Low | Low | Low | Low | Low |
| Berstein | trial 2 | Patients with 50% or more reduction in use of oral corticosteroids | Low | Low | Low | Low | Low |
| Wechsler 2022 | SOURCE | Patients with 50% or more reduction in use of oral corticosteroids | Low | Low | Low | Low | Low |
| Wenzel 2013 | NR | Patients with 50% or more reduction in use of oral corticosteroids | Low | Low | Low | Low | Low |
| Rabe | LIBERTY ASTHMA VENTURE | Patients with 50% or more reduction in use of oral corticosteroids | Probably low | Low | Low | Low | Low |
| Bel | SIRIUS | Patients with 50% or more reduction in use of oral corticosteroids | Probably low | Low | Low | Low | Low |
| Busse | NR | Patients with 50% or more reduction in use of oral corticosteroids | Probably low | Low | Low | Low | Probably low |
| Holgate | NR | Patients with 50% or more reduction in use of oral corticosteroids | Probably low | Low | Low | Low | Probably low |

### eTable 4. Exacerbations network estimates

**Exacerbations (all patients)**

| ***Exacerbations*** | ***Comparison*** | ***Network estimate*** | | | ***Network estimate*** | | | |
| --- | --- | --- | --- | --- | --- | --- | --- | --- |
| ***MCID 20%*** | **Baseline risk 47%** |  |  |  |  | | | |
| *Treatment 1* | Treatment 2 | Point estimate | CI Lower limit | CI upper limit | Point estimate | CI Lower limit | CI upper limit | **Final rating** |
| *Tau^2=0.0293* | I^2=48.9% |  | | | | | | |
| Astegolimab | Benralizumab | 1.08 | 0.66 | 1.74 | 22.936 | -97.478 | 212.158 | **Moderate** |
| Benralizumab | Dupilumab | 1.33 | 1 | 1.77 | 71.346 | 0 | 166.474 | **High** |
| Astegolimab | Dupilumab | 1.43 | 0.86 | 2.37 | 92.966 | -30.268 | 296.194 | **Moderate** |
| Benralizumab | Fevipiprant | 0.74 | 0.53 | 1.03 | -101.426 | -183.347 | 11.703 | **High** |
| Astegolimab | Fevipiprant | 0.79 | 0.46 | 1.35 | -81.921 | -210.654 | 136.535 | **Moderate** |
| Dupilumab | Fevipiprant | 0.55 | 0.38 | 0.8 | -175.545 | -241.862 | -78.02 | **Moderate** |
| Benralizumab | Mepolizumab | 1.11 | 0.85 | 1.44 | 28.435 | -38.775 | 113.74 | **High** |
| Dupilumab | Mepolizumab | 0.84 | 0.62 | 1.13 | -41.36 | -98.23 | 33.605 | **High** |
| Astegolimab | Mepolizumab | 1.19 | 0.73 | 1.95 | 49.115 | -69.795 | 245.575 | **Moderate** |
| Fevipiprant | Mepolizumab | 1.5 | 1.06 | 2.13 | 129.25 | 15.51 | 292.105 | **Moderate** |
| Benralizumab | Omalizumab | 1.02 | 0.8 | 1.31 | 5.64 | -56.4 | 87.42 | **High** |
| Dupilumab | Omalizumab | 0.77 | 0.58 | 1.03 | -64.86 | -118.44 | 8.46 | **High** |
| Mepolizumab | Omalizumab | 0.92 | 0.71 | 1.2 | -22.56 | -81.78 | 56.4 | **High** |
| Astegolimab | Omalizumab | 1.1 | 0.68 | 1.78 | 28.2 | -90.24 | 219.96 | **Moderate** |
| Fevipiprant | Omalizumab | 1.39 | 0.99 | 1.94 | 109.98 | -2.82 | 265.08 | **Moderate** |
| Fevipiprant | Placebo | 0.83 | 0.62 | 1.1 | -79.9 | -178.6 | 47 | **Moderate** |
| Tralokinumab | Placebo | 0.93 | 0.7 | 1.25 | -32.9 | -141 | 117.5 | **High** |
| Astegolimab | Placebo | 0.66 | 0.42 | 1.03 | -159.8 | -272.6 | 14.1 | **Moderate** |
| Benralizumab | Placebo | 0.61 | 0.51 | 0.73 | -183.3 | -230.3 | -126.9 | **Moderate** |
| Dupilumab | Placebo | 0.46 | 0.37 | 0.58 | -253.8 | -296.1 | -197.4 | **Moderate** |
| Mepolizumab | Placebo | 0.55 | 0.45 | 0.67 | -211.5 | -258.5 | -155.1 | **Moderate** |
| Omalizumab | Placebo | 0.6 | 0.5 | 0.71 | -188 | -235 | -136.3 | **Moderate** |
| Reslizumab | Placebo | 0.54 | 0.42 | 0.7 | -216.2 | -272.6 | -141 | **Moderate** |
| Tezepelumab | Placebo | 0.45 | 0.35 | 0.59 | -258.5 | -305.5 | -192.7 | **Moderate** |
| Benralizumab | Reslizumab | 1.12 | 0.83 | 1.52 | 30.456 | -43.146 | 131.976 | **High** |
| Dupilumab | Reslizumab | 0.85 | 0.6 | 1.19 | -38.07 | -101.52 | 48.222 | **High** |
| Mepolizumab | Reslizumab | 1.01 | 0.74 | 1.39 | 2.538 | -65.988 | 98.982 | **High** |
| Omalizumab | Reslizumab | 1.1 | 0.81 | 1.49 | 25.38 | -48.222 | 124.362 | **High** |
| Astegolimab | Reslizumab | 1.21 | 0.72 | 2.03 | 53.298 | -71.064 | 261.414 | **Moderate** |
| Fevipiprant | Reslizumab | 1.52 | 1.04 | 2.23 | 131.976 | 10.152 | 312.174 | **Moderate** |
| Placebo | Reslizumab | 1.84 | 1.43 | 2.36 | 213.192 | 109.134 | 345.168 | **Moderate** |
| Benralizumab | Tezepelumab | 1.36 | 0.99 | 1.86 | 76.14 | -2.115 | 181.89 | **High** |
| Dupilumab | Tezepelumab | 1.02 | 0.72 | 1.45 | 4.23 | -59.22 | 95.175 | **High** |
| Mepolizumab | Tezepelumab | 1.22 | 0.88 | 1.7 | 46.53 | -25.38 | 148.05 | **High** |
| Omalizumab | Tezepelumab | 1.32 | 0.96 | 1.82 | 67.68 | -8.46 | 173.43 | **High** |
| Reslizumab | Tezepelumab | 1.21 | 0.84 | 1.74 | 44.415 | -33.84 | 156.51 | **High** |
| Astegolimab | Tezepelumab | 1.46 | 0.86 | 2.46 | 97.29 | -29.61 | 308.79 | **Moderate** |
| Fevipiprant | Tezepelumab | 1.84 | 1.24 | 2.72 | 177.66 | 50.76 | 363.78 | **Moderate** |
| Placebo | Tezepelumab | 2.22 | 1.7 | 2.89 | 258.03 | 148.05 | 399.735 | **Moderate** |
| Fevipiprant | Tralokinumab | 0.88 | 0.59 | 1.33 | -52.452 | -179.211 | 144.243 | **High** |
| Placebo | Tralokinumab | 1.07 | 0.8 | 1.42 | 30.597 | -87.42 | 183.582 | **High** |
| Astegolimab | Tralokinumab | 0.7 | 0.41 | 1.2 | -131.13 | -257.889 | 87.42 | **Moderate** |
| Benralizumab | Tralokinumab | 0.65 | 0.47 | 0.91 | -152.985 | -231.663 | -39.339 | **Moderate** |
| Dupilumab | Tralokinumab | 0.49 | 0.34 | 0.71 | -222.921 | -288.486 | -126.759 | **Moderate** |
| Mepolizumab | Tralokinumab | 0.59 | 0.41 | 0.83 | -179.211 | -257.889 | -74.307 | **Moderate** |
| Omalizumab | Tralokinumab | 0.64 | 0.46 | 0.89 | -157.356 | -236.034 | -48.081 | **Moderate** |
| Reslizumab | Tralokinumab | 0.58 | 0.4 | 0.85 | -183.582 | -262.26 | -65.565 | **Moderate** |
| Tezepelumab | Tralokinumab | 0.48 | 0.32 | 0.71 | -227.292 | -297.228 | -126.759 | **Moderate** |

### eTable 5. Network estimates exacerbations (eos >=300)

| ***Exacerbations (high)*** | ***Comparison*** | ***Network estimate*** | | | ***Network estimate*** | | | |
| --- | --- | --- | --- | --- | --- | --- | --- | --- |
| ***MCID 20%*** | **Baseline risk 47%** |  |  |  | **Absolute risk difference per 1000** | | | |
| *Treatment 1* | Treatment 2 | Point estimate | CI Lower limit | CI upper limit | Point estimate | CI Lower limit | CI upper limit | **Final rating** |
| *Tau^2=0.0244* | I^2=36.3 |  |  |  |  |  |  |  |
| Omalizumab | astegolimab | 1.31 | 0.59 | 2.9 | 99.076 | -131.036 | 607.24 | **Very low** |
| Astegolimab | Benralizumab | 1.34 | 0.64 | 2.8 | 81.498 | -86.292 | 431.46 | **Low** |
| Omalizumab | benralizumab | 0.97 | 0.72 | 1.3 | -7.191 | -67.116 | 71.91 | **Low** |
| Benralizumab | Dupilumab | 1.59 | 1.13 | 2.26 | 88.736 | 19.552 | 189.504 | **Moderate** |
| Astegolimab | Dupilumab | 2.13 | 0.99 | 4.55 | 169.952 | -1.504 | 533.92 | **Low** |
| Omalizumab | dupilumab | 0.62 | 0.4 | 0.96 | -57.152 | -90.24 | -6.016 | **Moderate** |
| Dupilumab | Fevipiprant | 0.39 | 0.26 | 0.58 | -232.227 | -281.718 | -159.894 | **Moderate** |
| Benralizumab | Fevipiprant | 0.62 | 0.43 | 0.89 | -144.666 | -216.999 | -41.877 | **Moderate** |
| Astegolimab | Fevipiprant | 0.83 | 0.39 | 1.79 | -64.719 | -232.227 | 300.753 | **Low** |
| Omalizumab | fevipiprant | 1.58 | 1 | 2.48 | 220.806 | 0 | 563.436 | **Very low** |
| Dupilumab | Mepolizumab | 0.61 | 0.43 | 0.85 | -95.316 | -139.308 | -36.66 | **High** |
| Benralizumab | Mepolizumab | 0.97 | 0.72 | 1.3 | -7.332 | -68.432 | 73.32 | **High** |
| Astegolimab | Mepolizumab | 1.29 | 0.62 | 2.7 | 70.876 | -92.872 | 415.48 | **Low** |
| Fevipiprant | Mepolizumab | 1.56 | 1.09 | 2.21 | 136.864 | 21.996 | 295.724 | **Moderate** |
| Omalizumab | mepolizumab | 1.02 | 0.68 | 1.51 | 4.888 | -78.208 | 124.644 | **Low** |
| Tezepelumab | Placebo | 0.3 | 0.22 | 0.42 | -329 | -366.6 | -272.6 | **High** |
| Dupilumab | Placebo | 0.32 | 0.24 | 0.42 | -319.6 | -357.2 | -272.6 | **High** |
| Benralizumab | Placebo | 0.51 | 0.41 | 0.63 | -230.3 | -277.3 | -173.9 | **Moderate** |
| Resliuzmab | Placebo | 0.51 | 0.4 | 0.65 | -230.3 | -282 | -164.5 | **Moderate** |
| Mepolizumab | Placebo | 0.52 | 0.43 | 0.64 | -225.6 | -267.9 | -169.2 | **Moderate** |
| Astegolimab | Placebo | 0.68 | 0.33 | 1.38 | -150.4 | -314.9 | 178.6 | **Low** |
| Fevipiprant | Placebo | 0.81 | 0.61 | 1.09 | -89.3 | -183.3 | 42.3 | **High** |
| Tralokinumab | Placebo | 0.85 | 0.62 | 1.16 | -70.5 | -178.6 | 75.2 | **High** |
| Omalizumab | Placebo | 0.52 | 0.37 | 0.72 | -225.6 | -296.1 | -131.6 | **High** |
| Dupilumab | Resliuzmab | 0.62 | 0.43 | 0.9 | -91.086 | -136.629 | -23.97 | **High** |
| Benralizumab | Resliuzmab | 0.99 | 0.72 | 1.37 | -2.397 | -67.116 | 88.689 | **High** |
| Mepolizumab | Resliuzmab | 1.03 | 0.75 | 1.4 | 7.191 | -59.925 | 95.88 | **High** |
| Astegolimab | Resliuzmab | 1.33 | 0.63 | 2.81 | 79.101 | -88.689 | 433.857 | **Low** |
| Fevipiprant | Resliuzmab | 1.6 | 1.09 | 2.33 | 143.82 | 21.573 | 318.801 | **Moderate** |
| Omalizumab | Reslizumab | 1.01 | 0.66 | 1.54 | 2.397 | -81.498 | 129.438 | **Very low** |
| Dupilumab | Tezepelumab | 1.04 | 0.68 | 1.6 | 5.64 | -45.12 | 84.6 | **High** |
| Benralizumab | Tezepelumab | 1.66 | 1.12 | 2.46 | 93.06 | 16.92 | 205.86 | **Moderate** |
| Resliuzmab | Tezepelumab | 1.67 | 1.11 | 2.52 | 94.47 | 15.51 | 214.32 | **Moderate** |
| Mepolizumab | Tezepelumab | 1.72 | 1.17 | 2.52 | 101.52 | 23.97 | 214.32 | **Moderate** |
| Astegolimab | Tezepelumab | 2.22 | 1.02 | 4.85 | 172.02 | 2.82 | 542.85 | **Low** |
| Fevipiprant | Tezepelumab | 2.67 | 1.72 | 4.14 | 235.47 | 101.52 | 442.74 | **Low** |
| Omalizumab | Tezepelumab | 1.69 | 1.05 | 2.72 | 97.29 | 7.05 | 242.52 | **Moderate** |
| Tezepelumab | Tralokinumab | 0.36 | 0.23 | 0.57 | -255.68 | -307.615 | -171.785 | **Moderate** |
| Dupilumab | Tralokinumab | 0.37 | 0.25 | 0.57 | -251.685 | -299.625 | -171.785 | **Moderate** |
| Benralizumab | Tralokinumab | 0.6 | 0.41 | 0.88 | -159.8 | -235.705 | -47.94 | **Moderate** |
| Resliuzmab | Tralokinumab | 0.6 | 0.4 | 0.9 | -159.8 | -239.7 | -39.95 | **Moderate** |
| Mepolizumab | Tralokinumab | 0.62 | 0.42 | 0.9 | -151.81 | -231.71 | -39.95 | **Moderate** |
| Astegolimab | Tralokinumab | 0.8 | 0.37 | 1.73 | -79.9 | -251.685 | 291.635 | **Low** |
| Fevipiprant | Tralokinumab | 0.96 | 0.62 | 1.47 | -15.98 | -151.81 | 187.765 | **Moderate** |
| Omalizumab | Tralokinumab | 0.61 | 0.38 | 0.97 | -155.805 | -247.69 | -11.985 | **Moderate** |

### eTable 6. Network estimates exacerbations (eos <300)

| ***Exacerbations (Low)*** | ***Comparison*** | ***Network estimate*** | | | ***Network estimate*** | | | |
| --- | --- | --- | --- | --- | --- | --- | --- | --- |
| ***MCID 20%*** | **Baseline risk 47%** | **Relative estimate** | | | **Absolute risk difference per 1000** | | | |
| *Treatment 1* | Treatment 2 | Point estimate | CI Lower limit | CI upper limit | Point estimate | CI Lower limit | CI upper limit | **Final rating** |
| Omalizumab | astegolimab | 0.74 | 0.34 | 1.64 | -89.206 | -226.446 | 219.584 | **Low** |
| Astegolimab | Benralizumab | 1.06 | 0.48 | 2.34 | 20.586 | -178.412 | 459.754 | **Low** |
| Omalizumab | benralizumab | 0.7 | 0.36 | 1.36 | -102.93 | -219.584 | 123.516 | **Low** |
| Astegolimab | Dupilumab | 0.96 | 0.45 | 2.04 | -14.288 | -196.46 | 371.488 | **Low** |
| Benralizumab | Dupilumab | 0.91 | 0.51 | 1.63 | -32.148 | -175.028 | 225.036 | **Low** |
| Omalizumab | dupilumab | 0.76 | 0.54 | 1.09 | -85.728 | -164.312 | 32.148 | **Low** |
| Astegolimab | Placebo | 0.73 | 0.38 | 1.4 | -126.9 | -291.4 | 188 | **Low** |
| Benralizumab | Placebo | 0.69 | 0.44 | 1.08 | -145.7 | -263.2 | 37.6 | **Moderate** |
| Dupilumab | Placebo | 0.76 | 0.52 | 1.11 | -112.8 | -225.6 | 51.7 | **Moderate** |
| Omalizumab | Placebo | 0.98 | 0.59 | 1.63 | -9.4 | -192.7 | 296.1 | **Low** |
| Reslizumab | Placebo | 1.79 | 0.6 | 5.26 | 371.3 | -188 | 2002.2 | **Low** |
| Tezepelumab | Placebo | 0.63 | 0.41 | 0.95 | -173.9 | -277.3 | -23.5 | **Moderate** |
| Tralokinumab | Placebo | 0.81 | 0.48 | 1.37 | -89.3 | -244.4 | 173.9 | **Moderate** |
| Astegolimab | Reslizumab | 0.41 | 0.11 | 1.47 | -496.367 | -748.757 | 395.411 | **Low** |
| Benralizumab | Reslizumab | 0.39 | 0.12 | 1.26 | -513.193 | -740.344 | 218.738 | **Low** |
| Dupilumab | Reslizumab | 0.43 | 0.13 | 1.36 | -479.541 | -731.931 | 302.868 | **Low** |
| Omalizumab | Reslizumab | 0.55 | 0.17 | 1.8 | -378.585 | -698.279 | 673.04 | **Low** |
| Astegolimab | Tezepelumab | 1.17 | 0.54 | 2.54 | 50.337 | -136.206 | 455.994 | **Low** |
| Benralizumab | Tezepelumab | 1.1 | 0.6 | 2.04 | 29.61 | -118.44 | 307.944 | **Moderate** |
| Dupilumab | Tezepelumab | 1.22 | 0.69 | 2.14 | 65.142 | -91.791 | 337.554 | **Low** |
| Omalizumab | Tezepelumab | 1.57 | 0.82 | 2.99 | 168.777 | -53.298 | 589.239 | **Low** |
| Reslizumab | Tezepelumab | 2.85 | 0.88 | 9.22 | 547.785 | -35.532 | 2433.942 | **Low** |
| Astegolimab | Tralokinumab | 0.89 | 0.39 | 2.07 | -41.877 | -232.227 | 407.349 | **Low** |
| Benralizumab | Tralokinumab | 0.84 | 0.42 | 1.68 | -60.912 | -220.806 | 258.876 | **Low** |
| Dupilumab | Tralokinumab | 0.93 | 0.49 | 1.78 | -26.649 | -194.157 | 296.946 | **Low** |
| Omalizumab | Tralokinumab | 1.2 | 0.59 | 2.43 | 76.14 | -156.087 | 544.401 | **Low** |
| Reslizumab | Tralokinumab | 2.18 | 0.65 | 7.36 | 449.226 | -133.245 | 2421.252 | **Low** |
| Tezepelumab | Tralokinumab | 0.77 | 0.39 | 1.5 | -87.561 | -232.227 | 190.35 | **Low** |

### eTable 7. Network estimates ACQ (all)

| ***ACQ*** | ***Comparison*** | ***Network estimate*** | | | |
| --- | --- | --- | --- | --- | --- |
| ***MCID -0.5*** |  | **Relative estimate** | | | |
| *Treatment 1* | Treatment 2 | Point estimate | CI Lower limit | CI upper limit | **Final rating** |
| *Tau^2=0.0127* | I^2=52.5% |  |  |  |  |
| Benralizumab | Dupilumab | 0.31 | 0.12 | 0.51 | **Low** |
| Benralizumab | Fevipiprant | -0.09 | -0.28 | 0.1 | **High** |
| Dupilumab | Fevipiprant | -0.4 | -0.62 | -0.18 | **Low** |
| Benralizumab | Itepekimab | 0.24 | -0.12 | 0.6 | **Moderate** |
| Fevipiprant | Itepekimab | 0.33 | -0.04 | 0.7 | **Moderate** |
| Dupilumab | Itepekimab | -0.07 | -0.41 | 0.27 | **Moderate** |
| Itepekimab | Itepekimab/dupilumab | -0.1 | -0.48 | 0.28 | **High** |
| Benralizumab | Itepekimab/dupilumab | 0.14 | -0.22 | 0.5 | **Moderate** |
| Fevipiprant | Itepekimab/dupilumab | 0.23 | -0.14 | 0.6 | **Moderate** |
| Dupilumab | Itepekimab/dupilumab | -0.17 | -0.51 | 0.17 | **Low** |
| Benralizumab | Mepolizumab | 0.09 | -0.11 | 0.29 | **High** |
| Fevipiprant | Mepolizumab | 0.18 | -0.05 | 0.41 | **High** |
| Itepekimab/dupilumab | Mepolizumab | -0.05 | -0.43 | 0.33 | **High** |
| Itepekimab | Mepolizumab | -0.15 | -0.53 | 0.23 | **Moderate** |
| Dupilumab | Mepolizumab | -0.22 | -0.45 | 0.01 | **Moderate** |
| Benralizumab | Omalizumab | 0.03 | -0.25 | 0.31 | **High** |
| Fevipiprant | Omalizumab | 0.12 | -0.18 | 0.42 | **High** |
| Mepolizumab | Omalizumab | -0.06 | -0.37 | 0.24 | **High** |
| Itepekimab | Omalizumab | -0.21 | -0.64 | 0.22 | **Moderate** |
| Itepekimab/dupilumab | Omalizumab | -0.11 | -0.54 | 0.32 | **Moderate** |
| Dupilumab | Omalizumab | -0.28 | -0.59 | 0.02 | **Low** |
| Benralizumab | Placebo | -0.22 | -0.32 | -0.11 | **High** |
| Fevipiprant | Placebo | -0.13 | -0.28 | 0.03 | **High** |
| Itepekimab | Placebo | -0.46 | -0.8 | -0.12 | **High** |
| Reslizumab | Placebo | -0.32 | -0.46 | -0.18 | **High** |
| Itepekimab/dupilumab | Placebo | -0.36 | -0.7 | -0.02 | **Moderate** |
| Mepolizumab | Placebo | -0.31 | -0.47 | -0.14 | **Moderate** |
| Omalizumab | Placebo | -0.25 | -0.5 | 0.01 | **Moderate** |
| Tezepelumab | Placebo | -0.33 | -0.49 | -0.17 | **Moderate** |
| Tralokinumab | Placebo | -0.25 | -0.49 | -0.02 | **Moderate** |
| Dupilumab | Placebo | -0.53 | -0.69 | -0.37 | **Low** |
| Benralizumab | Reslizumab | 0.11 | -0.07 | 0.29 | **High** |
| Fevipiprant | Reslizumab | 0.19 | -0.02 | 0.4 | **High** |
| Itepekimab/dupilumab | Reslizumab | -0.04 | -0.4 | 0.33 | **High** |
| Mepolizumab | Reslizumab | 0.01 | -0.21 | 0.23 | **High** |
| Omalizumab | Reslizumab | 0.08 | -0.22 | 0.37 | **High** |
| Itepekimab | Reslizumab | -0.14 | -0.5 | 0.23 | **Moderate** |
| Dupilumab | Reslizumab | -0.21 | -0.42 | 0.01 | **Moderate** |
| Benralizumab | Tezepelumab | 0.11 | -0.08 | 0.3 | **High** |
| Fevipiprant | Tezepelumab | 0.2 | -0.02 | 0.42 | **High** |
| Itepekimab/dupilumab | Tezepelumab | -0.03 | -0.41 | 0.35 | **High** |
| Mepolizumab | Tezepelumab | 0.02 | -0.21 | 0.25 | **High** |
| Omalizumab | Tezepelumab | 0.08 | -0.22 | 0.38 | **High** |
| Reslizumab | Tezepelumab | 0 | -0.21 | 0.22 | **High** |
| Itepekimab | Tezepelumab | -0.13 | -0.51 | 0.25 | **Moderate** |
| Dupilumab | Tezepelumab | -0.2 | -0.43 | 0.02 | **Moderate** |
| Benralizumab | Tralokinumab | 0.04 | -0.22 | 0.3 | **High** |
| Fevipiprant | Tralokinumab | 0.13 | -0.16 | 0.41 | **High** |
| Mepolizumab | Tralokinumab | -0.05 | -0.34 | 0.24 | **High** |
| Omalizumab | Tralokinumab | 0.01 | -0.34 | 0.36 | **High** |
| Reslizumab | Tralokinumab | -0.07 | -0.34 | 0.21 | **High** |
| Tezepelumab | Tralokinumab | -0.07 | -0.36 | 0.21 | **High** |
| Itepekimab | Tralokinumab | -0.2 | -0.62 | 0.21 | **Moderate** |
| Itepekimab/dupilumab | Tralokinumab | -0.1 | -0.52 | 0.31 | **Moderate** |
| Dupilumab | Tralokinumab | -0.27 | -0.56 | 0.01 | **Low** |

### eTable 8. Network estimates ACQ (eos >=300)

| ***ACQ*** | ***Comparison*** | ***Network estimate*** | | | |
| --- | --- | --- | --- | --- | --- |
| ***MCID -0.5*** |  | **Relative estimate** | | | |
| *Treatment 1* | Treatment 2 | Point estimate | CI Lower limit | CI upper limit | **Final rating** |
| *Tau^2=0.0153* | I^2=50.1 |  |  |  |  |
| Benralizumab | Dupilumab | 0.43 | 0.14 | 0.72 | **Moderate** |
| Benralizumab | Fevipiprant | -0.15 | -0.4 | 0.1 | **High** |
| Dupilumab | Fevipiprant | -0.58 | -0.9 | -0.26 | **Moderate** |
| Benralizumab | Mepolizumab | 0.03 | -0.2 | 0.26 | **High** |
| Fevipiprant | Mepolizumab | 0.18 | -0.09 | 0.45 | **High** |
| Dupilumab | Mepolizumab | -0.4 | -0.71 | -0.09 | **Moderate** |
| Benralizumab | Placebo | -0.3 | -0.44 | -0.16 | **High** |
| Fevipiprant | Placebo | -0.15 | -0.35 | 0.05 | **High** |
| Reslizumab | Placebo | -0.28 | -0.44 | -0.11 | **High** |
| Dupilumab | Placebo | -0.73 | -0.98 | -0.48 | **Moderate** |
| Mepolizumab | Placebo | -0.33 | -0.51 | -0.15 | **Moderate** |
| Tezepelumab | Placebo | -0.4 | -0.61 | -0.19 | **Moderate** |
| Tralokinumab | Placebo | -0.52 | -0.94 | -0.1 | **Low** |
| Benralizumab | Reslizumab | -0.02 | -0.24 | 0.2 | **High** |
| Fevipiprant | Reslizumab | 0.13 | -0.13 | 0.39 | **High** |
| Mepolizumab | Reslizumab | -0.05 | -0.3 | 0.19 | **High** |
| Dupilumab | Reslizumab | -0.45 | -0.75 | -0.15 | **Moderate** |
| Benralizumab | Tezepelumab | 0.1 | -0.15 | 0.36 | **High** |
| Mepolizumab | Tezepelumab | 0.07 | -0.21 | 0.35 | **High** |
| Reslizumab | Tezepelumab | 0.12 | -0.14 | 0.39 | **High** |
| Dupilumab | Tezepelumab | -0.33 | -0.66 | 0 | **Moderate** |
| Fevipiprant | Tezepelumab | 0.25 | -0.04 | 0.54 | **Moderate** |
| Dupilumab | Tralokinumab | -0.21 | -0.7 | 0.28 | **High** |
| Benralizumab | Tralokinumab | 0.22 | -0.23 | 0.67 | **Moderate** |
| Fevipiprant | Tralokinumab | 0.37 | -0.1 | 0.84 | **Moderate** |
| Mepolizumab | Tralokinumab | 0.19 | -0.27 | 0.65 | **Moderate** |
| Reslizumab | Tralokinumab | 0.24 | -0.21 | 0.7 | **Moderate** |
| Tezepelumab | Tralokinumab | 0.12 | -0.35 | 0.59 | **Moderate** |

### eTable 9. Network estimates ACQ (eos <300)

| ***ACQ*** | ***Comparison*** | ***Network estimate*** | | | |
| --- | --- | --- | --- | --- | --- |
| ***MCID -0.5*** |  | **Relative estimate** | | | |
| *Treatment 1* | Treatment 2 | Point estimate | CI Lower limit | CI upper limit | **Final rating** |
| *Tau^2=<0.0001* | I^2=0% |  |  |  |  |
| Benralizumab | Dupilumab | -0.03 | -0.31 | 0.25 | **High** |
| Benralizumab | Mepolizumab | -0.72 | -1.24 | -0.21 | **Moderate** |
| Dupilumab | Mepolizumab | -0.69 | -1.22 | -0.16 | **Moderate** |
| Benralizumab | Placebo | -0.23 | -0.41 | -0.06 | **High** |
| Dupilumab | Placebo | -0.2 | -0.42 | 0.02 | **High** |
| Reslizumab | Placebo | 0.12 | -0.09 | 0.33 | **High** |
| Tezepelumab | Placebo | -0.23 | -0.36 | -0.09 | **High** |
| Mepolizumab | Placebo | 0.49 | 0.01 | 0.97 | **Moderate** |
| Benralizumab | Reslizumab | -0.35 | -0.63 | -0.08 | **Moderate** |
| Dupilumab | Reslizumab | -0.32 | -0.63 | -0.02 | **Moderate** |
| Mepolizumab | Reslizumab | 0.37 | -0.16 | 0.89 | **Moderate** |
| Benralizumab | Tezepelumab | -0.01 | -0.23 | 0.21 | **High** |
| Dupilumab | Tezepelumab | 0.03 | -0.23 | 0.28 | **High** |
| Mepolizumab | Tezepelumab | 0.72 | 0.22 | 1.22 | **Moderate** |
| Reslizumab | Tezepelumab | 0.35 | 0.1 | 0.6 | **Moderate** |

### eTable 10. Network estimates change in FEV1 (all patients)

| ***FEV1*** | ***Comparison*** | ***Network estimate*** |  | | |
| --- | --- | --- | --- | --- | --- |
| ***MCID 100 mL*** |  | **Relative estimate** | | |  |
| *Treatment 1* | Treatment 2 | Point estimate | CI Lower limit | CI upper limit | **Final rating** |
| *Tau^2=0.0020* | I^2=49.4% |  |  |  |  |
| Astegolimab | Benralizumab | -0.02 | -0.12 | 0.08 | Moderate |
| Astegolimab | Dupilumab | -0.14 | -0.25 | -0.03 | Moderate |
| Benralizumab | Dupilumab | -0.12 | -0.19 | -0.05 | Moderate |
| Astegolimab | Fevipiprant | 0.01 | -0.1 | 0.12 | Low |
| Benralizumab | Fevipiprant | 0.03 | -0.04 | 0.1 | Moderate |
| Dupilumab | Fevipiprant | 0.15 | 0.08 | 0.23 | Moderate |
| Astegolimab | Itepekimab | -0.11 | -0.28 | 0.06 | Moderate |
| Benralizumab | Itepekimab | -0.09 | -0.24 | 0.06 | Moderate |
| Dupilumab | Itepekimab | 0.03 | -0.11 | 0.18 | Low |
| Fevipiprant | Itepekimab | -0.12 | -0.27 | 0.04 | Moderate |
| Astegolimab | Itepekimab/dupilumab | -0.07 | -0.24 | 0.1 | Low |
| Benralizumab | Itepekimab/dupilumab | -0.05 | -0.2 | 0.1 | Low |
| Dupilumab | Itepekimab/dupilumab | 0.07 | -0.07 | 0.22 | Moderate |
| Fevipiprant | Itepekimab/dupilumab | -0.08 | -0.23 | 0.08 | Moderate |
| Itepekimab | Itepekimab/dupilumab | 0.04 | -0.12 | 0.2 | Low |
| Astegolimab | Mepolizumab | -0.05 | -0.17 | 0.06 | Moderate |
| Benralizumab | Mepolizumab | -0.03 | -0.11 | 0.04 | Moderate |
| Dupilumab | Mepolizumab | 0.09 | 0.01 | 0.17 | Moderate |
| Fevipiprant | Mepolizumab | -0.06 | -0.15 | 0.02 | Moderate |
| Itepekimab | Mepolizumab | 0.06 | -0.1 | 0.21 | Low |
| Itepekimab/dupilumab | Mepolizumab | 0.02 | -0.14 | 0.17 | Low |
| Astegolimab | Omalizumab | -0.04 | -0.16 | 0.08 | Moderate |
| Benralizumab | Omalizumab | -0.02 | -0.1 | 0.06 | Moderate |
| Dupilumab | Omalizumab | 0.1 | 0.01 | 0.19 | Moderate |
| Fevipiprant | Omalizumab | -0.05 | -0.14 | 0.04 | Moderate |
| Itepekimab | Omalizumab | 0.07 | -0.09 | 0.23 | Moderate |
| Itepekimab/dupilumab | Omalizumab | 0.03 | -0.13 | 0.19 | Low |
| Mepolizumab | Omalizumab | 0.01 | -0.08 | 0.11 | Moderate |
| Astegolimab | Placebo | 0.05 | -0.05 | 0.14 | Low |
| Benralizumab | Placebo | 0.07 | 0.02 | 0.11 | Moderate |
| Dupilumab | Placebo | 0.19 | 0.13 | 0.24 | High |
| Fevipiprant | Placebo | 0.03 | -0.02 | 0.09 | Moderate |
| Itepekimab | Placebo | 0.15 | 0.01 | 0.3 | Moderate |
| Itepekimab/dupilumab | Placebo | 0.11 | -0.03 | 0.26 | Moderate |
| Mepolizumab | Placebo | 0.1 | 0.04 | 0.16 | Moderate |
| Omalizumab | Placebo | 0.09 | 0.02 | 0.16 | Moderate |
| Reslizumab | Placebo | 0.13 | 0.07 | 0.18 | Moderate |
| Tezepelumab | Placebo | 0.16 | 0.09 | 0.23 | Moderate |
| Tralokinumab | Placebo | 0.14 | 0.05 | 0.23 | Moderate |
| Astegolimab | Reslizumab | -0.08 | -0.19 | 0.03 | Moderate |
| Benralizumab | Reslizumab | -0.06 | -0.13 | 0.01 | Moderate |
| Dupilumab | Reslizumab | 0.06 | -0.01 | 0.14 | Moderate |
| Fevipiprant | Reslizumab | -0.09 | -0.17 | -0.01 | Moderate |
| Itepekimab | Reslizumab | 0.03 | -0.13 | 0.18 | Low |
| Itepekimab/dupilumab | Reslizumab | -0.01 | -0.17 | 0.14 | Low |
| Mepolizumab | Reslizumab | -0.03 | -0.11 | 0.05 | Moderate |
| Omalizumab | Reslizumab | -0.04 | -0.13 | 0.05 | Moderate |
| Astegolimab | Tezepelumab | -0.12 | -0.23 | 0 | Moderate |
| Benralizumab | Tezepelumab | -0.1 | -0.18 | -0.01 | Moderate |
| Dupilumab | Tezepelumab | 0.03 | -0.06 | 0.11 | Moderate |
| Fevipiprant | Tezepelumab | -0.13 | -0.22 | -0.04 | Moderate |
| Itepekimab | Tezepelumab | -0.01 | -0.17 | 0.15 | Low |
| Itepekimab/dupilumab | Tezepelumab | -0.05 | -0.21 | 0.11 | Low |
| Mepolizumab | Tezepelumab | -0.06 | -0.16 | 0.03 | Moderate |
| Omalizumab | Tezepelumab | -0.08 | -0.17 | 0.02 | Moderate |
| Reslizumab | Tezepelumab | -0.04 | -0.12 | 0.05 | Moderate |
| Astegolimab | Tralokinumab | -0.09 | -0.22 | 0.04 | Moderate |
| Benralizumab | Tralokinumab | -0.07 | -0.17 | 0.03 | Moderate |
| Dupilumab | Tralokinumab | 0.05 | -0.06 | 0.15 | Moderate |
| Fevipiprant | Tralokinumab | -0.1 | -0.21 | 0 | Moderate |
| Itepekimab | Tralokinumab | 0.02 | -0.15 | 0.19 | Low |
| Itepekimab/dupilumab | Tralokinumab | -0.02 | -0.19 | 0.15 | Low |
| Mepolizumab | Tralokinumab | -0.04 | -0.15 | 0.07 | Moderate |
| Omalizumab | Tralokinumab | -0.05 | -0.17 | 0.06 | Moderate |
| Reslizumab | Tralokinumab | -0.01 | -0.12 | 0.09 | Moderate |
| Tezepelumab | Tralokinumab | 0.02 | -0.09 | 0.14 | Moderate |

### eTable 11. Network estimates change in FEV1 (eos >=300)

| ***FEV1*** | ***Comparison*** | ***Network estimate*** | | | |
| --- | --- | --- | --- | --- | --- |
| ***MCID 100 mL*** |  | **Relative estimate** | | | |
| *Treatment 1* | Treatment 2 | Point estimate | CI Lower limit | CI upper limit | **Final rating** |
| *Tau^2= 0.0008* | I^2=38.8% |  |  |  |  |
| Benralizumab | Dupilumab | -0.11 | -0.16 | -0.05 | **Moderate** |
| Benralizumab | Fevipiprant | 0.09 | 0.01 | 0.16 | **Moderate** |
| Dupilumab | Fevipiprant | 0.19 | 0.12 | 0.27 | **High** |
| Benralizumab | Itepekimab | -0.04 | -0.22 | 0.14 | **Low** |
| Dupilumab | Itepekimab | 0.07 | -0.11 | 0.25 | **Low** |
| Fevipiprant | Itepekimab | -0.13 | -0.32 | 0.06 | **Moderate** |
| Benralizumab | Itepekimab/dupilumab | -0.01 | -0.21 | 0.19 | **Low** |
| Dupilumab | Itepekimab/dupilumab | 0.1 | -0.1 | 0.29 | **Low** |
| Fevipiprant | Itepekimab/dupilumab | -0.1 | -0.3 | 0.11 | **Low** |
| Itepekimab | Itepekimab/dupilumab | 0.03 | -0.19 | 0.25 | **Low** |
| Benralizumab | Mepolizumab | 0.05 | -0.02 | 0.11 | **Moderate** |
| Dupilumab | Mepolizumab | 0.15 | 0.08 | 0.22 | **Moderate** |
| Fevipiprant | Mepolizumab | -0.04 | -0.13 | 0.05 | **Moderate** |
| Itepekimab | Mepolizumab | 0.08 | -0.1 | 0.27 | **Low** |
| Itepekimab/dupilumab | Mepolizumab | 0.05 | -0.15 | 0.26 | **Low** |
| Benralizumab | Placebo | 0.14 | 0.11 | 0.18 | **Moderate** |
| Dupilumab | Placebo | 0.25 | 0.21 | 0.29 | **High** |
| Fevipiprant | Placebo | 0.05 | -0.01 | 0.12 | **Moderate** |
| Itepekimab | Placebo | 0.18 | 0 | 0.36 | **Moderate** |
| Itepekimab/dupilumab | Placebo | 0.15 | -0.05 | 0.34 | **Low** |
| Mepolizumab | Placebo | 0.1 | 0.04 | 0.15 | **Moderate** |
| Reslizumab | Placebo | 0.19 | 0.12 | 0.25 | **High** |
| Tezepelumab | Placebo | 0.24 | 0.16 | 0.32 | **High** |
| Benralizumab | Reslizumab | -0.04 | -0.12 | 0.03 | **Moderate** |
| Dupilumab | Reslizumab | 0.06 | -0.01 | 0.14 | **Moderate** |
| Fevipiprant | Reslizumab | -0.13 | -0.22 | -0.04 | **Moderate** |
| Itepekimab | Reslizumab | -0.01 | -0.2 | 0.18 | **Low** |
| Itepekimab/dupilumab | Reslizumab | -0.04 | -0.24 | 0.17 | **Low** |
| Mepolizumab | Reslizumab | -0.09 | -0.17 | -0.01 | **Moderate** |
| Benralizumab | Tezepelumab | -0.1 | -0.19 | -0.01 | **Moderate** |
| Dupilumab | Tezepelumab | 0.01 | -0.08 | 0.1 | **Moderate** |
| Fevipiprant | Tezepelumab | -0.19 | -0.29 | -0.08 | **Moderate** |
| Itepekimab | Tezepelumab | -0.06 | -0.26 | 0.13 | **Low** |
| Itepekimab/dupilumab | Tezepelumab | -0.09 | -0.3 | 0.12 | **Low** |
| Mepolizumab | Tezepelumab | -0.15 | -0.24 | -0.05 | **Moderate** |
| Reslizumab | Tezepelumab | -0.06 | -0.16 | 0.05 | **Moderate** |

### eTable 12. Network estimates change in FEV1 (eos <300)

| ***FEV1*** | ***Comparison*** | ***Network estimate*** |  |  |  |
| --- | --- | --- | --- | --- | --- |
| ***MCID 100 mL*** |  | **Relative estimate** |  |  |  |
| *Treatment 1* | Treatment 2 | Point estimate | CI Lower limit | CI upper limit | **Final rating** |
| *Tau^2=0.0037* | I^2=54.9% |  |  |  |  |
| Benralizumab | Dupilumab | -0.06 | -0.2 | 0.08 | **Moderate** |
| Benralizumab | Itepekimab | -0.05 | -0.26 | 0.16 | **Low** |
| Dupilumab | Itepekimab | 0.01 | -0.18 | 0.19 | **Low** |
| Benralizumab | Itepekimab/Dupilumab | -0.01 | -0.22 | 0.2 | **Low** |
| Dupilumab | Itepekimab/Dupilumab | 0.05 | -0.14 | 0.23 | **Low** |
| Itepekimab | Itepekimab/Dupilumab | 0.04 | -0.16 | 0.24 | **Low** |
| Benralizumab | Placebo | 0.04 | -0.05 | 0.13 | **Moderate** |
| Dupilumab | Placebo | 0.1 | 0 | 0.2 | **Low** |
| Itepekimab | Placebo | 0.09 | -0.1 | 0.28 | **Low** |
| Itepekimab/Dupilumab | Placebo | 0.05 | -0.14 | 0.24 | **Low** |
| Reslizumab | Placebo | 0.09 | -0.04 | 0.22 | **Low** |
| Tezepelumab | Placebo | 0.1 | 0 | 0.19 | **Low** |
| Benralizumab | Reslizumab | -0.05 | -0.21 | 0.11 | **Low** |
| Dupilumab | Reslizumab | 0.01 | -0.15 | 0.18 | **Low** |
| Itepekimab | Reslizumab | 0 | -0.23 | 0.23 | **Low** |
| Itepekimab/Dupilumab | Reslizumab | -0.04 | -0.27 | 0.19 | **Low** |
| Benralizumab | Tezepelumab | -0.06 | -0.19 | 0.08 | **Moderate** |
| Dupilumab | Tezepelumab | 0 | -0.14 | 0.15 | **Low** |
| Itepekimab | Tezepelumab | 0 | -0.22 | 0.21 | **Low** |
| Itepekimab/Dupilumab | Tezepelumab | -0.04 | -0.26 | 0.17 | **Low** |
| Reslizumab | Tezepelumab | -0.01 | -0.17 | 0.16 | **Low** |

### eTable 13. Network estimates hospital admissions

| ***Hospitalizations*** | ***Comparison*** | ***Network estimate*** | | | ***Network estimate*** | | | |
| --- | --- | --- | --- | --- | --- | --- | --- | --- |
| ***MID 5%*** | **Basline risk 13.7%** | **Relative estimate** | | | **Absolute risk difference per 1000** | | | |
| *Treatment 1* | Treatment 2 | Point estimate | CI Lower limit | CI upper limit | Point estimate | CI Lower limit | CI upper limit | **Final rating** |
| *Tau^2=0.0197* | I^2=25.2% |  |  |  |  |  |  |  |
| Benralizumab | Mepolizumab | 2.98 | 0.88 | 10.14 | 235.9962 | -14.3028 | 1089.3966 | **Moderate** |
| Benralizumab | Omalizumab | 2.27 | 1.28 | 4.03 | 66.1162 | 14.5768 | 157.7418 | **Moderate** |
| Mepolizumab | Omalizumab | 0.76 | 0.2 | 2.84 | -12.4944 | -41.648 | 95.7904 | **Moderate** |
| Benralizumab | Placebo | 0.87 | 0.7 | 1.08 | -17.81 | -41.1 | 10.96 | **High** |
| Mepolizumab | Placebo | 0.29 | 0.09 | 0.97 | -97.27 | -124.67 | -4.11 | **Moderate** |
| Omalizumab | Placebo | 0.38 | 0.23 | 0.65 | -84.94 | -105.49 | -47.95 | **Moderate** |
| Tezepelumab | Placebo | 0.19 | 0.12 | 0.31 | -110.97 | -120.56 | -94.53 | **Moderate** |
| Tralokinumab | Placebo | 0.76 | 0.53 | 1.09 | -32.88 | -64.39 | 12.33 | **Moderate** |
| Benralizumab | Tezepelumab | 4.52 | 2.67 | 7.64 | 91.6256 | 43.4701 | 172.8392 | **Moderate** |
| Mepolizumab | Tezepelumab | 1.51 | 0.41 | 5.53 | 13.2753 | -15.3577 | 117.9159 | **Low** |
| Omalizumab | Tezepelumab | 1.99 | 0.97 | 4.05 | 25.7697 | -0.7809 | 79.3915 | **Moderate** |
| Benralizumab | Tralokinumab | 1.14 | 0.75 | 1.73 | 14.5768 | -26.03 | 76.0076 | **Moderate** |
| Mepolizumab | Tralokinumab | 0.38 | 0.11 | 1.34 | -64.5544 | -92.6668 | 35.4008 | **Low** |
| Omalizumab | Tralokinumab | 0.5 | 0.27 | 0.95 | -52.06 | -76.0076 | -5.206 | **Moderate** |
| Tezepelumab | Tralokinumab | 0.25 | 0.14 | 0.46 | -78.09 | -89.5432 | -56.2248 | **High** |

### eTable 14. Network estimates Corticosteroid sparing

| ***Corticosteroid sparing*** | ***Comparison*** | ***Network estimate*** | | | ***Network estimate*** | | | |
| --- | --- | --- | --- | --- | --- | --- | --- | --- |
| ***Baseline risk 56%*** | **MID 20%** | **Relative estimate** | | | **Absolute risk difference per 1000** | | | |
| *Treatment 1* | Treatment 2 | Point estimate | CI Lower limit | CI upper limit | Point estimate | CI Lower limit | CI upper limit | **Final rating** |
| *Tau^2=0* | I^2=0 |  |  |  |  |  |  |  |
| Benralizumab | Dupilumab | 1.19 | 0.82 | 1.73 | 158.536 | -150.192 | 609.112 | **Low** |
| Benralizumab | Mepolizumab | 1.1 | 0.66 | 1.84 | 99.12 | -337.008 | 832.608 | **Low** |
| Dupilumab | Mepolizumab | 0.93 | 0.59 | 1.46 | -69.384 | -406.392 | 455.952 | **Low** |
| Benralizumab | Omalizumab | 1.34 | 0.95 | 1.89 | 251.328 | -36.96 | 657.888 | **Low** |
| Dupilumab | Omalizumab | 1.13 | 0.89 | 1.44 | 96.096 | -81.312 | 325.248 | **Moderate** |
| Mepolizumab | Omalizumab | 1.22 | 0.8 | 1.87 | 162.624 | -147.84 | 643.104 | **Low** |
| Benralizumab | Placebo | 1.77 | 1.29 | 2.43 | 431.2 | 162.4 | 800.8 | **Moderate** |
| Dupilumab | Placebo | 1.49 | 1.22 | 1.83 | 274.4 | 123.2 | 464.8 | **Moderate** |
| Mepolizumab | Placebo | 1.61 | 1.07 | 2.41 | 341.6 | 39.2 | 789.6 | **Moderate** |
| Omalizumab | Placebo | 1.32 | 1.15 | 1.51 | 179.2 | 84 | 285.6 | **Moderate** |
| Resilizumab | Placebo | 1.23 | 0.85 | 1.78 | 128.8 | -84 | 436.8 | **low** |
| Tezepelumab | Placebo | 1.06 | 0.87 | 1.3 | 33.6 | -72.8 | 168 | **Low** |
| Benralizumab | Resilizumab | 1.44 | 0.89 | 2.33 | 303.072 | -75.768 | 916.104 | **Low** |
| Dupilumab | Resilizumab | 1.21 | 0.8 | 1.84 | 144.648 | -137.76 | 578.592 | **Low** |
| Mepolizumab | Resilizumab | 1.31 | 0.76 | 2.25 | 213.528 | -165.312 | 861 | **Low** |
| Omalizumab | Resilizumab | 1.07 | 0.73 | 1.58 | 48.216 | -185.976 | 399.504 | **Low** |
| Benralizumab | Tezepelumab | 1.66 | 1.15 | 2.42 | 391.776 | 89.04 | 842.912 | **Moderate** |
| Dupilumab | Tezepelumab | 1.4 | 1.06 | 1.86 | 237.44 | 35.616 | 510.496 | **Moderate** |
| Mepolizumab | Tezepelumab | 1.51 | 0.96 | 2.37 | 302.736 | -23.744 | 813.232 | **Low** |
| Omalizumab | Tezepelumab | 1.24 | 0.97 | 1.57 | 142.464 | -17.808 | 338.352 | **Moderate** |
| Resilizumab | Tezepelumab | 1.16 | 0.76 | 1.75 | 94.976 | -142.464 | 445.2 | **Moderate** |

### eTable 15. Adverse events leading to drug discontinuation

| ***ADE*** | ***Comparison*** | ***Network estimate*** | | | ***Network estimate*** | | | |
| --- | --- | --- | --- | --- | --- | --- | --- | --- |
| ***Baseline risk 1.90%*** | **MID 10%** | **Relative estimate** | | | **Absolute risk difference per 1000** | | | |
| *Treatment 1* | Treatment 2 | Point estimate | CI Lower limit | CI upper limit | Point estimate | CI Lower limit | CI upper limit | **Final rating** |
| *I^2=5.7%* | Tau^2=0.042 |  |  |  |  |  |  |  |
| Benralizumab | Dupliumab | 1.61 | 0.54 | 4.77 | 6.6063 | -4.9818 | 40.8291 | **High** |
| Benralizumab | Fevipiprant | 1.63 | 0.69 | 3.84 | 31.2797 | 13.2411 | 73.6896 | **Moderate** |
| Dupliumab | Fevipiprant | 1.02 | 0.41 | 2.54 | 19.5738 | 7.8679 | 48.7426 | **High** |
| Benralizumab | Mepolizumab | 2.56 | 1 | 6.56 | 19.266 | 0 | 68.666 | **Moderate** |
| Dupliumab | Mepolizumab | 1.6 | 0.59 | 4.31 | 7.41 | -5.0635 | 40.8785 | **High** |
| Fevipiprant | Mepolizumab | 1.57 | 0.75 | 3.27 | 7.0395 | -3.0875 | 28.0345 | **High** |
| Benralizumab | Omalizumab | 1.38 | 0.59 | 3.2 | 8.664 | -9.348 | 50.16 | **Low** |
| Dupliumab | Omalizumab | 0.86 | 0.35 | 2.11 | -3.192 | -14.82 | 25.308 | **Moderate** |
| Fevipiprant | Omalizumab | 0.84 | 0.46 | 1.54 | -3.648 | -12.312 | 12.312 | **Moderate** |
| Mepolizumab | Omalizumab | 0.54 | 0.26 | 1.1 | -10.488 | -16.872 | 2.28 | **Moderate** |
| Benralizumab | Placebo | 1.65 | 0.79 | 3.45 | 12.35 | -3.99 | 46.55 | **High** |
| Dupliumab | Placebo | 1.03 | 0.46 | 2.3 | 0.57 | -10.26 | 24.7 | **High** |
| Fevipiprant | Placebo | 1.01 | 0.65 | 1.57 | 0.19 | -6.65 | 10.83 | **High** |
| Mepolizumab | Placebo | 0.65 | 0.36 | 1.16 | -6.65 | -12.16 | 3.04 | **High** |
| Reslizumab | Placebo | 0.65 | 0.41 | 1.02 | -6.65 | -11.21 | 0.38 | **High** |
| Tezepelumab | Placebo | 0.68 | 0.34 | 1.35 | -6.08 | -12.54 | 6.65 | **High** |
| Tralokinumab | Placebo | 2.7 | 1.45 | 5 | 32.3 | 8.55 | 76 | **High** |
| Omalizumab | Placebo | 1.2 | 0.8 | 1.81 | 3.8 | -3.8 | 15.39 | **Moderate** |
| Benralizumab | Reslizumab | 2.55 | 1.07 | 6.06 | 19.1425 | 0.8645 | 62.491 | **Moderate** |
| Omalizumab | Reslizumab | 1.85 | 1 | 3.42 | 10.4975 | 0 | 29.887 | **Moderate** |
| Dupliumab | Reslizumab | 1.59 | 0.63 | 3.99 | 7.2865 | -4.5695 | 36.9265 | **High** |
| Fevipiprant | Reslizumab | 1.56 | 0.83 | 2.94 | 6.916 | -2.0995 | 23.959 | **High** |
| Mepolizumab | Reslizumab | 1 | 0.47 | 2.09 | 0 | -6.5455 | 13.4615 | **High** |
| Omalizumab | Tezepelumab | 1.77 | 0.79 | 3.93 | 1.2705 | -0.3465 | 4.8345 | **Moderate** |
| Benralizumab | Tezepelumab | 2.43 | 0.89 | 6.65 | 2.3595 | -0.1815 | 9.3225 | **High** |
| Dupliumab | Tezepelumab | 1.51 | 0.53 | 4.35 | 0.8415 | -0.7755 | 5.5275 | **High** |
| Fevipiprant | Tezepelumab | 1.49 | 0.66 | 3.37 | 0.8085 | -0.561 | 3.9105 | **High** |
| Mepolizumab | Tezepelumab | 0.95 | 0.38 | 2.34 | -0.0825 | -1.023 | 2.211 | **High** |
| Reslizumab | Tezepelumab | 0.95 | 0.42 | 2.17 | -0.0825 | -0.957 | 1.9305 | **High** |
| Omalizumab | Tralokinumab | 0.44 | 0.21 | 0.94 | -28.728 | -40.527 | -3.078 | **Moderate** |
| Benralizumab | Tralokinumab | 0.61 | 0.23 | 1.6 | -20.007 | -39.501 | 30.78 | **High** |
| Dupliumab | Tralokinumab | 0.38 | 0.14 | 1.05 | -31.806 | -44.118 | 2.565 | **High** |
| Fevipiprant | Tralokinumab | 0.37 | 0.17 | 0.81 | -32.319 | -42.579 | -9.747 | **High** |
| Mepolizumab | Tralokinumab | 0.24 | 0.1 | 0.56 | -38.988 | -46.17 | -22.572 | **High** |
| Reslizumab | Tralokinumab | 0.24 | 0.11 | 0.52 | -38.988 | -45.657 | -24.624 | **High** |
| Tezepelumab | Tralokinumab | 0.25 | 0.1 | 0.64 | -38.475 | -46.17 | -18.468 | **High** |

### eTable 16. Meta-regression for change in FEV1

| **Drug** | **Independent variable** | **p value** |
| --- | --- | --- |
| *Tezepelumab* |  |  |
|  | *age* | *0.72* |
|  | *sex* | *0.52* |
|  | *follow-up* | *0.7* |
|  | *Year of publication* | *0.7* |
| *Dupilumab* |  |  |
|  | *age* | *<0.0001* |
|  | *sex* | *<0.0001* |
|  | *follow-up* | *0.14* |
|  | *time from diagnosis* | *0.91* |
|  | *year of publication* | *0.03* |
|  | *FEV1* | *0.11* |
|  | *oral steroids* | *0.46* |
| *Mepolizumab* |  |  |
|  | *age* | *0.91* |
|  | *sex* | *0.49* |
|  | *follow-up* | *0.52* |
|  | *time from diagnosis* | *0.76* |
|  | *year of publication* | *0.73* |
|  | *FEV1* | *0.9* |
|  | *oral steroids* | *0.46* |
| *Reslizumab* |  |  |
|  | *age* | *0.72* |
|  | *sex* | *0.09* |
|  | *follow-up* | *0.7* |
|  | *time from diagnosis* | *0.99* |
|  | *year of publication* | *0.51* |
|  | *FEV1* | *0.76* |
|  | *oral steroids* | *0.79* |
| *Benralizumab* |  |  |
|  | *age* | *0.26* |
|  | *sex* | *0.23* |
|  | *follow-up* | *0.43* |
|  | *time from diagnosis* | *0.41* |
|  | *year of publication* | *0.04* |
|  | *FEV1* | *0.76* |
|  | *oral steroids* | *0.82* |
| *Omalizumab* |  |  |
|  | *age* | *0.57* |
|  | *sex* | *0.62* |
|  | *follow-up* | *0.85* |
|  | *time from diagnosis* | *0.41* |
|  | *year of publication* | *0.94* |
|  | *FEV1* | *0.61* |
|  | *oral steroids* | *0.82* |
| *Fevipiprant* |  |  |
|  | *age* | *0.07* |
|  | *sex* | *0.4* |
|  | *follow-up* | *0.19* |
|  | *time from diagnosis* | *0.07* |
|  | *year of publication* | *0.24* |
|  | *FEV1* | *0.92* |

### eFigure 1. PRISMA flow diagram for inclusion and exclusion

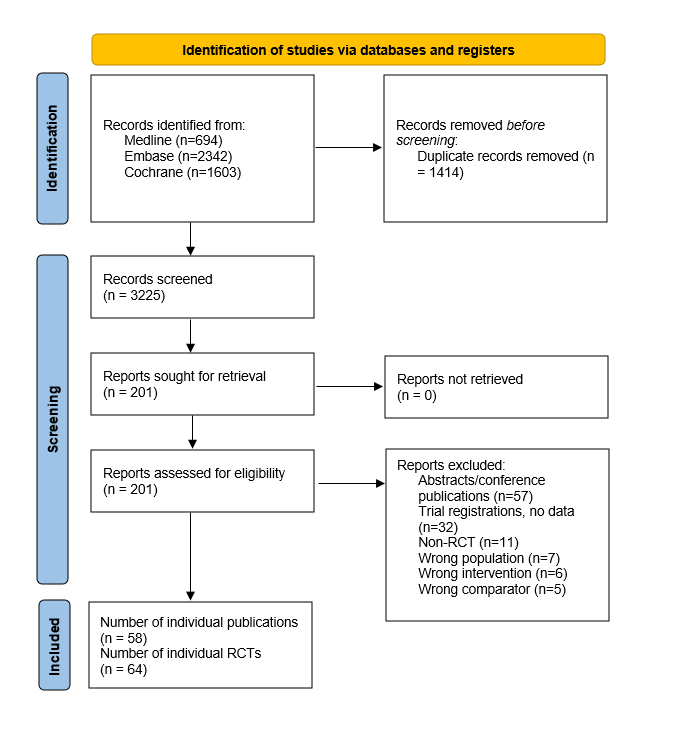

### eFigure 2. Exacerbations (all) Network diagram

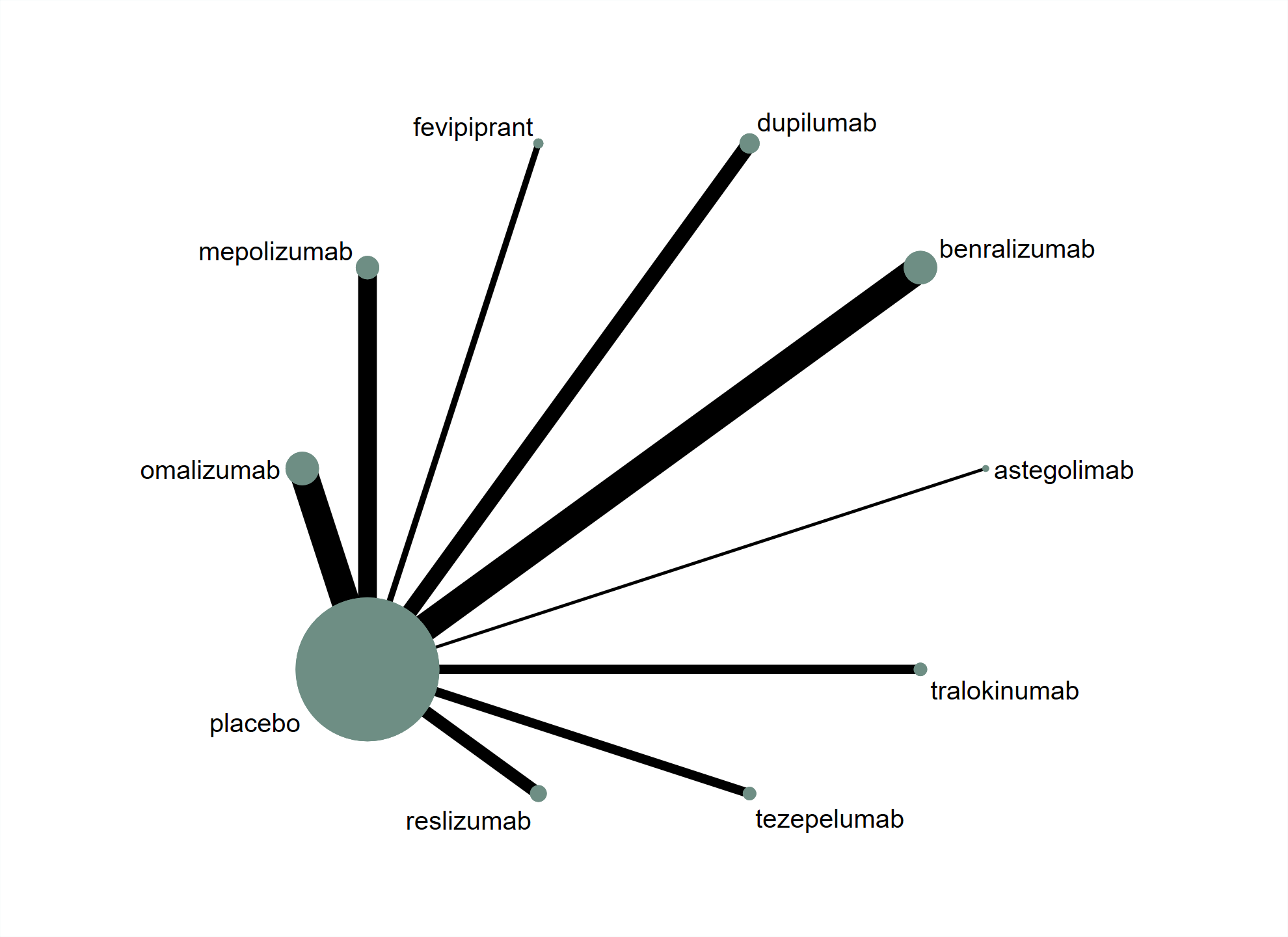

### eFigure 3. Exacerbations (all) network forest plot

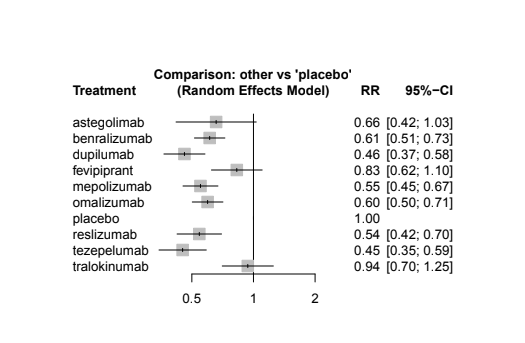

### eFigure 4. Exacerbations (all) pairwise forest plot

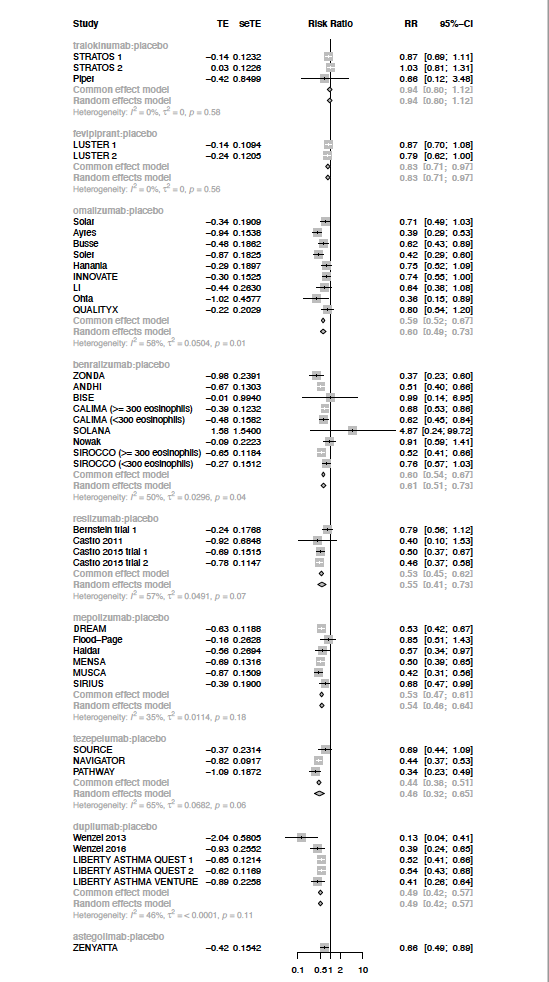

### eFigure 5. Exacerbations (all) funnel plot

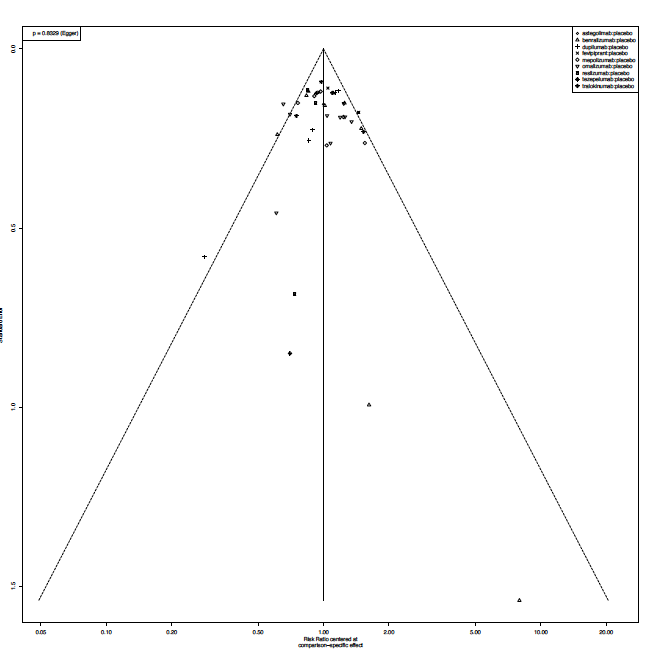

### eFigure 6: Exacerbations (high eos) network diagram

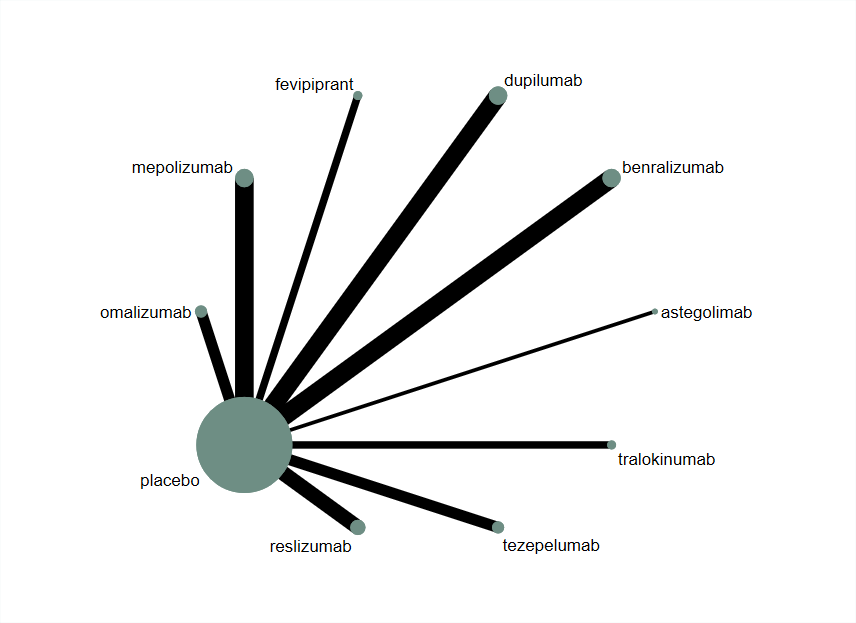

### eFigure 7. Exacerbations (high eos) network forest plot

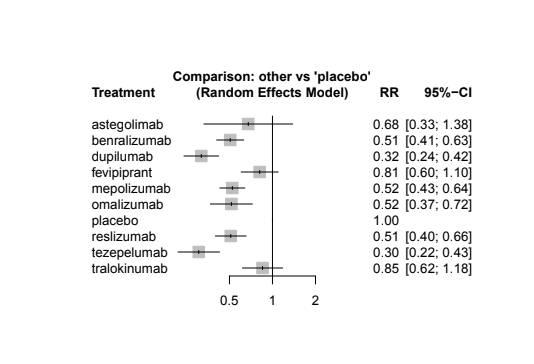

### eFigure 8. Exacerbations (high eos) pairwise forest plot

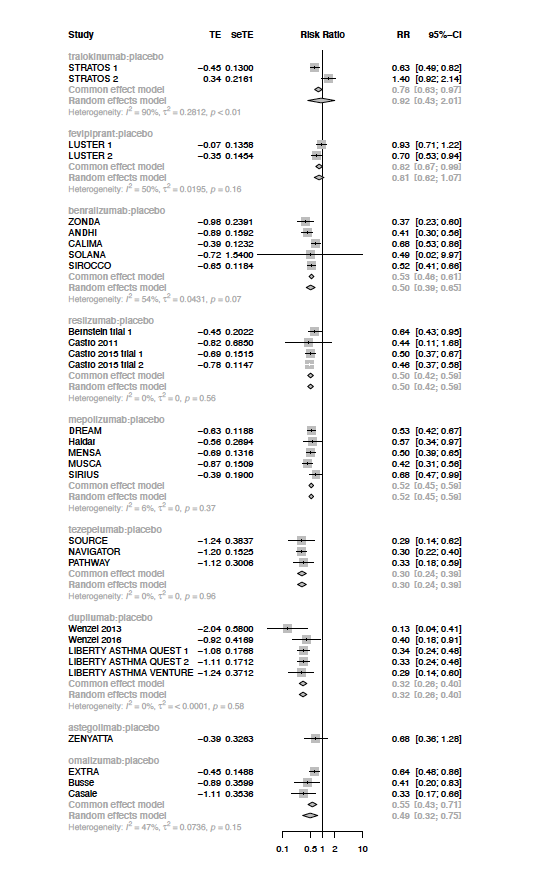

### eFigure 9. Exacerbations (High eos) funnel plot

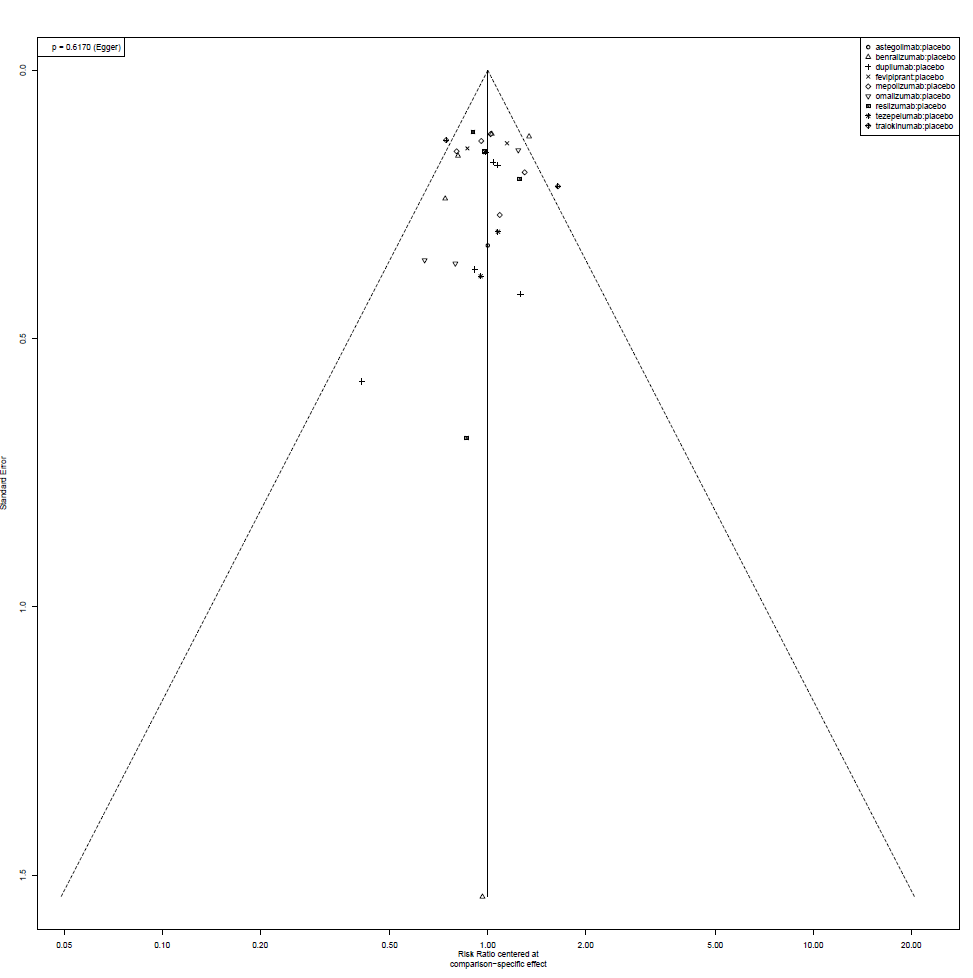

### eFigure 10. Exacerbations (Low eos) network diagram

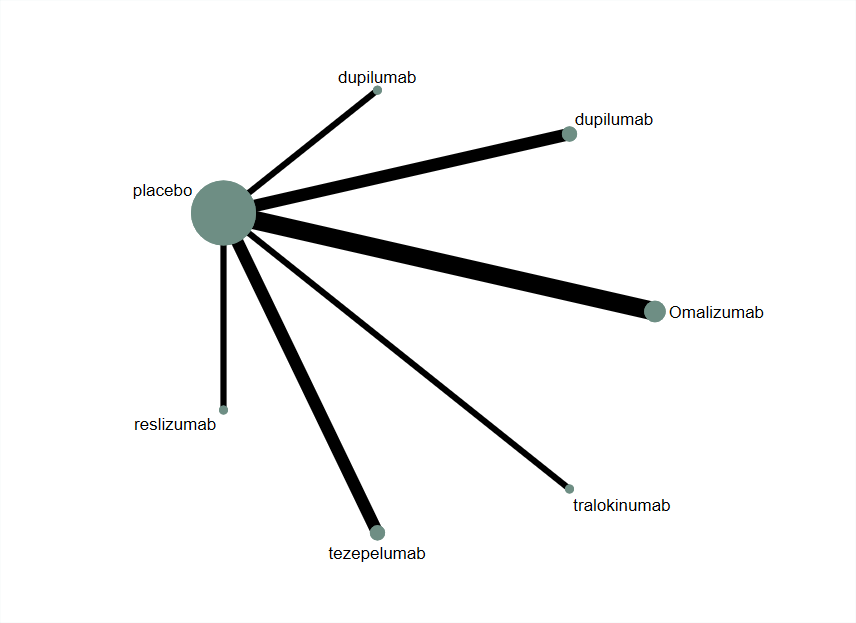

### eFigure 11. Exacerbations (low eos) network forest plot

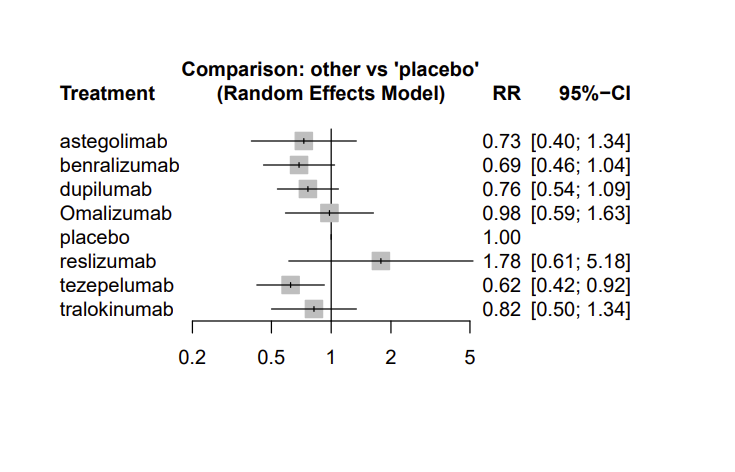

### eFigure 12. Exacerbations (low eos) pairwise forest plot

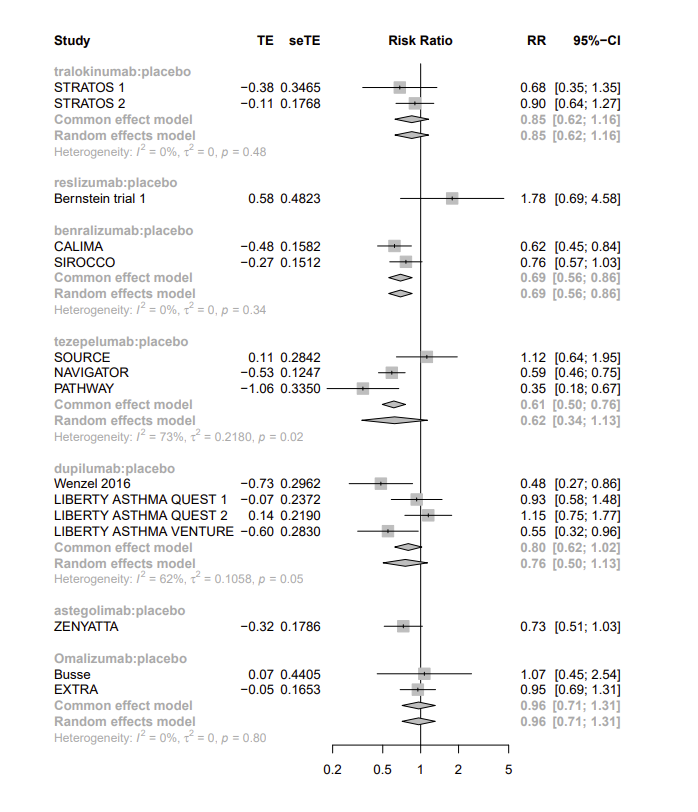

### eFigure 13. Exacerbations (low eos) funnel plot

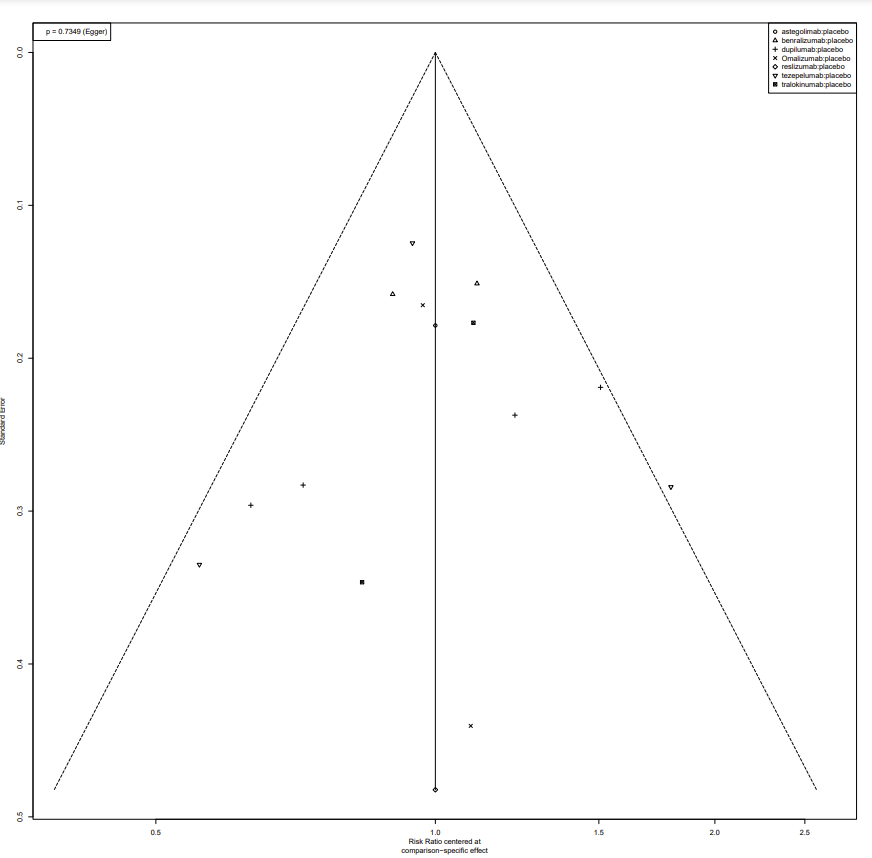

### eFigure 14. ACQ (all) network diagram

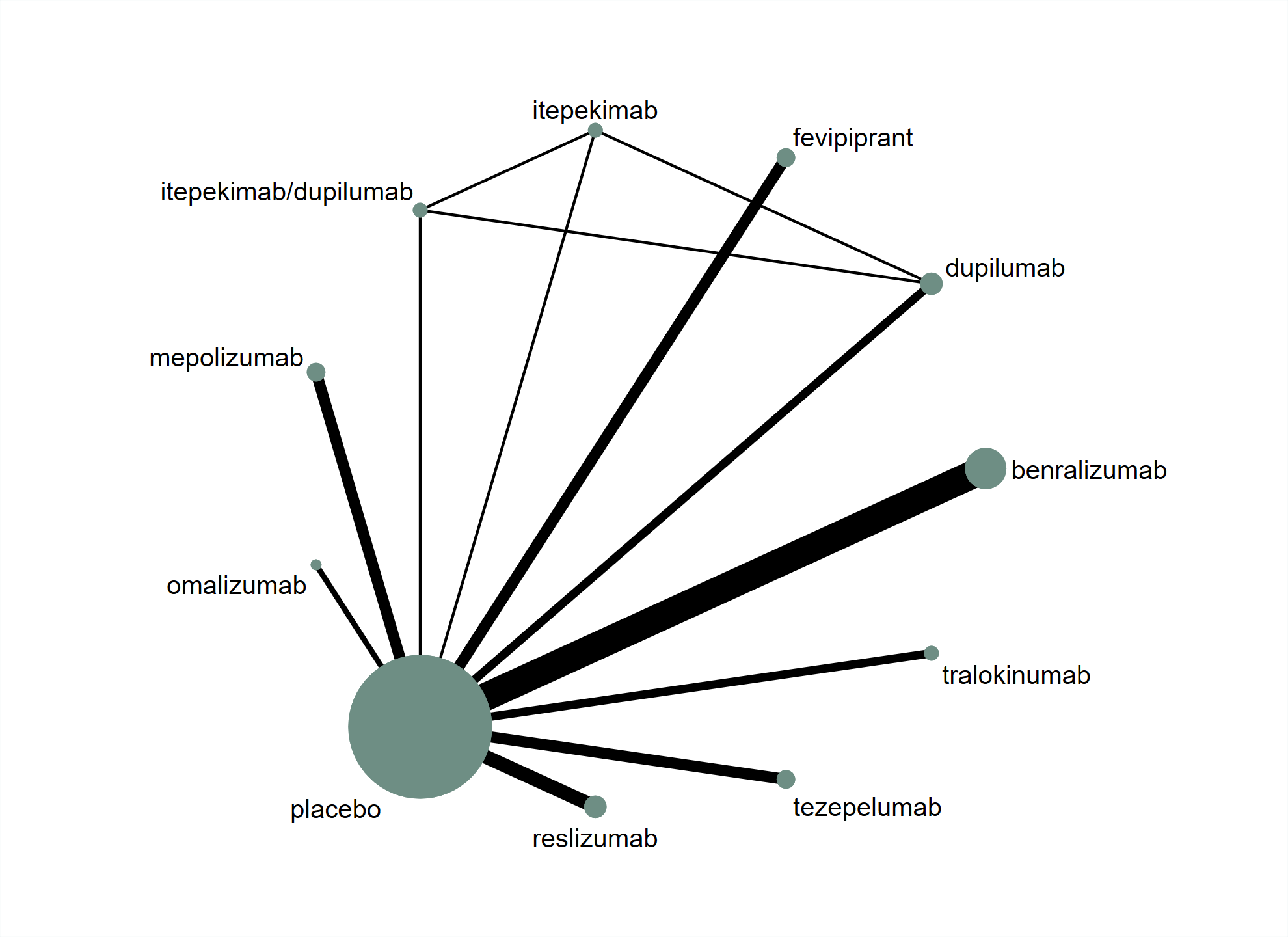

### eFigure 15. ACQ (all) network forest plot

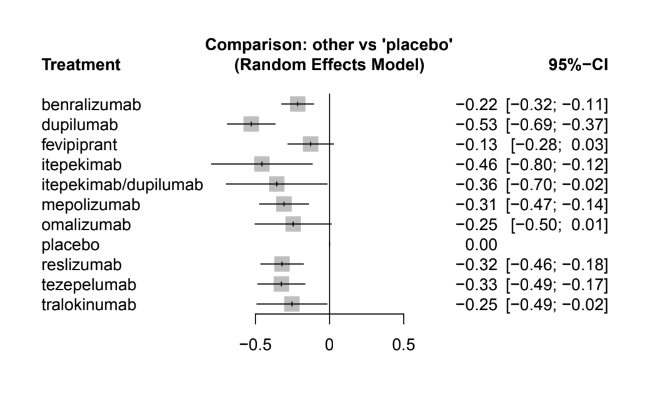

### eFigure 16. ACQ (all) pairwise forest plot

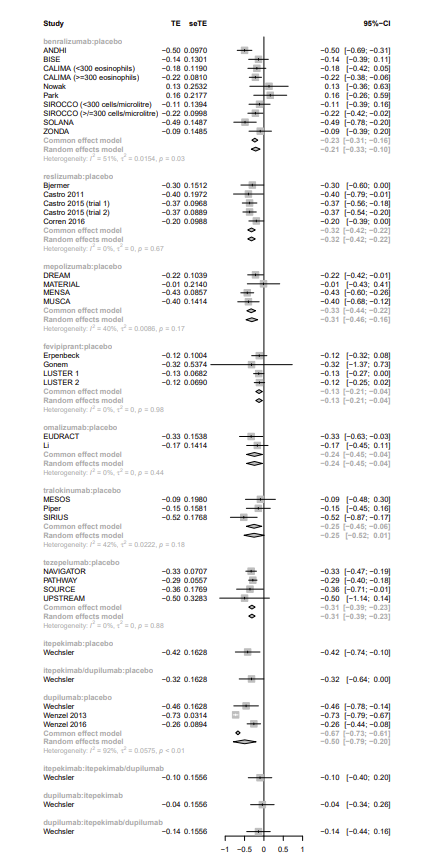

### eFigure 17. ACQ (all) funnel plots

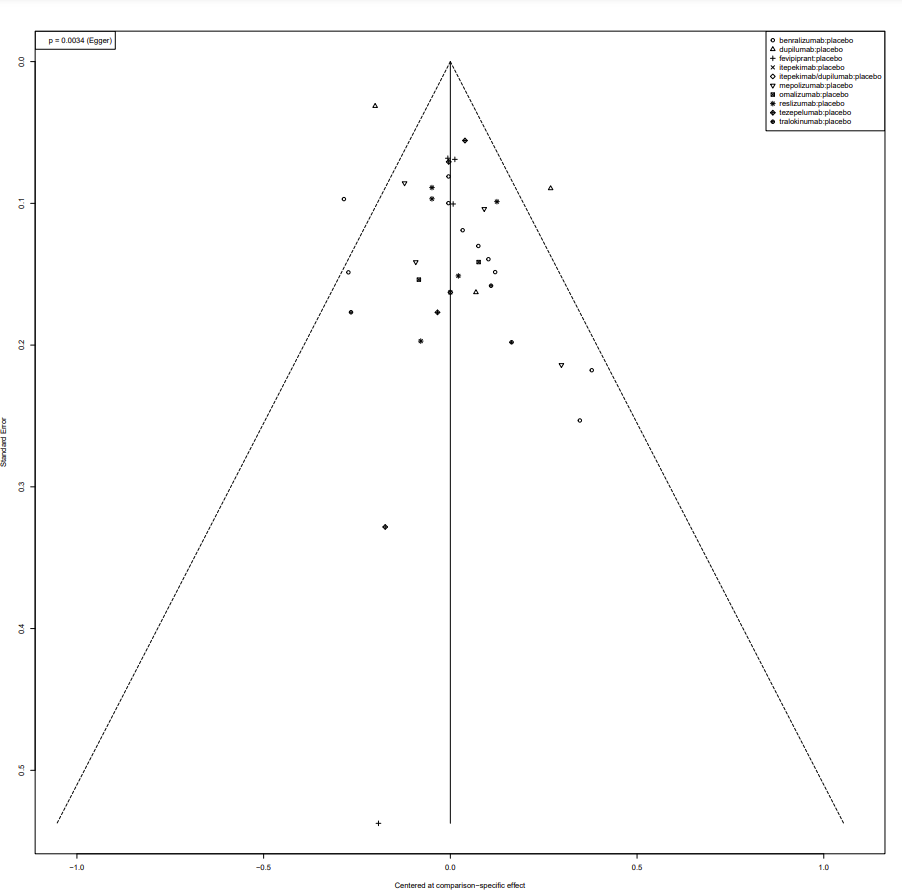

### eFigure 18. ACQ (high eos) network diagram

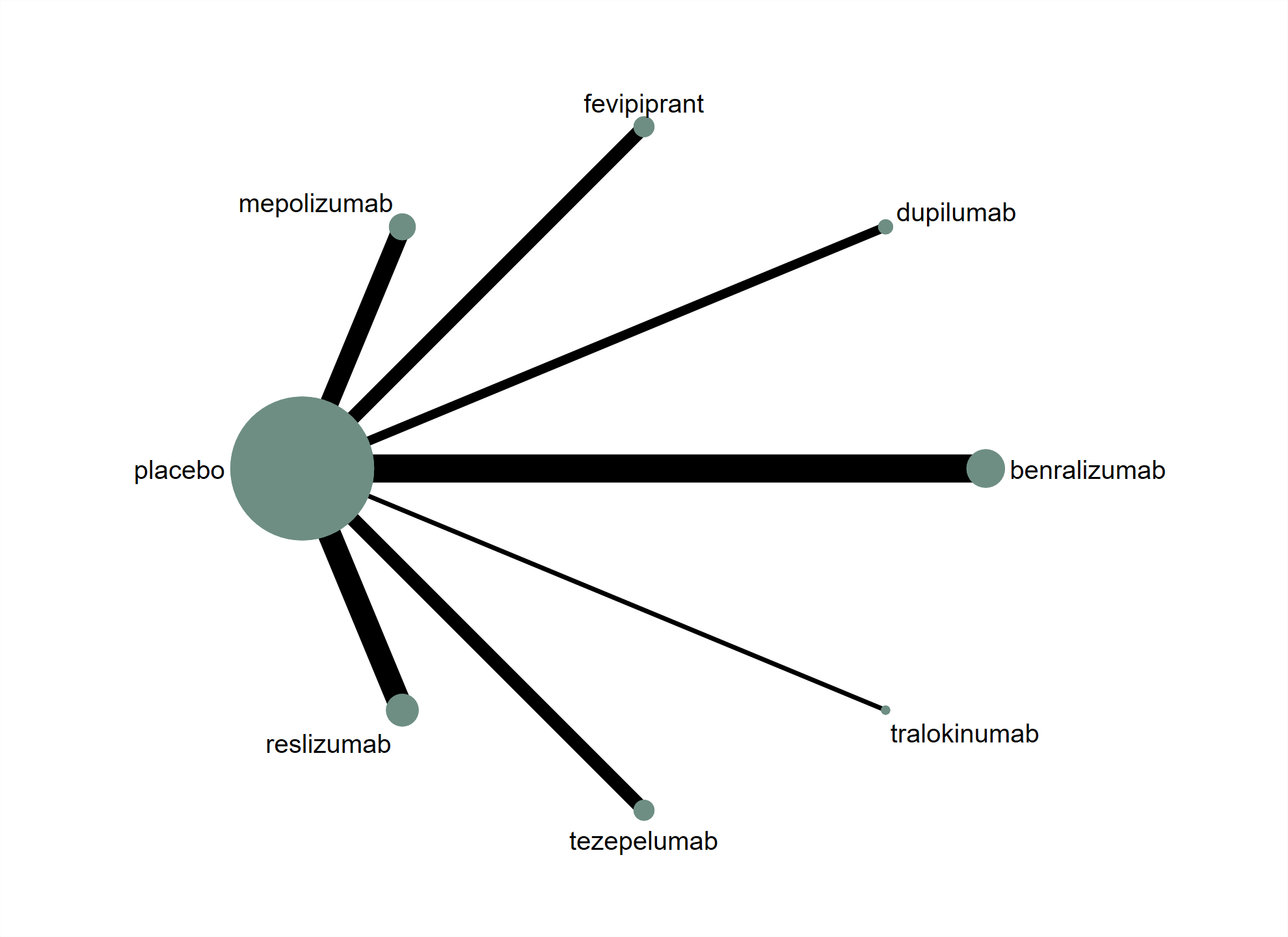

### eFigure 19. ACQ (high eos) network forest plot

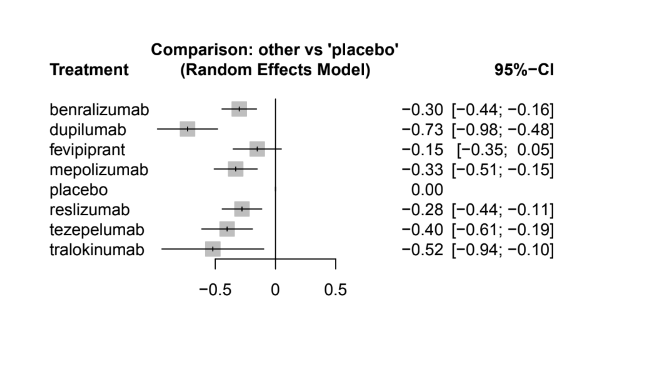

### eFigure 20. ACQ (high eos) pairwise forest plot

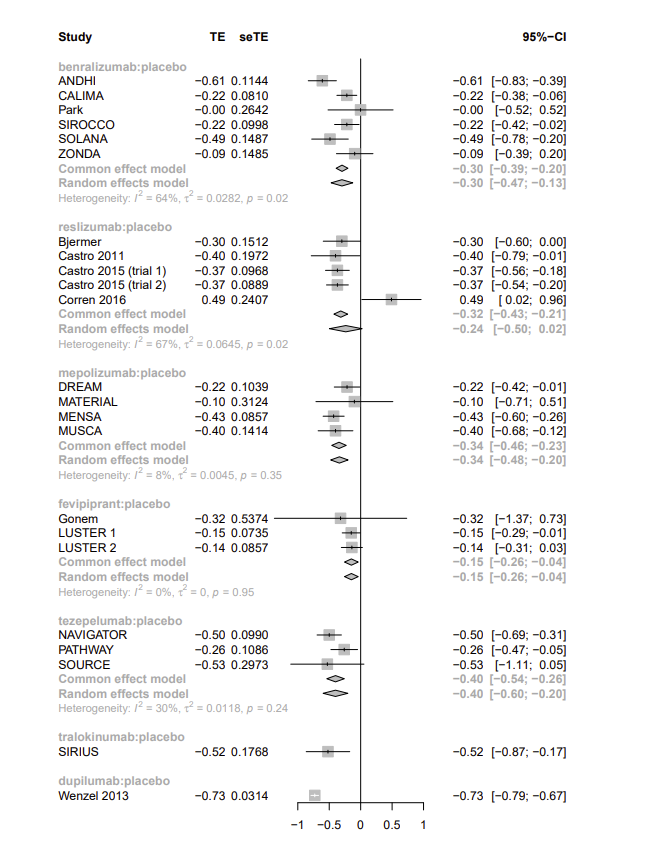

### eFigure 21. ACQ (high eos) funnel plot

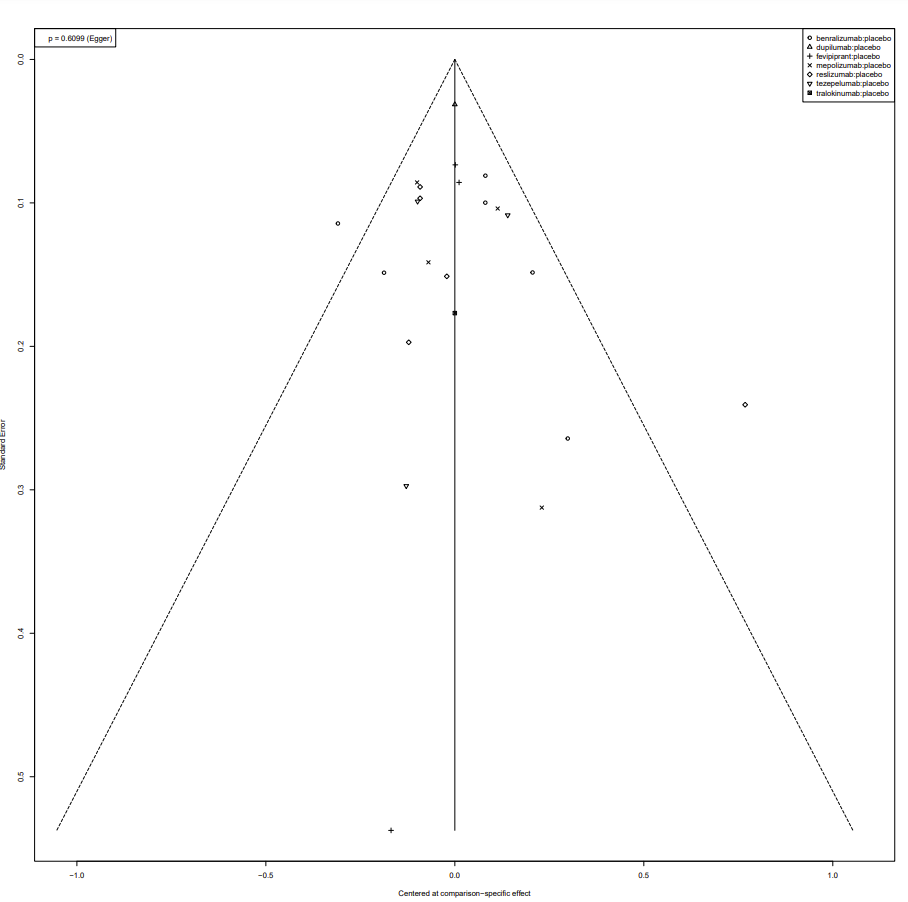

### eFigure 22. ACQ (low eos) network diagram

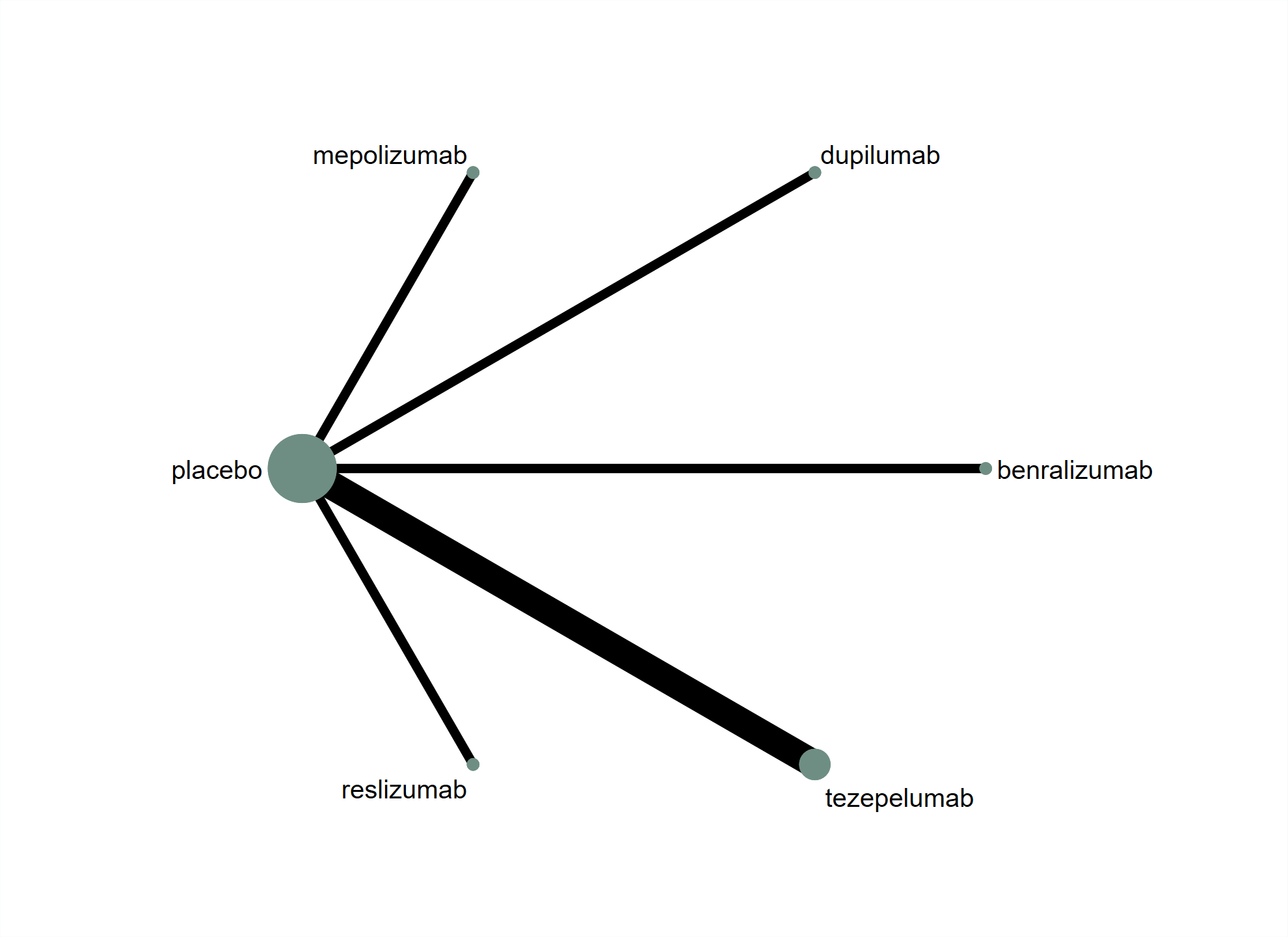

### eFigure 23. ACQ (low eos) network forest plot

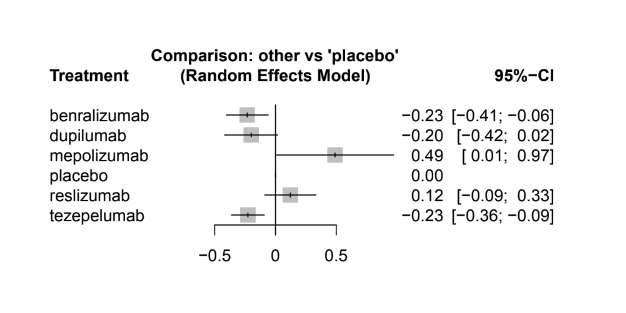

### eFigure 24. ACQ (low eos) pairwise forest plot

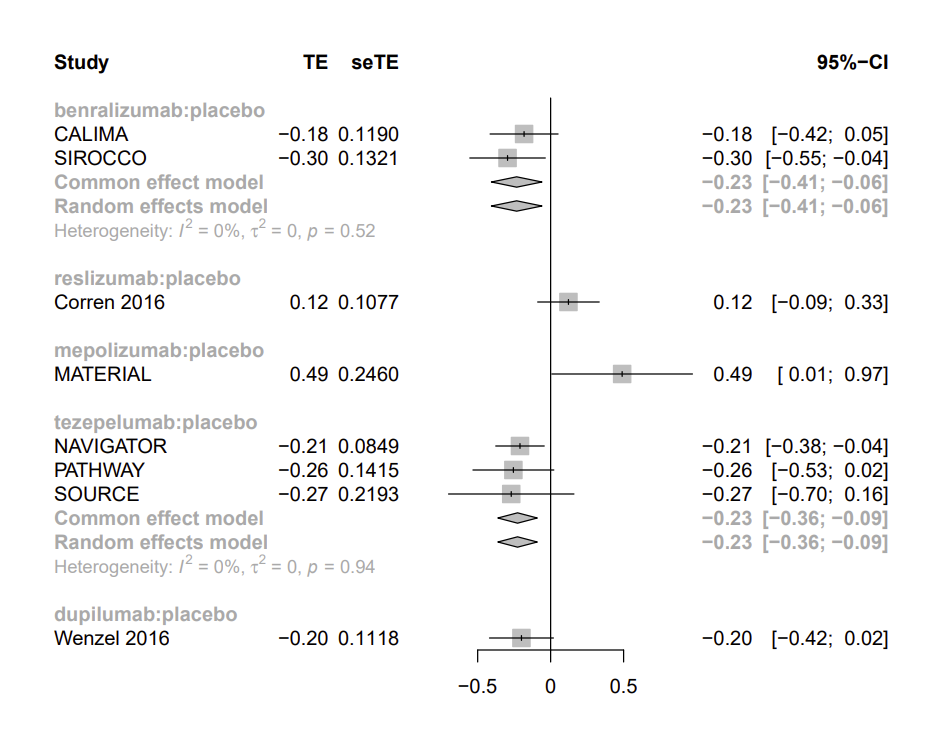

### eFigure 25. ACQ (low eos) funnel plot

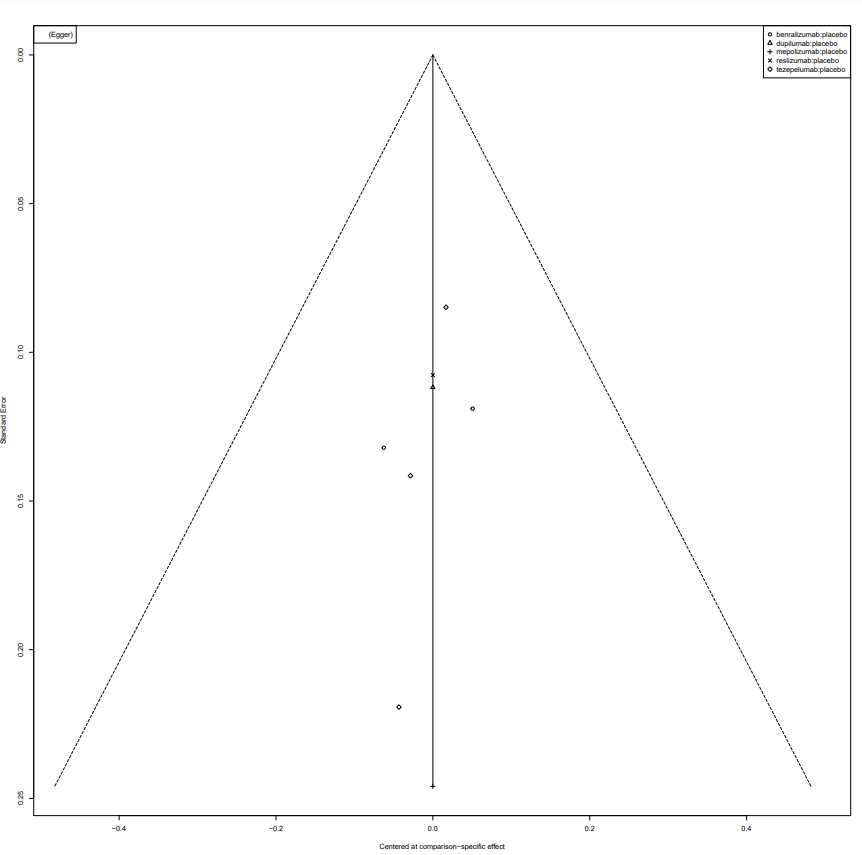

### eFigure 26. FEV1 (all) network diagram

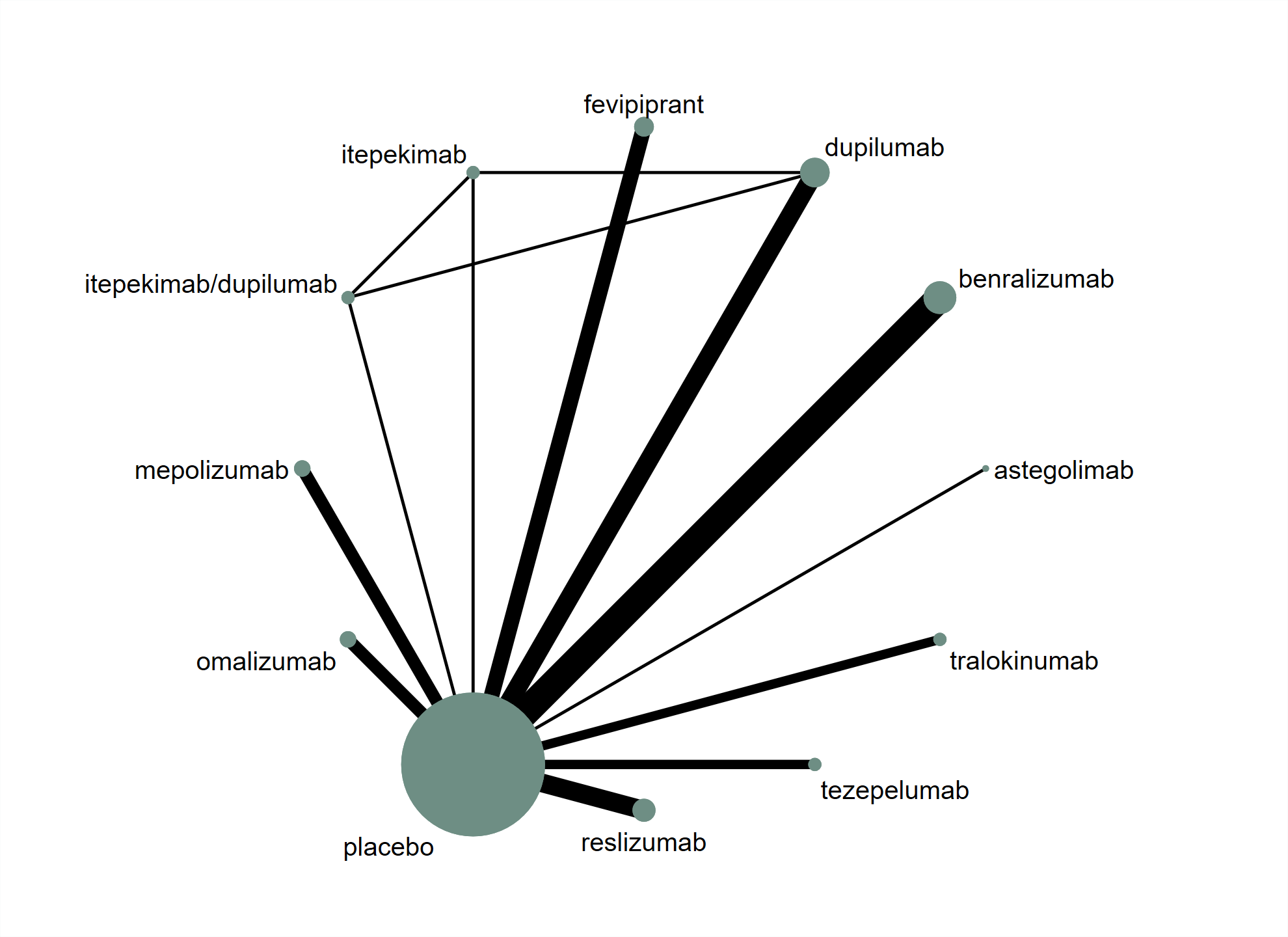

### eFigure 27. FEV1 (all) network forest plot

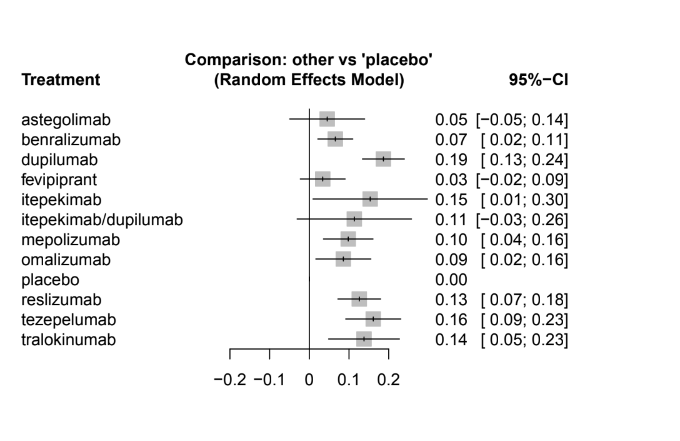

### eFigure 28. FEV1 (all) pairwise forest plot

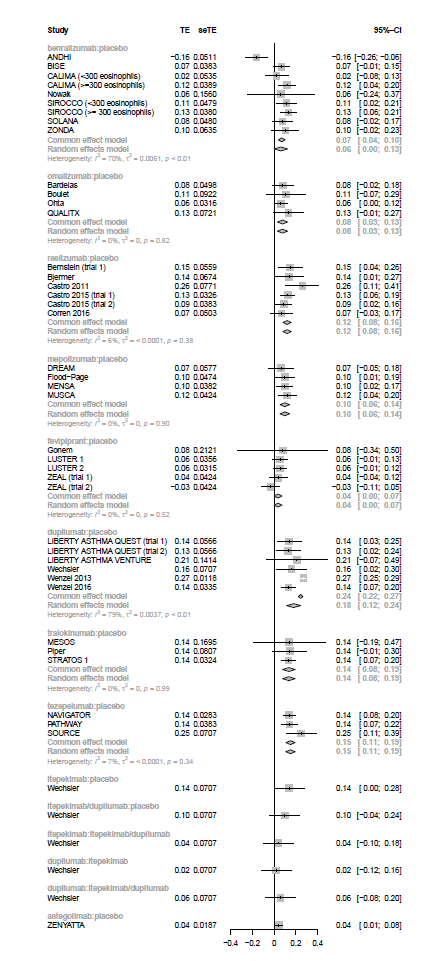

### eFigure 29. FEV1 (all) funnel plot

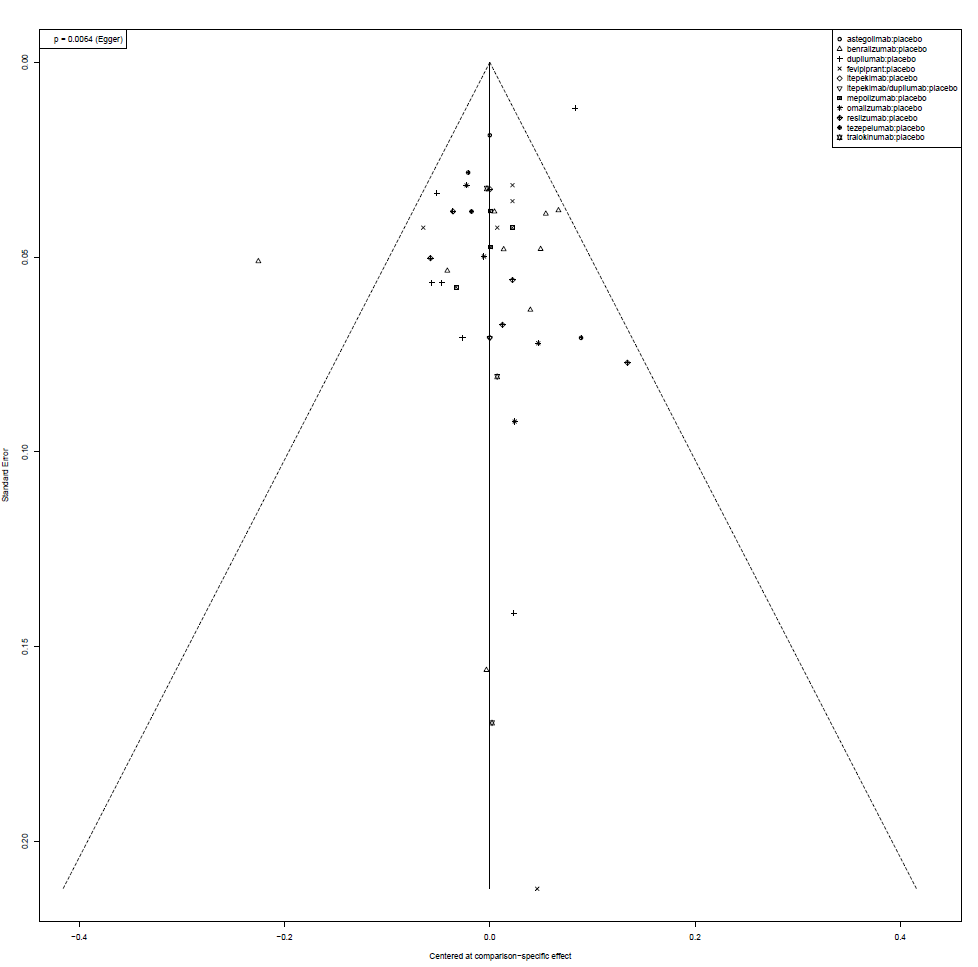

### eFigure 30. FEV1 (high eos) network diagram

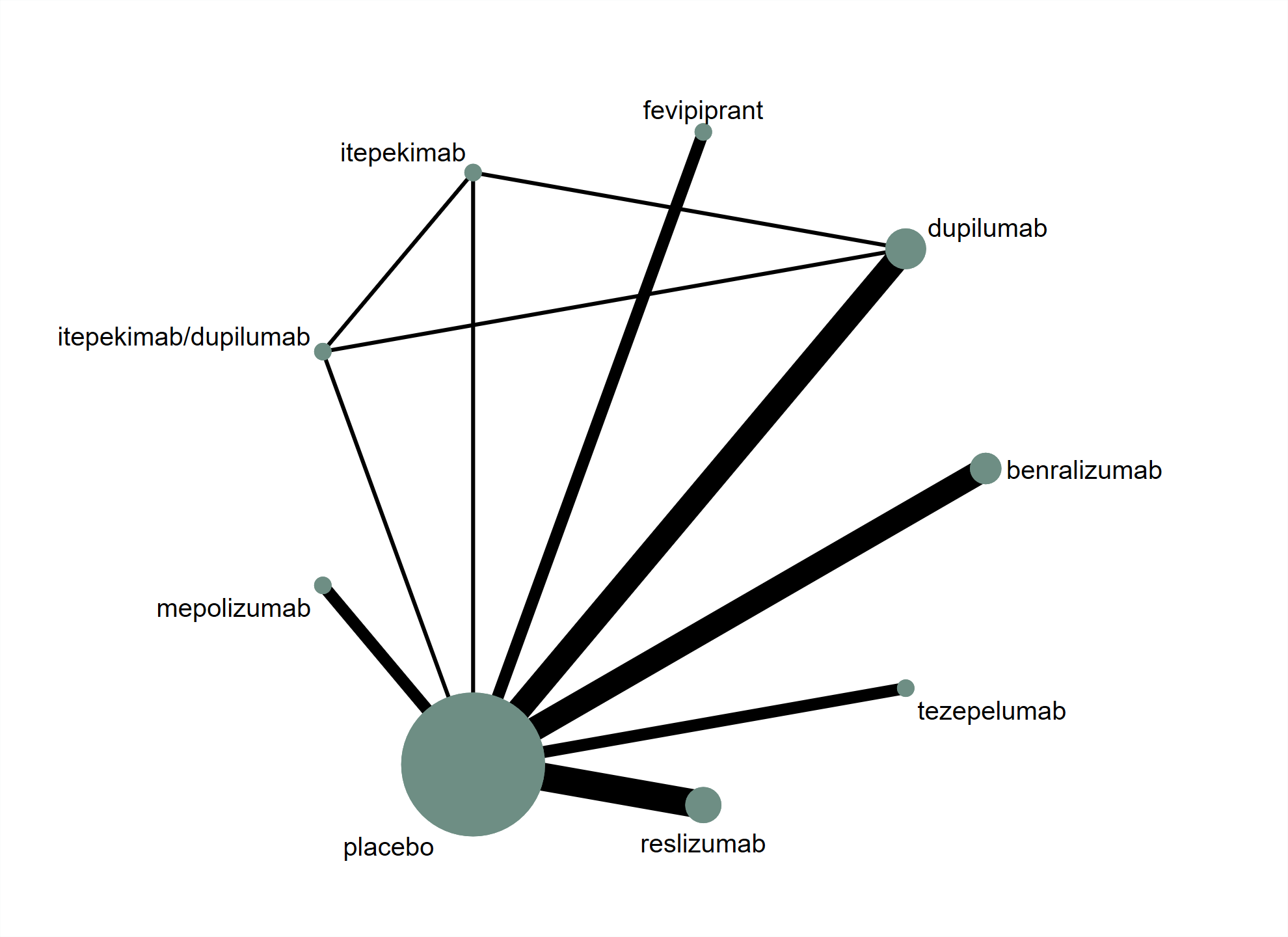

### eFigure 31. FEV1 (high eos) network forest plot

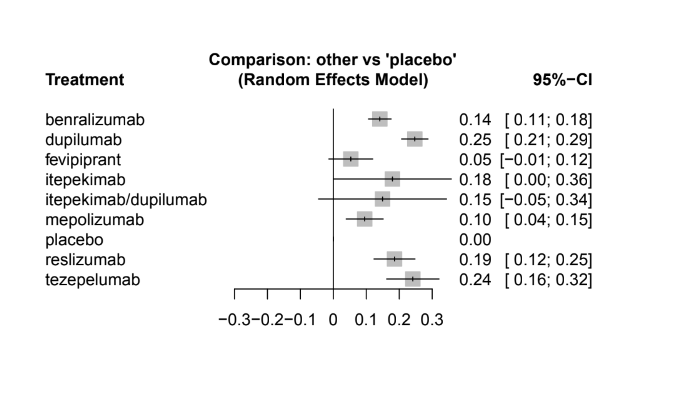

### eFigure 32. FEV1 (high eos) pairwise forest plot

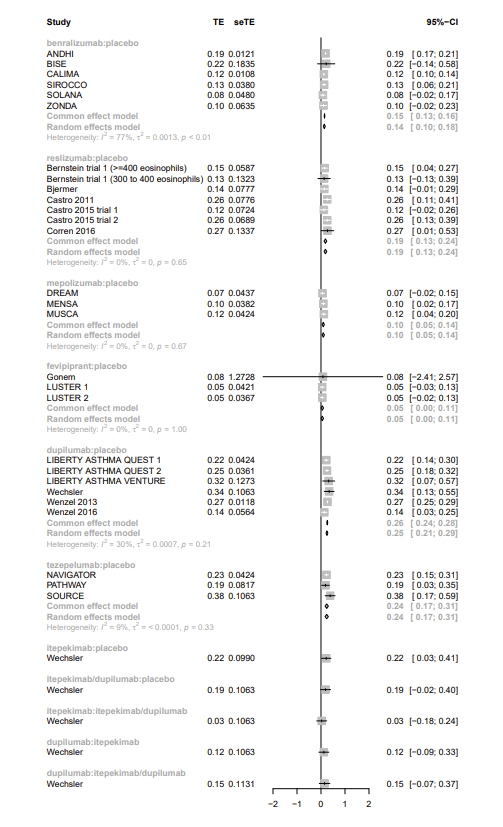

### eFigure 33. FEV1 (high eos) funnel plot

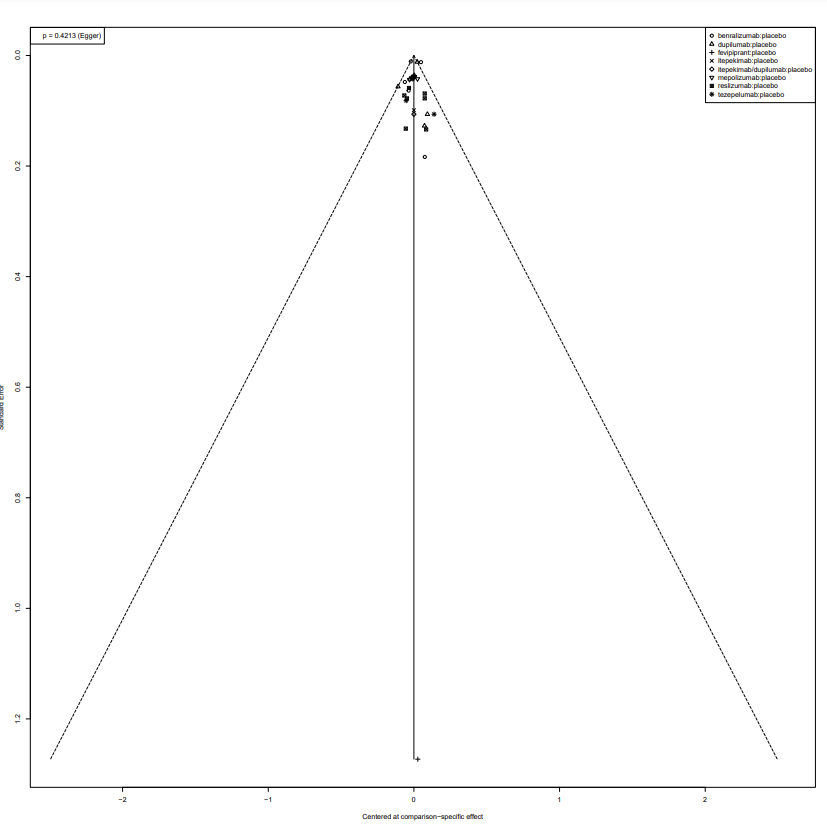

### eFigure 34. FEV1 (low eos) network diagram

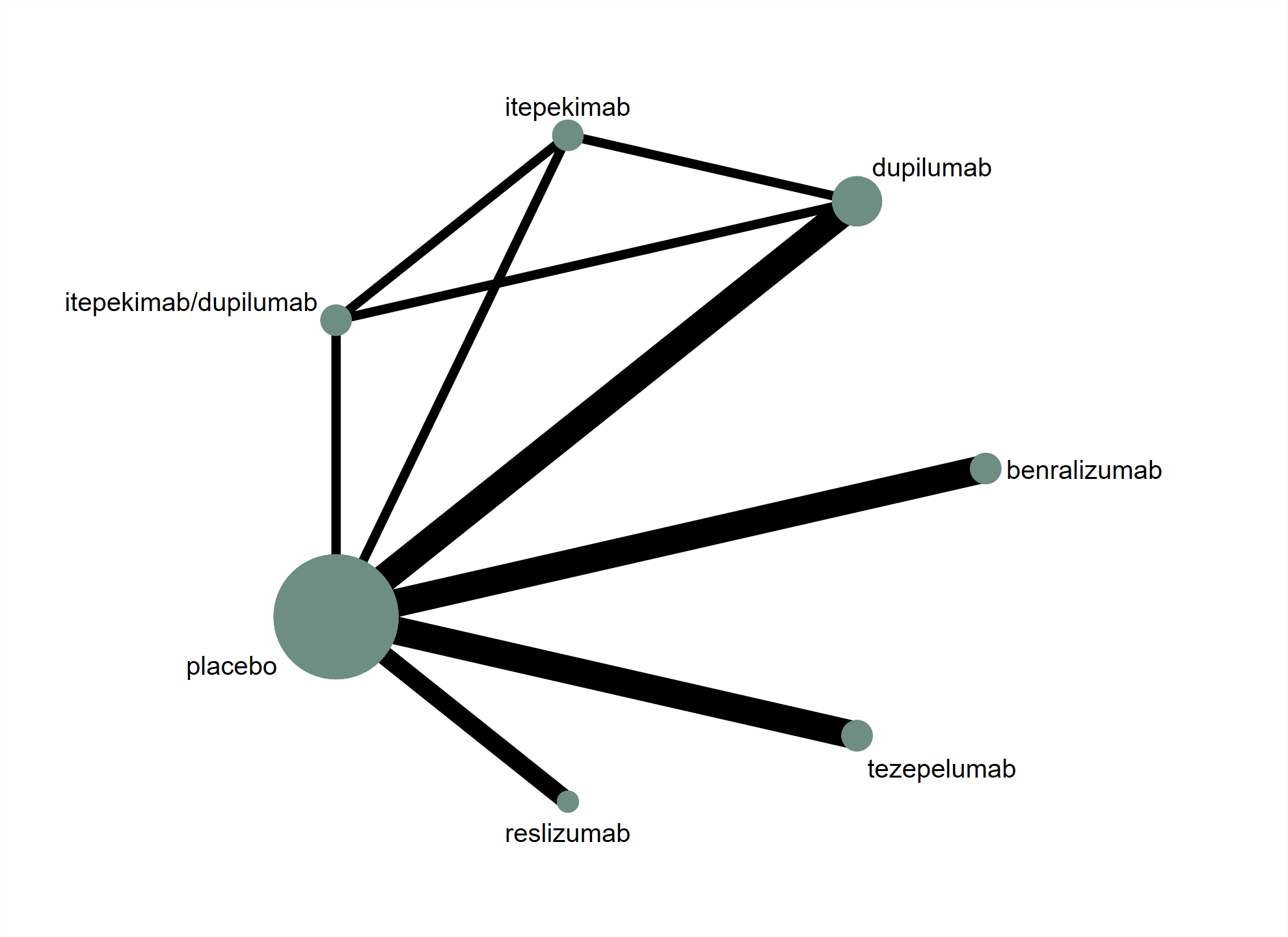

### eFigure 35. FEV1 (low eos) network forest plot

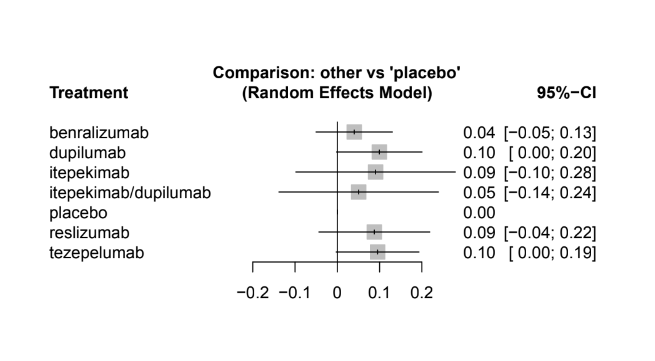

### eFigure 36. FEV1 (low eos) pairwise forest plot

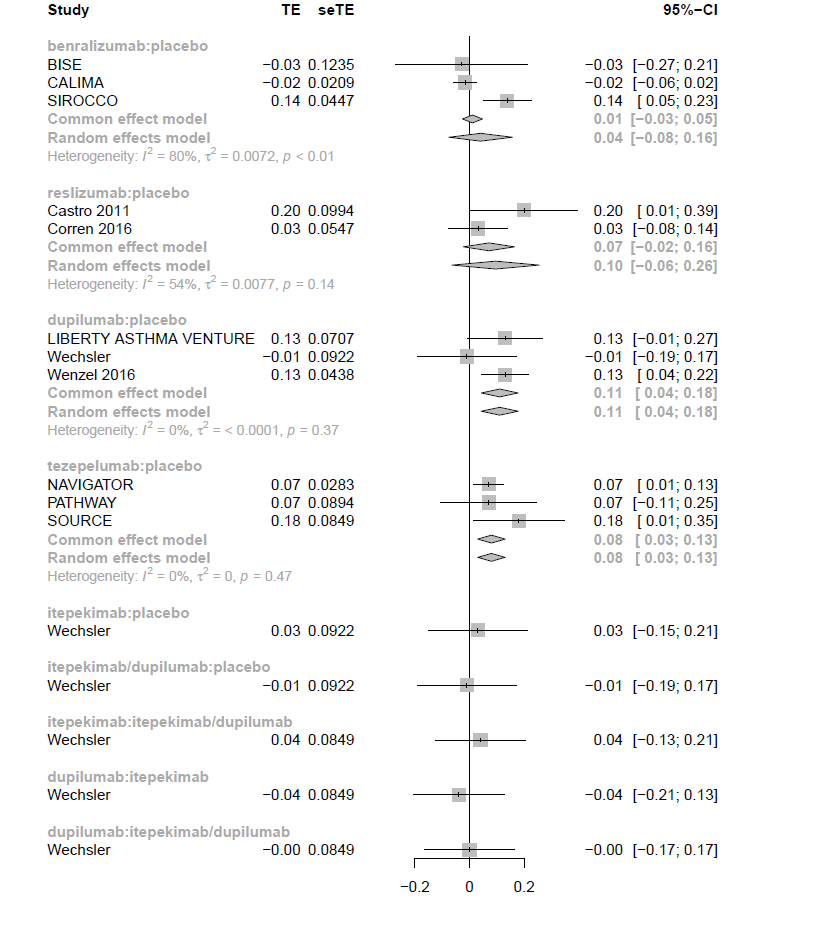

### eFigure 37. FEV1 (low eos) funnel plot

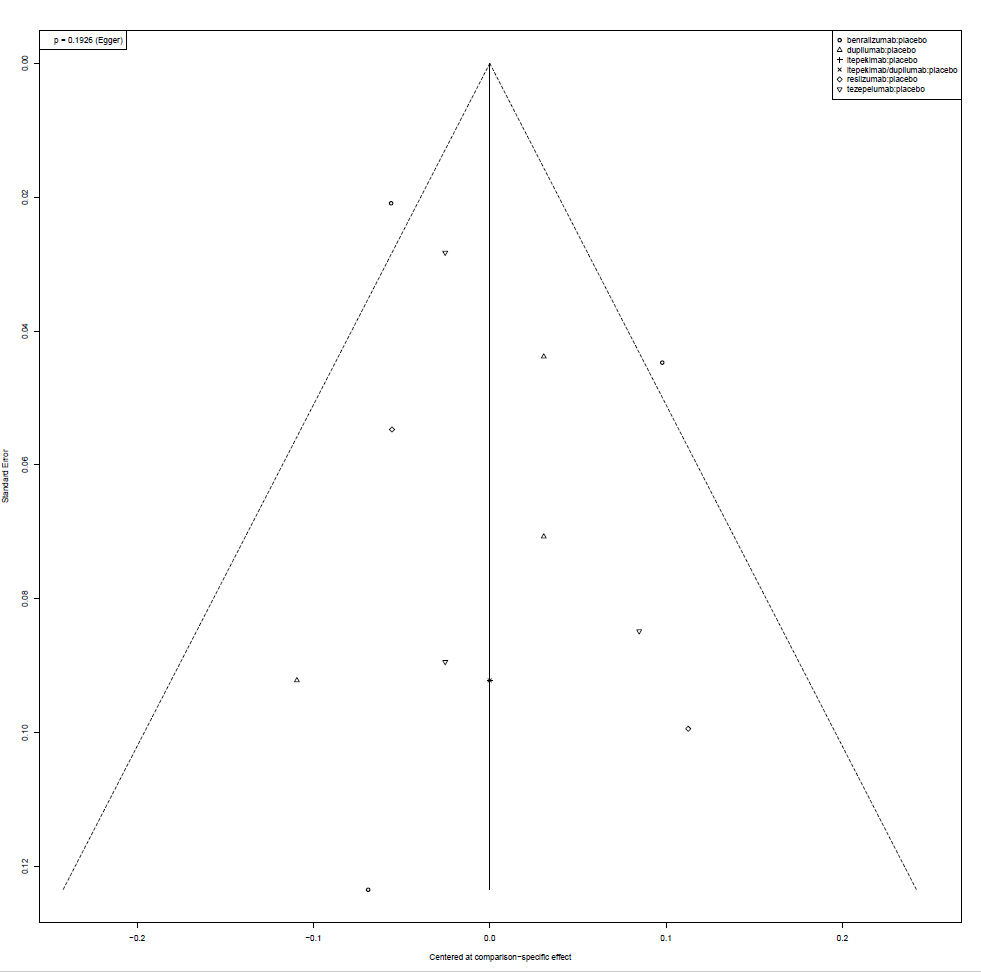

### eFigure 38. Hospitalizations network diagram

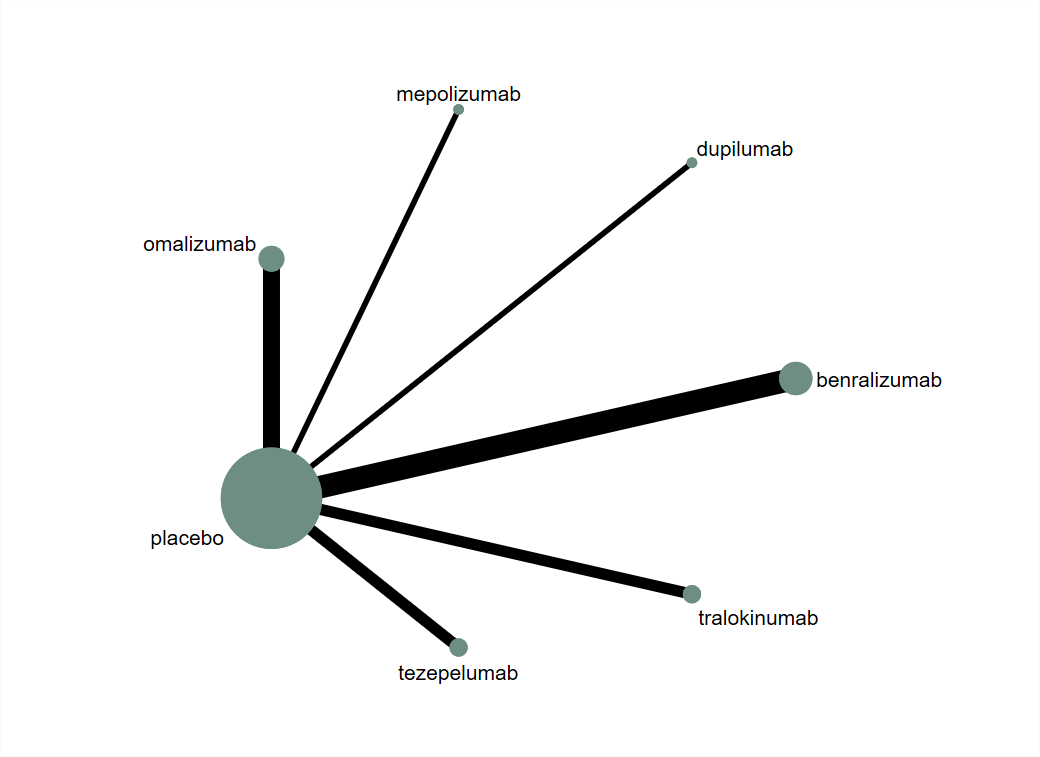

### eFigure 39. Hospitalizations forest plot

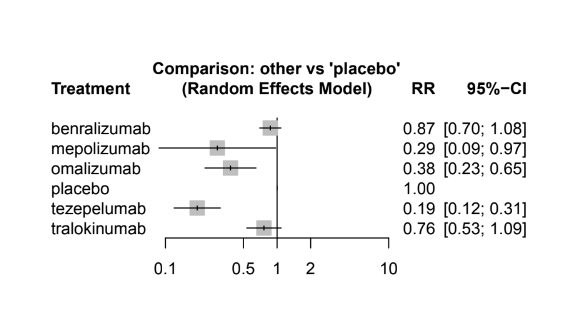

### eFigure 40. Hospitalizations pairwise forest plot

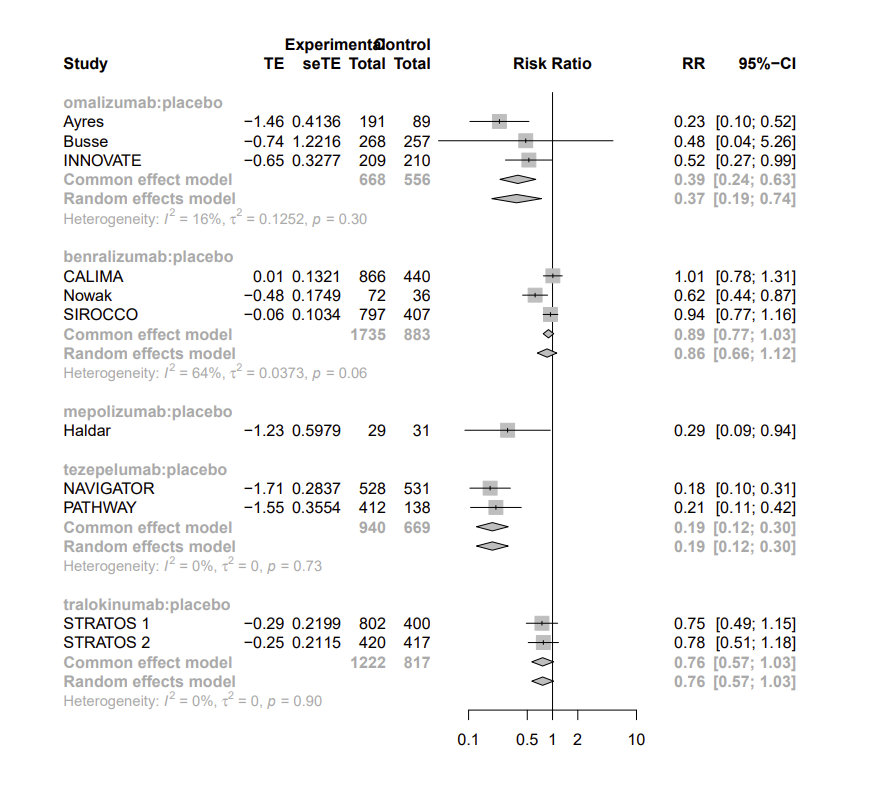

### eFigure 41. Hospitalizations funnel plot

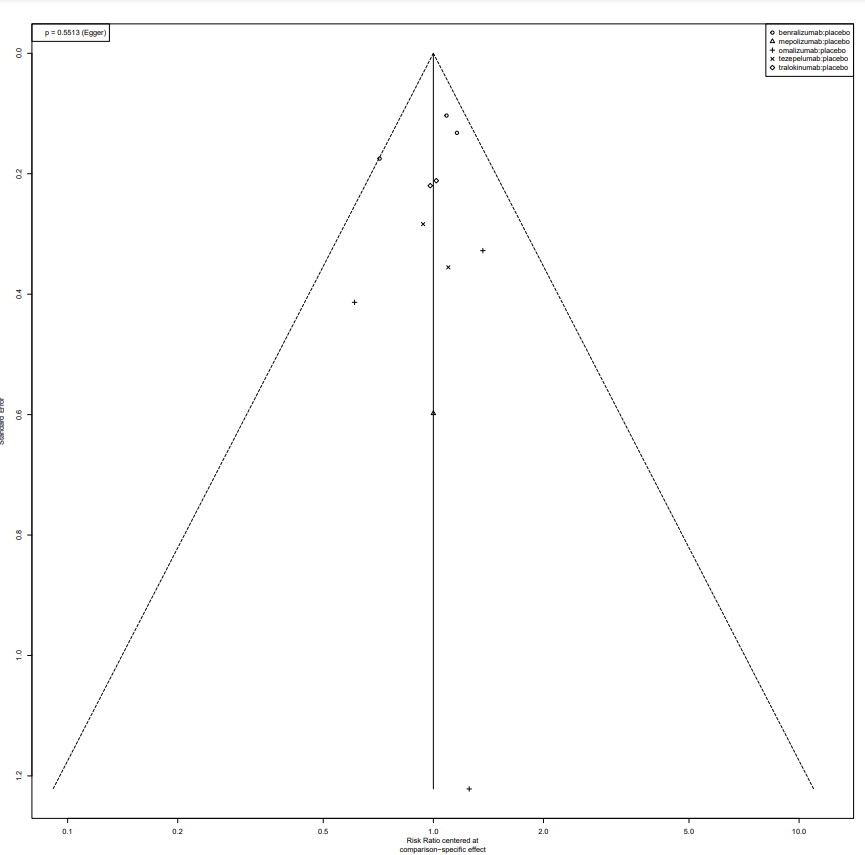

### eFigure 42. Reduction in use of oral corticosteroids network diagram

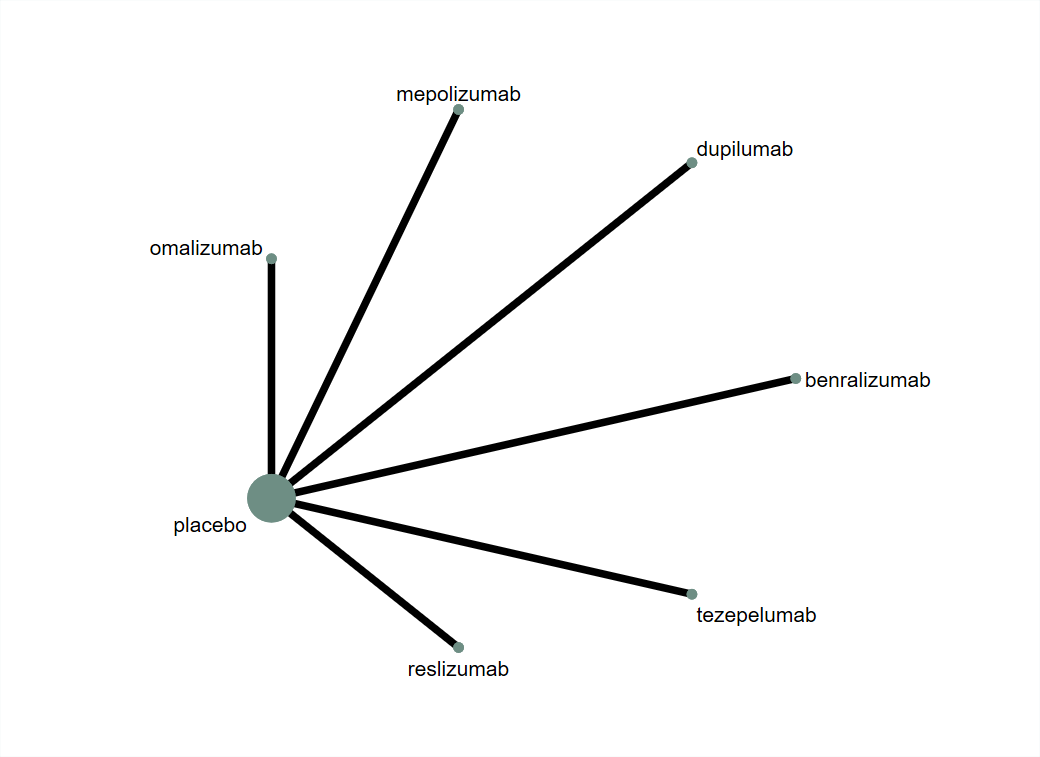

### eFigure 43. Reduction in use of oral corticosteroids network forest plot

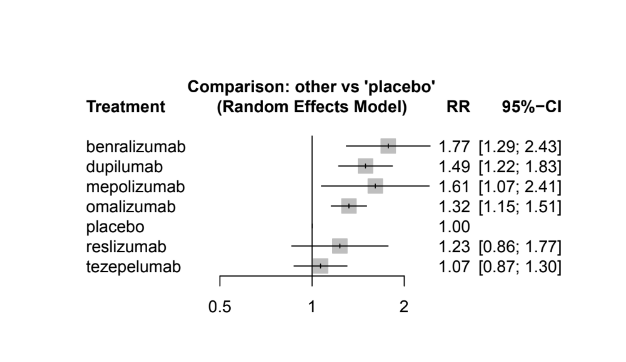

### eFigure 44. Reduction in use of oral corticosteroids pairwise forest plot

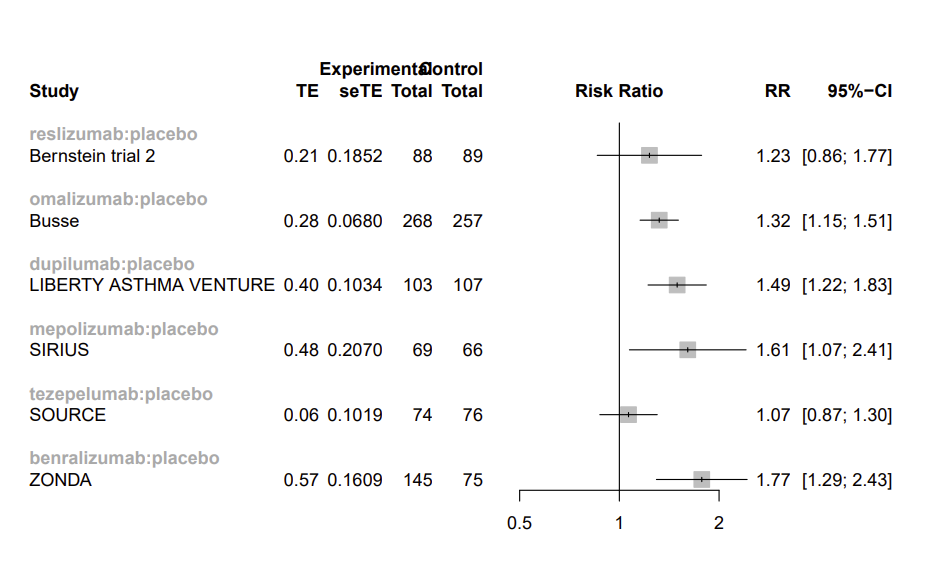

### eFigure 45. Reduction in use of oral corticosteroids funnel plot

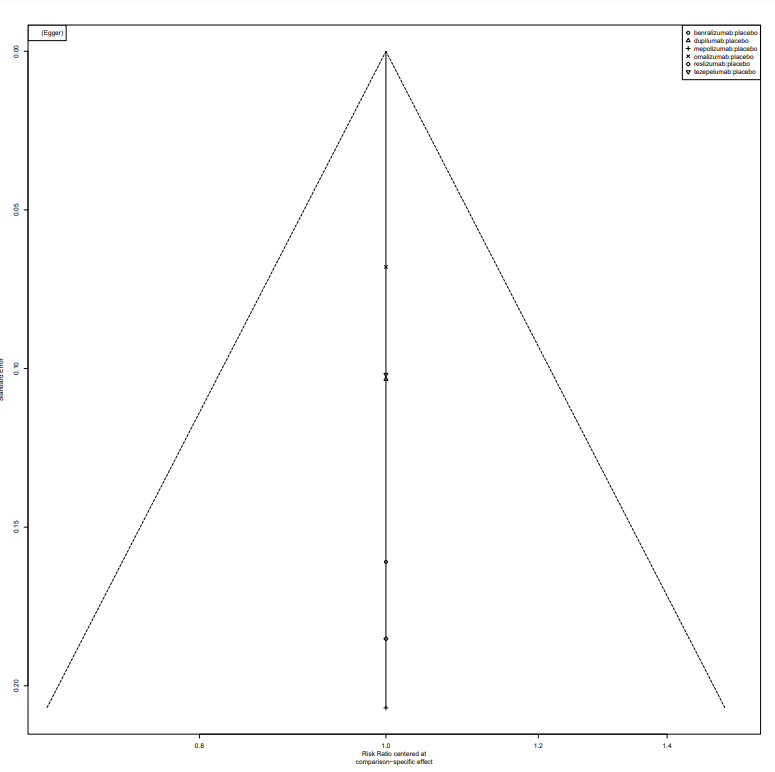

### eFigure 46. Adverse events leading to discontinuation network diagram

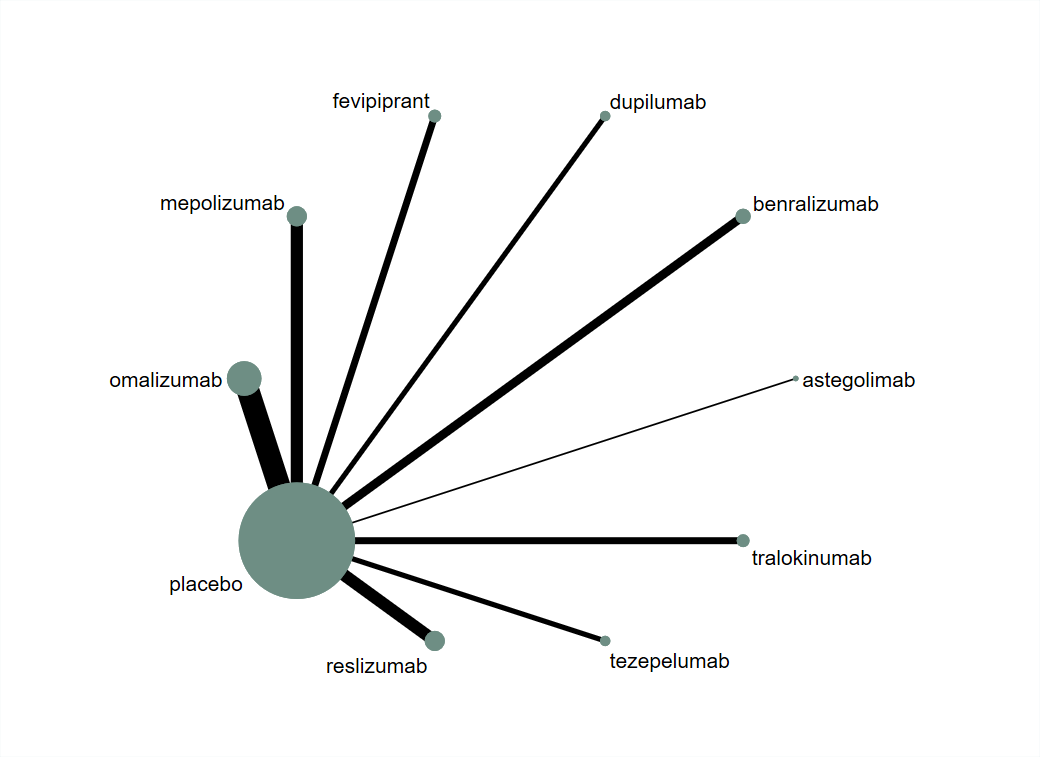

### eFigure 47. Adverse events leading to discontinuation network forest plot

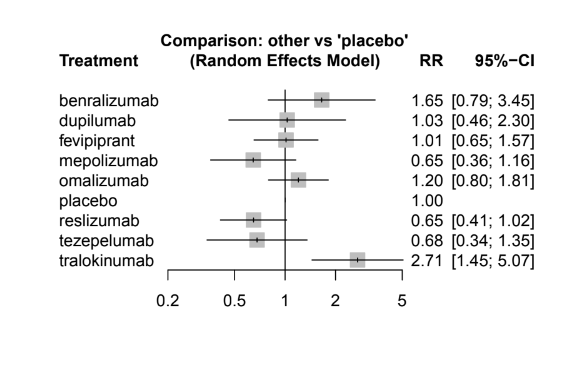

### eFigure 48. Adverse events leading to discontinuation pairwise forest plot

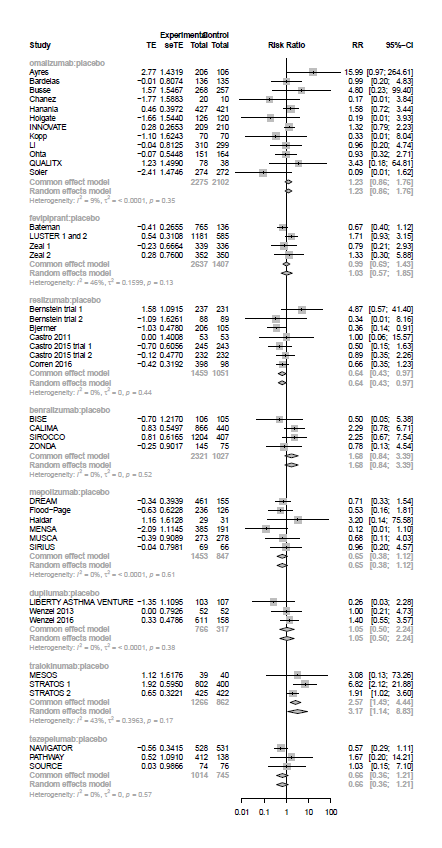

### eFigure 49. Adverse events leading to discontinuation funnel plot

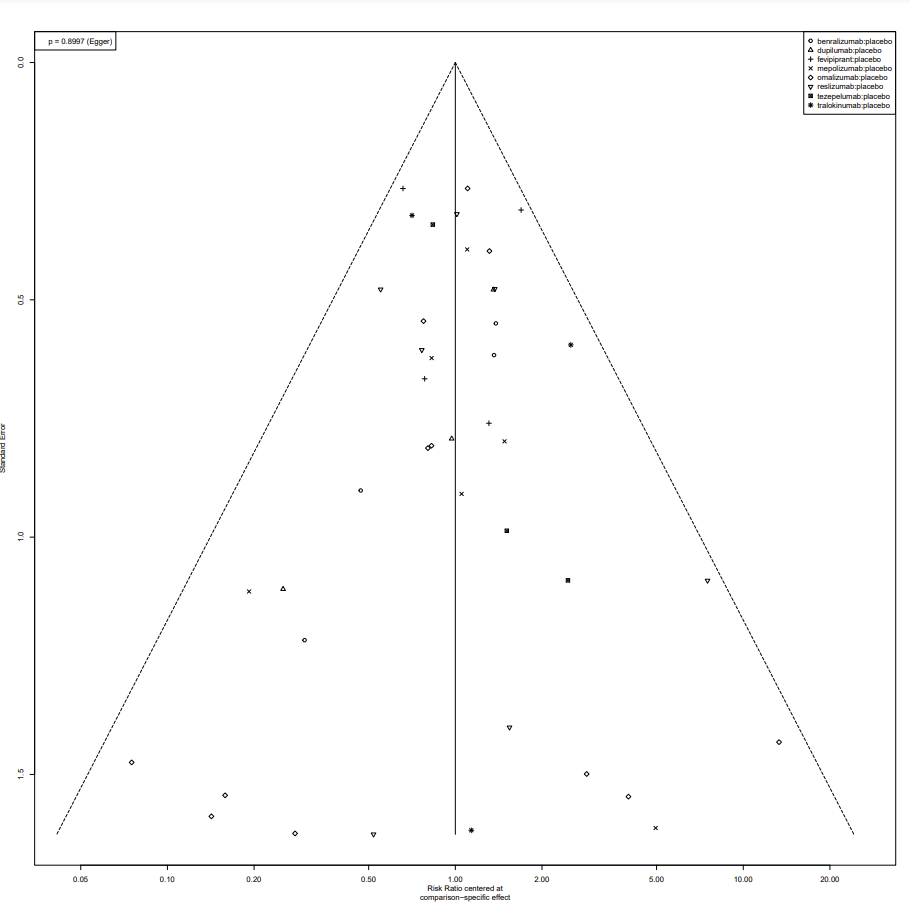

### eFigure 50. Exacerbations (all) all severities vs moderate-to-severe only

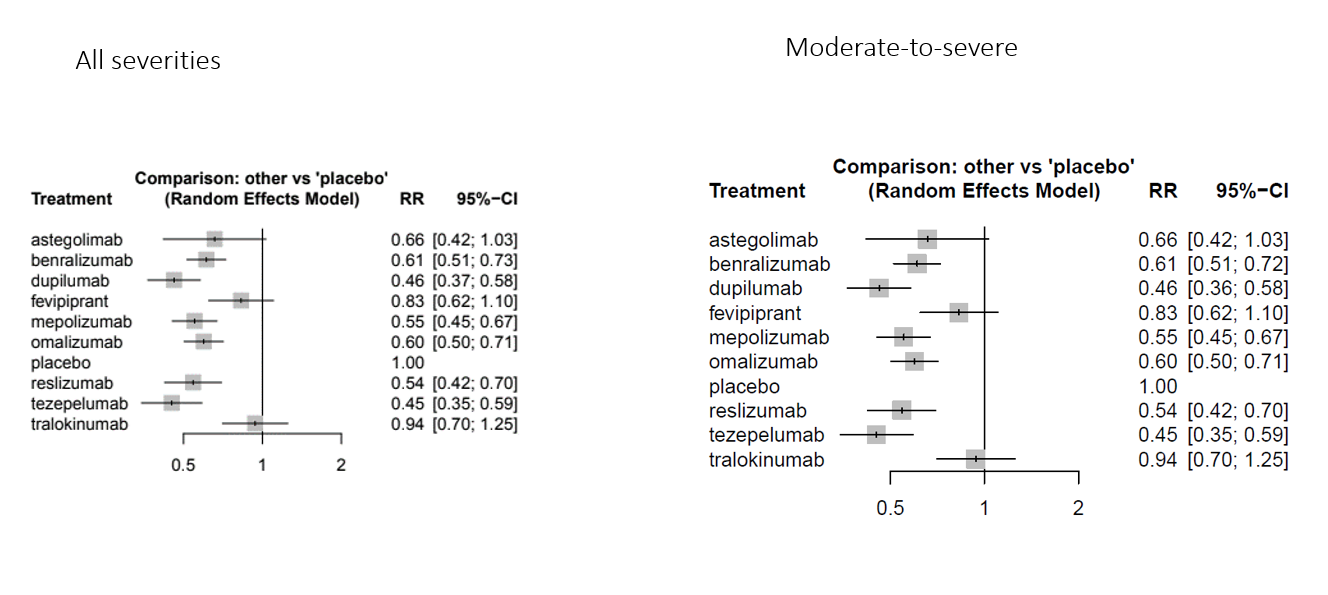

### eFigure 51. Exacerbations (high eos) all severities vs moderate-to-severe only

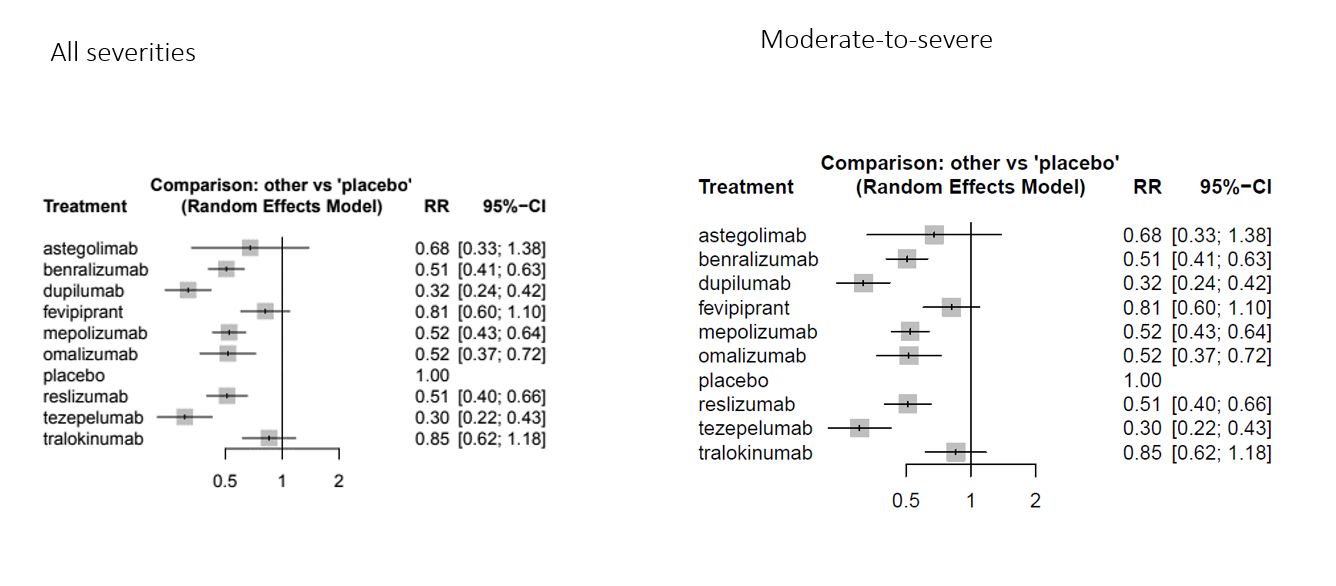

### eFigure 52. Exacerbations (low eos) all severities vs moderate-to-severe only

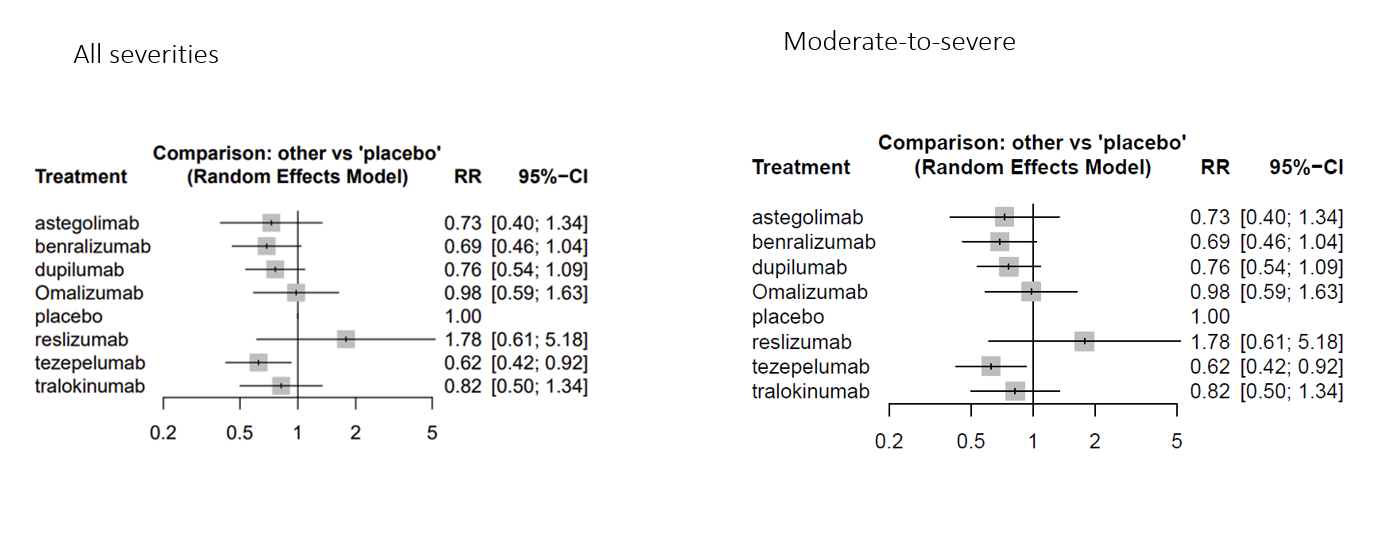

### eFigure 53. ACQ (all) all severities vs moderate-to-severe only

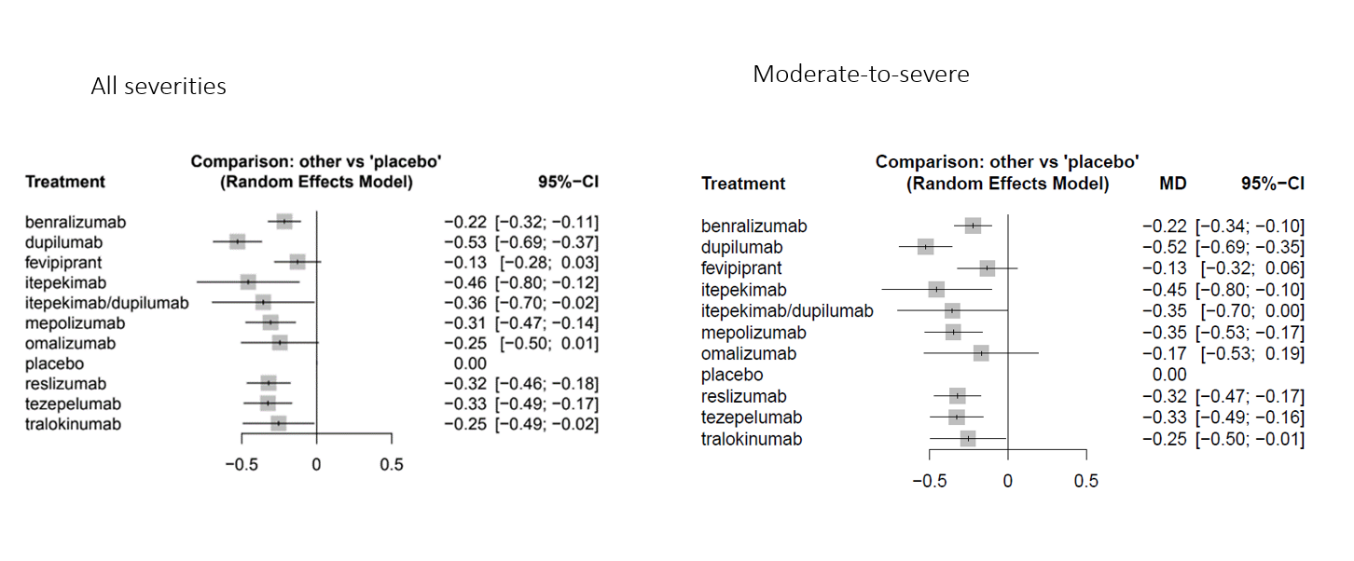

### eFigure 54. FEV1 (all) all severities vs moderate-to-severe only

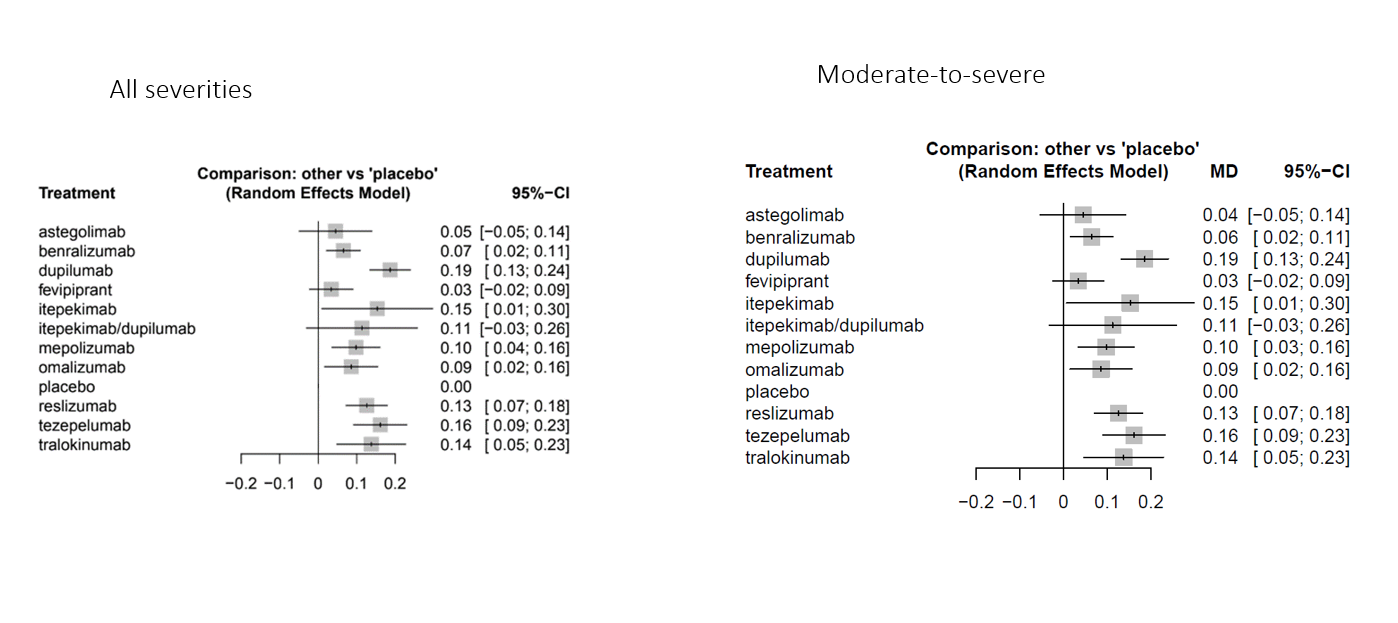

### eFigure 55. FEV1 (high eos) all severities vs moderate-to-severe only

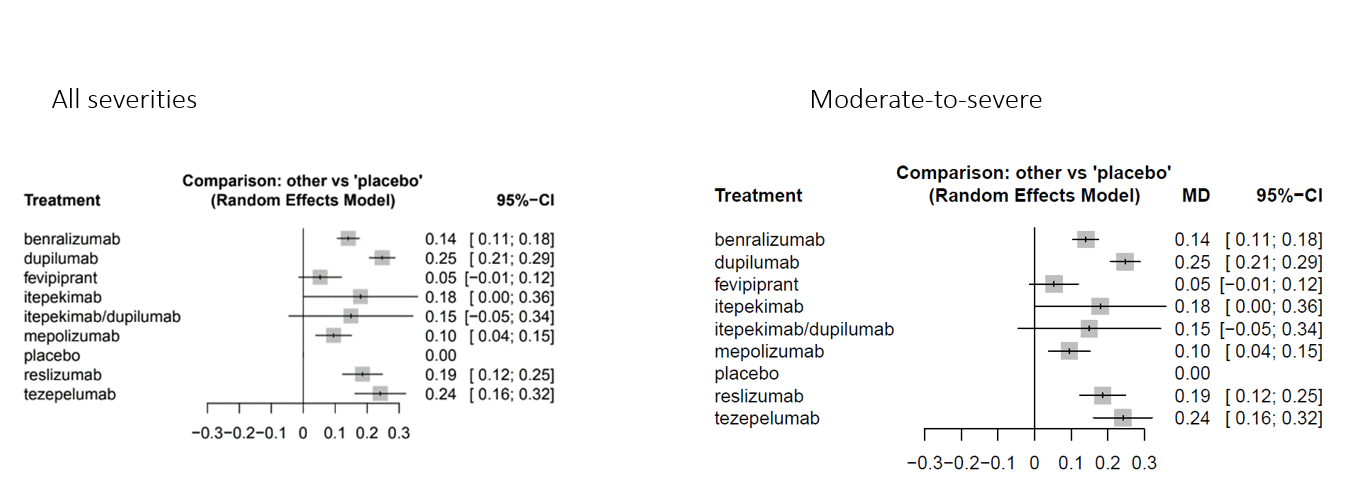

### eFigure 56. FEV1 (low eos) all severities vs moderate-to-severe only

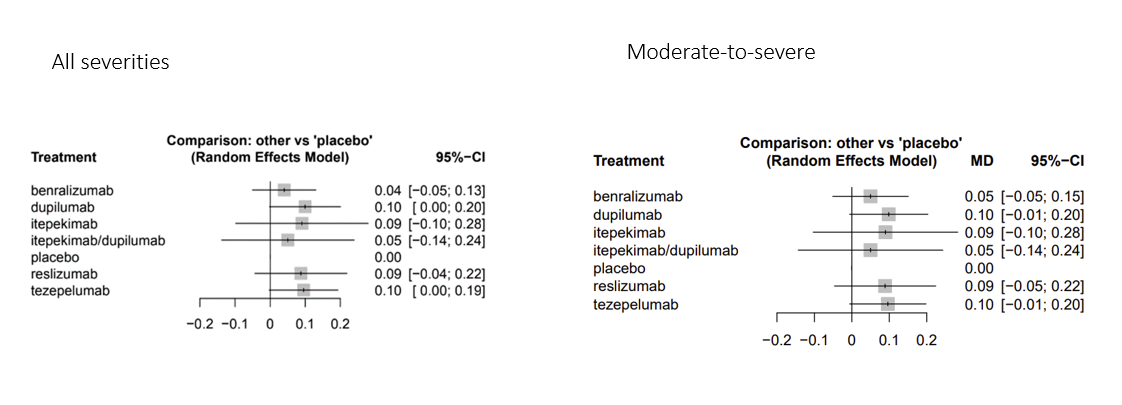

### eFigure 57. Hospitalizations severities vs moderate-to-severe only

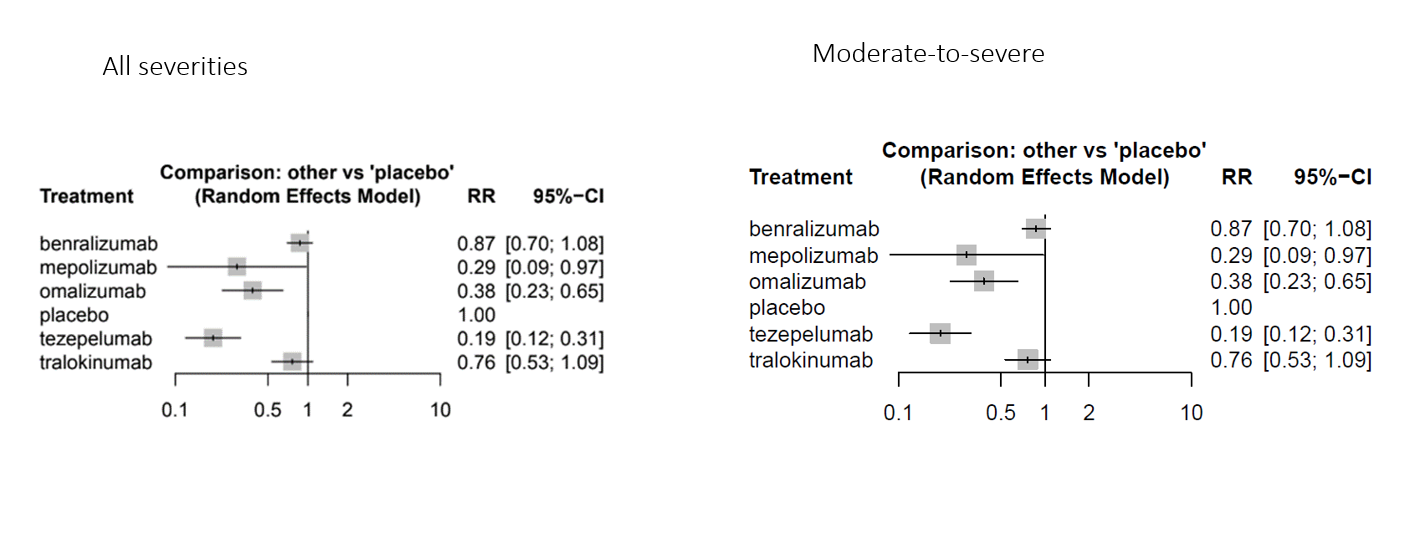

### eFigure 58. Adverse events leading to discontinuation all severities vs moderate-to-severe only

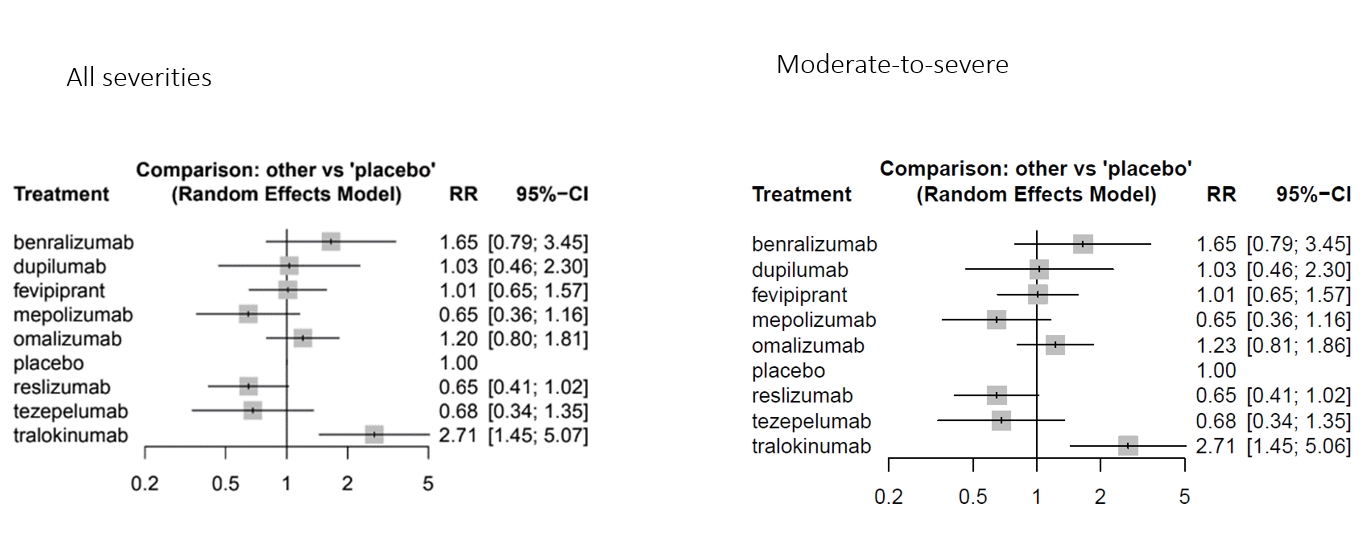
